# Evidence-based recommendations for gene-specific ACMG/AMP variant classification from the ClinGen ENIGMA BRCA1 and BRCA2 Variant Curation Expert Panel

**DOI:** 10.1101/2024.01.22.24301588

**Authors:** Michael T. Parsons, Miguel de la Hoya, Marcy E. Richardson, Emma Tudini, Michael Anderson, Windy Berkofsky-Fessler, Sandrine M. Caputo, Raymond C. Chan, Melissa C. Cline, Bing-Jian Feng, Cristina Fortuno, Encarna Gomez-Garcia, Johanna Hadler, Susan Hiraki, Megan Holdren, Claude Houdayer, Kathleen Hruska, Paul James, Rachid Karam, Huei San Leong, Alexandra Martins, Arjen R. Mensenkamp, Alvaro N. Monteiro, Vaishnavi Nathan, Robert O’Connor, Inge Sokilde Pedersen, Tina Pesaran, Paolo Radice, Gunnar Schmidt, Melissa Southey, Sean Tavtigian, Bryony A. Thompson, Amanda E. Toland, Clare Turnbull, Maartje J. Vogel, Jamie Weyandt, George A.R. Wiggins, Lauren Zec, Fergus J. Couch, Logan C. Walker, Maaike P. G. Vreeswijk, David E. Goldgar, Amanda B. Spurdle

## Abstract

The ENIGMA research consortium (https://enigmaconsortium.org/) develops and applies methods to determine clinical significance of variants in Hereditary Breast and Ovarian Cancer genes. An ENIGMA *BRCA1/2* classification sub-group, originally formed in 2016 as a ClinGen external expert panel, evolved into a ClinGen internal Variant Curation Expert Panel (VCEP) to align with Federal Drug Administration recognized processes for ClinVar contributions.

The VCEP reviewed American College of Medical Genetics/Association of Molecular Pathology (ACMG/AMP) classification criteria for relevance to interpreting *BRCA1* and *BRCA2* variants. Statistical methods were used to calibrate evidence strength for different data types. Pilot specifications were tested on 40 variants, and documentation revised for clarity and ease-of-use.

The original criterion descriptions for 13 evidence codes were considered non-applicable or overlapping with other criteria. Scenario of use was extended or re-purposed for eight codes. Extensive analysis and/or data review informed specification descriptions and weights for all codes. Specifications were applied to pilot variants with pre-existing ClinVar classification as follows: 13 Uncertain Significance or Conflicting, 14 Pathogenic and/or Likely Pathogenic, and 13 Benign and/or Likely Benign. Review resolved classification for 11/13 Uncertain Significance or Conflicting variants, and retained or improved confidence in classification for the remaining variants.

Alignment of pre-existing ENIGMA research classification processes with ACMG/AMP classification guidelines highlighted several gaps in both the research processes and the baseline ACMG/AMP criteria. Calibration of evidence types was key to justify utility and strength of different evidence types for gene-specific application. The gene-specific criteria demonstrated value for improving ACMG/AMP-aligned classification of *BRCA1* and *BRCA2* variants.

## Introduction

The role of *BRCA1* (MIM 113705) and *BRCA2* (MIM 600185) in Hereditary Breast and Ovarian Cancer (HBOC) has long been recognized, with genetic testing initiated soon after discovery of these genes in the 1990s.^1; 2^ The ENIGMA international research consortium (https://enigmaconsortium.org/)^3^ focuses on development and application of methods to determine the clinical significance of sequence variants in HBOC genes. The consortium has members from six continents that provide a broad range of expertise, under the umbrellas of analytical, splicing, functional, pathology and clinical working groups, for translational research projects. At the request of ClinGen, in 2016 ENIGMA formed an external expert panel for curation of *BRCA1* and *BRCA2* variants. The classification criteria documented for this purpose (no longer used, Supplemental Information, Section 1) captured qualitative criteria generally adopted clinically (e.g., most premature termination codon variants were assumed to be pathogenic), and quantitative multifactorial likelihood analysis methods developed in the research setting.^4–8^ The key component of the multifactorial likelihood approach is the statistical calibration of independent data types using variants of known pathogenicity status, to inform the weight of evidence towards or against pathogenicity. The external expert panel guidelines were then used to assign 7,456 expert curations for *BRCA1 and BRCA2* variants in ClinVar.

In parallel to these efforts, there was increasing international uptake of variant classification guidelines published by the American College of Medical Genetics and Genomics/Association for Molecular Pathology (ACMG/AMP)^9^ for diagnostic interpretation of germline sequence variants, with applicability to any Mendelian disease gene. In 2020, the ENIGMA external expert panel sought to become an internal ClinGen Variant Curation Expert Panel (VCEP),^10; 11^ to align with Federal Drug Agency (FDA) recognized processes for expert panel contributions to ClinVar. Here, we provide an overview of the evidence-based approach taken to consider relevance of each ACMG/AMP evidence code for curation of variants in *BRCA1* and *BRCA2*, and report pilot study results demonstrating the value of detailed (gene-specific) specifications to assist variant curation and resolve discordances and uncertainty in variant classification.

## Methods

The establishment and activities of the ClinGen *BRCA1* and *BRCA2* VCEP followed the ClinGen FDA-recognised approval process (https://clinicalgenome.org/docs/guidelines-for-applying-for-variant-or-gene-curation-expert-panel-status/), with reference to Protocol Version 8 at the time of VCEP initiation.

The original ENIGMA *BRCA1* and *BRCA2* expert panel membership, which was largely comprised of representatives from major national clinical and research initiatives in Australia, Europe and USA, was expanded to include representatives from several major diagnostic testing laboratories from the United States with extensive experience in the application of ACMG/AMP guidelines. The resulting ClinGen ENIGMA *BRCA1* and *BRCA2* VCEP consists of research and clinical experts from around the world, including Australasia, Europe and the United States. VCEP members met approximately monthly to review the baseline ACMG/AMP sequence variant classification guidelines^9^ to determine whether each classification criterion should be adopted, modified, or omitted for *BRCA1* and *BRCA2* variant interpretation.

Both *BRCA1* and *BRCA2* were designated as genes for which loss of function is a known mechanism of disease. Reference sequences used for annotation are as follows:

*BRCA1.* Coding DNA reference sequence from genomic RefSeq NG_005905.2 (same as LRG 292, Ensembl ENSG00000012048) covering *BRCA1* transcript NM_007294.4 (Ensembl transcript ENST00000357654.9). Exons are sequentially numbered to match the exon descriptions of the MANE Select transcript (NM_007294.4). Exon numbering of *BRCA1* has historically been according to GenBank U14680.1, with exon 4 missing due to a correction made after the initial description of the gene, termed here as legacy exon numbering.

*BRCA2.* Coding DNA reference sequence from genomic RefSeq NG_012772.3 (same as LRG 293, Ensembl ENSG00000139618), covering *BRCA2* transcript NM_000059.4 (MANE Select transcript; Ensembl transcript ENST00000380152.8).

The classification tiers in pre-existing ENIGMA external panel classification criteria for *BRCA1* and *BRCA2* (Supplemental Information, Section 1) grouped multiple sources of information (e.g. frequency data, variant type, tumor pathology, co-occurrence with a pathogenic variant, etc). A critical aspect of VCEP activities was to convert these grouped criteria to align with ACMG/AMP codes representing different classification criteria, falling under the broad evidence types described for the ACMG/AMP framework (i.e. population, computation/predictive, functional, segregation, de novo, allelic, other).^9^ Statistical methods were used to calibrate strength of evidence (i.e. supporting, moderate, strong, very strong and stand-alone) for different data types following approaches as outlined previously; and likelihood ratio (LR) estimates towards or against pathogenicity were derived for a given evidence type ^12^ and used to assign weights for or against pathogenicity following recommendations arising from Bayesian modelling of the ACMG/AMP guidelines.^13^ Alongside, key members of the ClinGen Sequence Variant Interpretation Working Group (SVI WG) (https://clinicalgenome.org/working-groups/sequence-variant-interpretation/) were consulted about how to capture valuable information sources and analytical approaches that were used previously for external expert panel classification, but that did not strictly conform to ACMG/AMP evidence types and designated codes/strengths. Extensive documentation supporting the rationale for application and weighting of each code was compiled for ClinGen SVI WG review, following the standard VCEP approval protocol. Specifications for codes relating to the use of computational and experimental evidence relevant to variant impact on RNA splicing were informed by parallel development of recommendations from the ClinGen SVI Splicing Subgroup.^14^ After addressing feedback from the SVI WG, the draft documentation was provided to nine VCEP members who had self-nominated to act as biocurators. As biocurators they review and evaluate evidence relevant for variant classification, assign relevant ACMG/AMP codes and weights for the available evidence, and ascribe a final pathogenicity classification based on the information reviewed. The draft specifications were tested on 40 pilot variants, selected to capture variants spanning different assumed molecular impact, and different pre-existing classifications in ClinVar (Supplemental Information, Section 2, Table S1). VCEP members were requested to provide any internal data of relevance for classification of these 40 variants. Initial ClinVar summary classification descriptions were as follows: Pathogenic (P), n=11; Likely Pathogenic/Pathogenic (LP/P), n=3; Uncertain Significance (VUS), n=4; Benign/Likely Benign (B/LB), n=1; Benign (B), n=12; Conflicting, n=9. Conflicting Classifications represented various combinations of individual submitter classifications (details provided in Table S1).

To facilitate the curation process, each biocurator was provided a file with the following variant-specific information: population frequency as reported in gnomAD (v2.1 exomes only and v3.1); existing multifactorial LR data (spanning segregation, family history, tumour pathology, case-control and co-occurrence LRs); protein functional assay data; mRNA splicing assay data; bioinformatic impact predictions (missense, in-frame, splicing); clinical features of Fanconi Anemia (FA) cases as drawn from the literature and additional internal laboratory data relevant for classification as provided by VCEP members. The latter included splicing assay results, co-segregation data, presence or absence of FA phenotype for individuals with co-occurring variants. Protein functional assay data was provided with functional category (impact, no impact or partial/indeterminate) assigned for all functional assays considered relevant, with a summary description of the combined results. RNA assay data required evaluation by individual biocurators to assign code weights. Each variant was curated by the lead biocurator (MTP) and two additional biocurators. The lead biocurator reviewed curations for consistency in code application (including code strength), and final variant classification. Collated findings were discussed with all VCEP members to identify factors contributing to between-biocurator differences in use of the specifications.

After this initial phase of variant review, biocurator feedback was used to inform revision of the documentation for clarity and ease of use. This included development of simplified look-up tables. At this time codes relating to bioinformatic predictions were updated to allow three categories: evidence towards pathogenicity, against pathogenicity and no bioinformatic code applicable. These updates were based on results from published splicing prediction analyses^14^, and VCEP-specific re-analysis conducted to refine calibrations for missense prediction.

The revised documentation was then used for a second phase of the pilot curation. Variants classified with inter-biocurator differences including at least one VUS and one non-VUS classification, labelled as “VUS/other”, were reviewed by two additional VCEP biocurators. Variants with classification confidence differences (P versus LP, or B versus LB) were reviewed by one independent biocurator with extensive experience from the ClinGen *TP53* VCEP. Finally, code assignment was checked for all variants with concordant classification from the first pilot phase, by two VCEP biocurators. Further minor revisions were introduced following biocurator feedback on the revised documentation, and after final review from the ClinGen SVI WG.

## Results and Discussion

An overview of the migration of the ENIGMA external expert panel to current operation as a ClinGen-approved VCEP, following FDA-recognized processes, is shown in Figure 1. Development and documentation of the specifications was an iterative process that involved: (i) discussions and/or review at multiple levels (within the ClinGen SVI WG, and the VCEP members); (ii) coordination with and consideration of other ClinGen activities - including other ClinGen Hereditary Cancer Domain VCEPs, the ClinGen SVI Splicing Subgroup^14^, and a subgroup of the ClinGen SVI WG focussed on calibration of computational tools for missense prediction^15^. The extended timeline reflects the evidence-based approach taken to justify - to both VCEP and ClinGen SVI WG members - the appropriateness and/or strength of different information sources for application to *BRCA1* and *BRCA2* variants.

**Figure 1:**
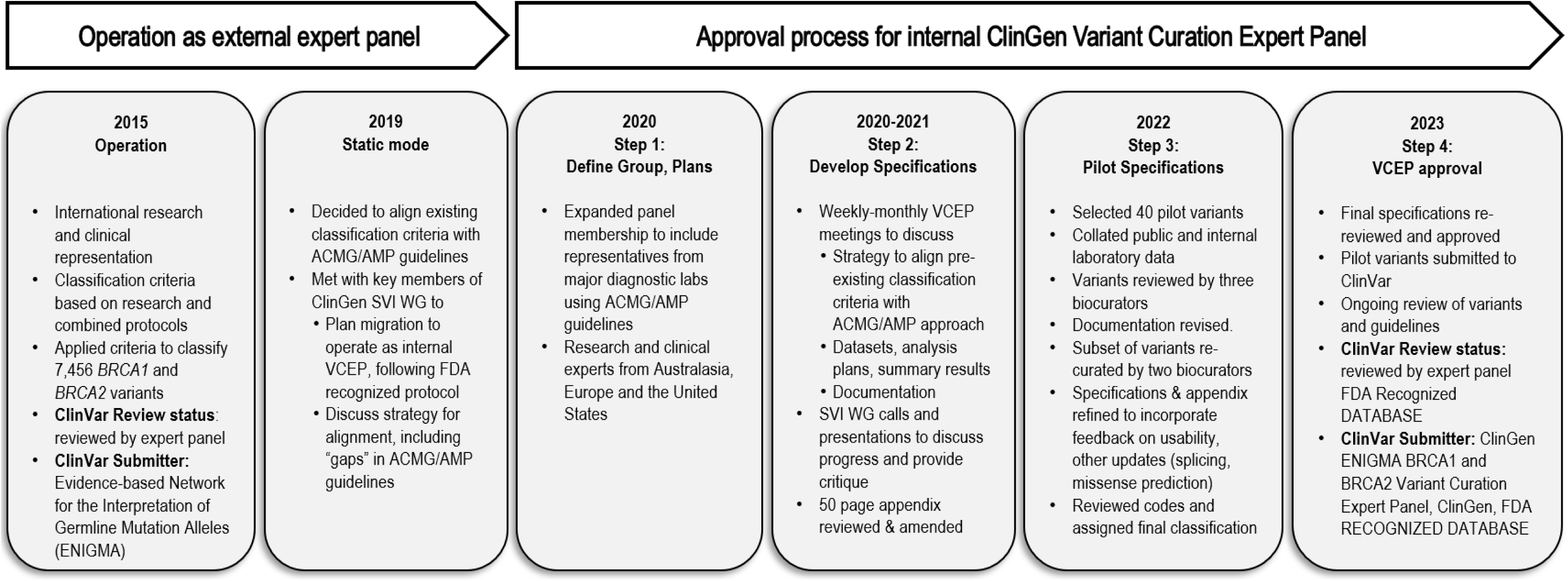
Timeline for migration from “external” expert panel to operation as a ClinGen Variant Curation Expert Panel following the ClinGen FDA-recognized protocol. Discussions with key members of the SVI WG during 2019 relating to the strategy for alignment included the need to capture information sources used previously for *BRCA1* and *BRCA2* variant interpretation that are not explicitly designated under the baseline ACMG/AMP guidelines, especially information used in the context of multifactorial likelihood modelling. Revision of specifications at Step 3 included updates in response to ClinGen SVI Splicing subgroup recommendations^14^, and re-calibration of bioinformatic prediction of missense impact in response to ClinGen SVI Computational subgroup recommendations.^15^ Details of ENIGMA *BRCA1* and *BRCA2* Variant Curation Expert Panel membership and biocurator workforce are available at https://clinicalgenome.org/affiliation/50087/. Pre-existing ENIGMA expert panel classification guidelines for *BRCA1* and *BRCA2* are shown in Supplemental Information, Section 1, as point of reference (though no longer currently used). The final specifications approved for internal ClinGen *BRCA1/2* VCEP use are available via https://cspec.genome.network/cspec/ui/svi/affiliation/50087.

### Overview of *BRCA1* and *BRCA2* Specifications

A broad outline of the evidence informing specifications for each baseline ACMG/AMP code is shown in Figure 2. A summary of the specifications designated for each ACMG/AMP code is described in Supplemental Information, Section 2, Table S2, together with a brief description of mode of application or reasons for excluding a given code. The complete ClinGen SVI WG approved specifications for *BRCA1* and *BRCA2* VCEP are available via the ClinGen C-spec registry (https://cspec.genome.network/cspec/ui/svi/affiliation/50087), together with supporting documentation, and an extensive 50 page document with appendices describing analyses and justifications for code applicability and weighting. These specifications are expected to be updated over time to follow on scientific knowledge progress, and version changes will be documented via this same portal. Summary findings reported in this study refer to Version 1.0 specifications and supporting documentation, also included here as Supplemental Information, Section 3.

**Figure 2:**
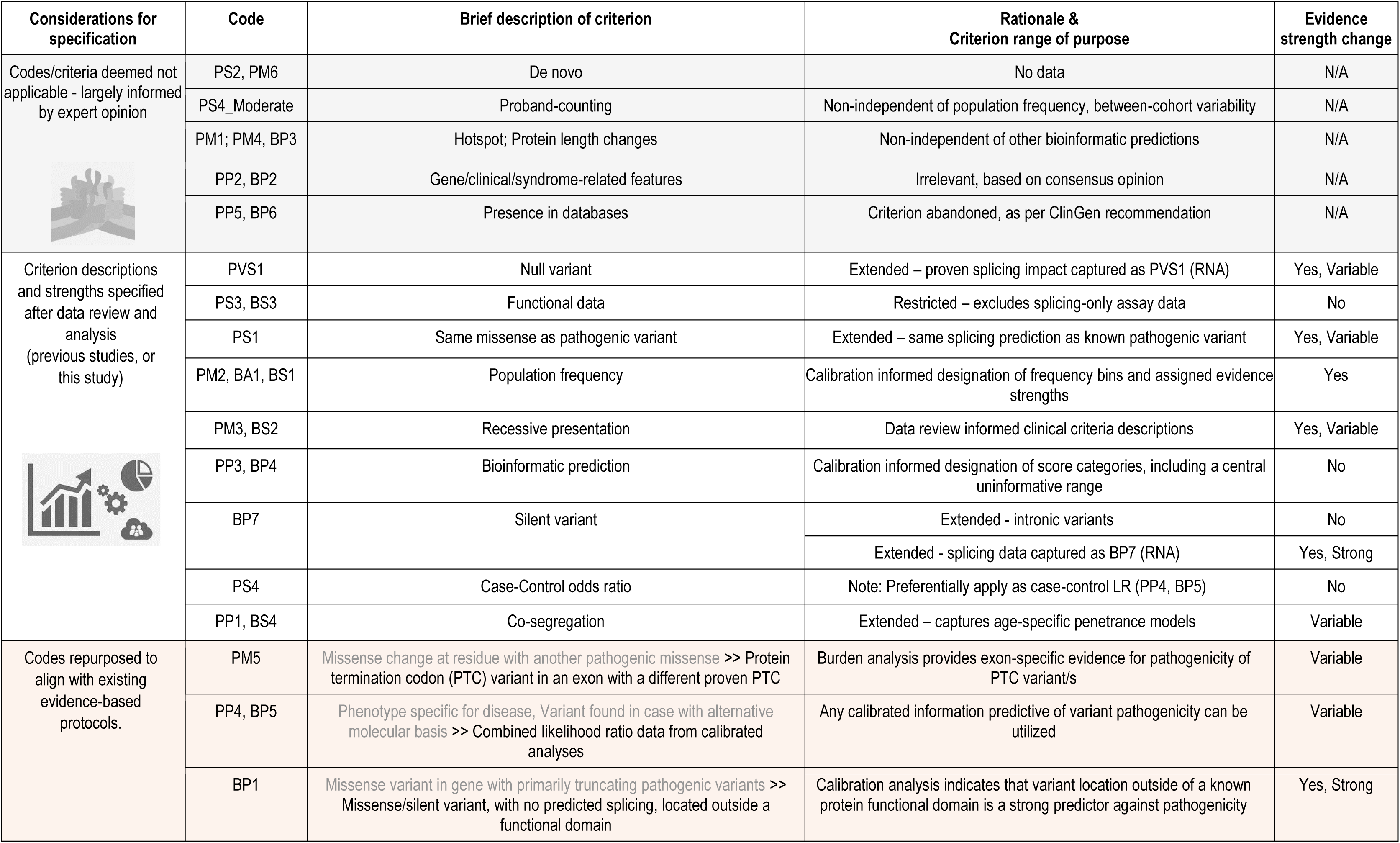
Outline of *BRCA1* and *BRCA2* VCEP specifications designated for baseline ACMG/AMP codes and criteria. Code designations, and original criterion descriptions for codes, are derived from Richards et al 2015.^9^ For repurposed codes, the original criterion description is noted in light blue font, and a broad description is provided for the evidence type to be captured as per specifications. Evidence strength changes are recorded as follows: N/A – not applicable; No - codes used only at original specified weight; Yes - codes used with changed weight; Variable – different strength options can be applicable. Evidence strengths were informed by Bayesian modelling results.^13; 24^ Splicing-related codes were informed by parallel work.^14^

After initial review of ACMG/AMP criteria for relevance to interpretation of *BRCA1* and *BRCA2* variants, the original criterion descriptions for 13 codes were considered non-applicable to these genes or overlapping (non-independent) with other criteria, largely based on expert opinion (Figure 2, Table S2). Specific examples were: PS2/PM6 (de novo), given that *BRCA1/2-*related cancers are common, and there was no information available to calibrate use of this information type; PM1 (location in a hot spot or critical domain), since this was considered to be captured as a component of bioinformatic analysis, since missense prediction tools inherently capture this information. In addition, directed calibration analysis was undertaken to justify that generalised use of proband counting as PS4_Moderate is inappropriate for these genes due to overlap with frequency codes, and variability in evidence strengths observed between cohorts.^16^

Specifications were denoted for 15 codes. Extensive data review and/or analysis from previous ENIGMA-wide and/or VCEP activities was used to inform processes and relevant weights applicable for most of these 15 codes, as described in comprehensive supplementary documentation provided with the VCEP specifications, captured as V 1.0 in Supplemental Information, Section 3. (All versions of specifications are made available via the ClinGen online registry for VCEP specifications. https://cspec.genome.network/cspec/ui/svi/). Several specifications were implemented specifically to follow recommendations from parallel work of the ClinGen SVI Splicing Subgroup^14^: codes PVS1, PS1, and BP7 were extended to capture RNA splicing experimental data or *in silico* predictions; PS3 and BS3 were restricted to capture results from assays that measure protein functional effect (either only protein impact, or protein impact that also measures underlying mRNA impact/s); and splicing impact thresholds defined for SpliceAI were set for bioinformatic prediction of variant impact on splicing, captured under various codes. Probability analysis combined with LR estimation^17^ was used to select and weight bioinformatic tool score categories for missense variant prediction under PP3 and BP4. BayesDel was selected as tool of choice based on results from heterogeneity analysis, performance compared to similar tools, and this tool was able to provide scores for in-frame indels. Extended calibration analysis was undertaken during the pilot phase (Supplemental Information, Section 2) to include an uninformative bioinformatic score range category, and specifically to compare BayesDel score categories to those recommended for general use by Pejaver et al.^15^, published during the VCEP specification process. The optimal binary cutpoint for BayesDel score prediction of impact for a missense variant within a clinically important functional domain was 0.27 for *BRCA1* and 0.20 for *BRCA2* (Figure S1). The binary cutpoint values were used to designate the central point for an uncertain zone comprised of <20% of each reference set, and for which the outer score categories provided at least moderate evidence towards or against pathogenicity based on estimated LR. The VCEP opted, conservatively, to apply this evidence type at supporting weight only (Table 1). Optimal BayesDel score ranges across three categories for both *BRCA1* and *BRCA2* missense prediction (Table 1) did not align with those recommended for generic use by Pejaver et al.^15^ (Table S3).

**Table 1:**
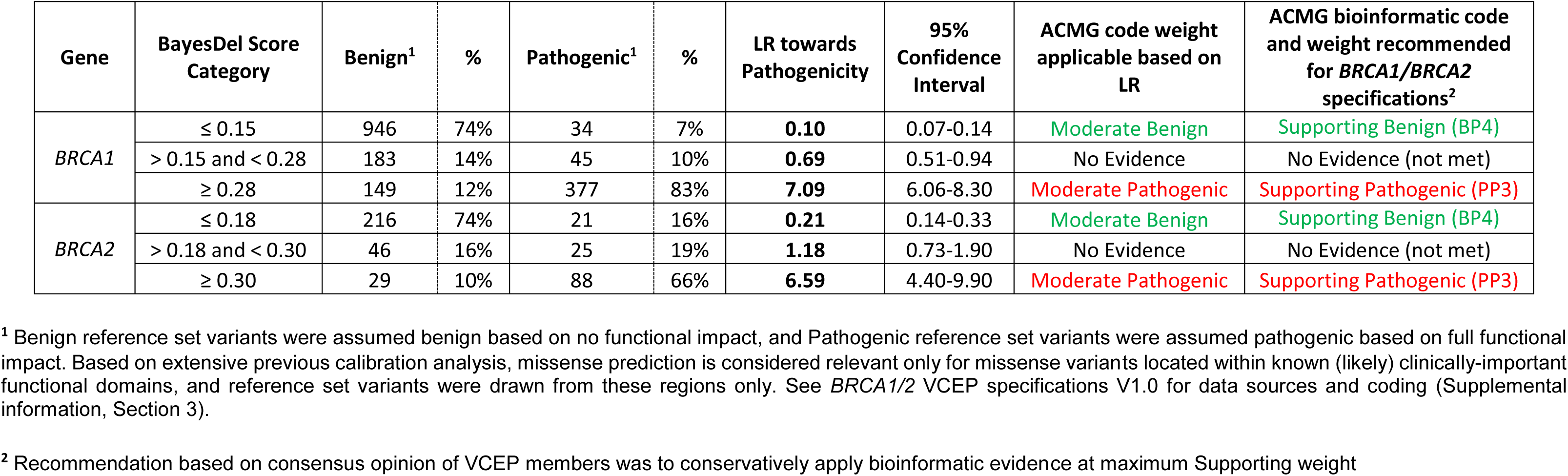
LR towards pathogenicity for BayesDel categories selected for *BRCA1* and *BRCA2* VCEP specifications.

| Gene | BayesDel Score Category | Benign<sup>1</sup> | % | Pathogenic<sup>1</sup> | % | LR towards Pathogenicity | 95% Confidence Interval | ACMG code weight applicable based on LR | ACMG bioinformatic code and weight recommended for *BRCA1/BRCA2* specifications<sup>2</sup> |
| --- | --- | --- | --- | --- | --- | --- | --- | --- | --- |
| *BRCA1* | ≤ 0.15 | 946 | 74% | 34 | 7% | **0.10** | 0.07-0.14 | Moderate Benign | Supporting Benign (BP4) |
|  | > 0.15 and < 0.28 | 183 | 14% | 45 | 10% | **0.69** | 0.51-0.94 | No Evidence | No Evidence (not met) |
|  | ≥ 0.28 | 149 | 12% | 377 | 83% | **7.09** | 6.06-8.30 | Moderate Pathogenic | Supporting Pathogenic (PP3) |
| *BRCA2* | ≤ 0.18 | 216 | 74% | 21 | 16% | **0.21** | 0.14-0.33 | Moderate Benign | Supporting Benign (BP4) |
|  | > 0.18 and < 0.30 | 46 | 16% | 25 | 19% | **1.18** | 0.73-1.90 | No Evidence | No Evidence (not met) |
|  | ≥ 0.30 | 29 | 10% | 88 | 66% | **6.59** | 4.40-9.90 | Moderate Pathogenic | Supporting Pathogenic (PP3) |

<sup>1</sup> Benign reference set variants were assumed benign based on no functional impact, and Pathogenic reference set variants were assumed pathogenic based on full functional impact. Based on extensive previous calibration analysis, missense prediction is considered relevant only for missense variants located within known (likely) clinically-important functional domains, and reference set variants were drawn from these regions only. See *BRCA1/2* VCEP specifications V1.0 for data sources and coding (Supplemental information, Section 3).
<sup>2</sup> Recommendation based on consensus opinion of VCEP members was to conservatively apply bioinformatic evidence at maximum Supporting weight

Specifically, use of the Pejaver et al. scale performed very poorly for benign reference set variants, in that a BP4 code (at minimum supporting strength for BayesDel ≤-0.18) would be assigned to < 10% of *BRCA1* and *BRCA2* benign reference set variants, with the large majority having no code applicable. Further, a considerable proportion (29% for *BRCA1*, 36% for *BRCA2*) would be incorrectly assigned PP3 at minimum supporting evidence strength, for BayesDel ≥ 0.13.

Specification of the frequency codes PM2/BS1/BA1 was informed by a combination of LR-based methods^17^, and minimal credible allele frequency estimation^18^ as recommended by ClinGen. LR estimation approaches previously used for weighting combined results from functional assays^12^ were repeated using an expanded dataset, and confirmed applicability of PS3 and BS3 at Strong level. Extensive review of the literature and consideration of FA-designated features in GeneReviews (https://www.ncbi.nlm.nih.gov/books/NBK1401/) informed the use of presence or (apparent) absence of FA phenotype for application of codes PM3 and BS2, respectively. Recommendations for use and weighting of segregation data for codes PP1 and BS4 built on methods previously established and enhanced for *BRCA1* and *BRCA2* variant interpretation by the ENIGMA consortium,^8; 19^ which consider gene-specific age-specific cumulative risk (penetrance) and background population incidence in assessing variant causality.

The application (i.e. “criterion” description) was completely re-purposed for four codes in consultation with the ClinGen SVI WG, after consideration of empirical data on *BRCA1/2*-related clinical features. PM5 was designated to assign exon-specific weights for a premature termination codon (PTC) variant found in an exon in which functional data and/or case-control burden analysis and/or family history burden analysis proves that PTCs in the exon are indeed pathogenic (as justified by VCEP analysis, Figure 3). BP1 was used to capture strong evidence against pathogenicity for a variant outside of a known clinically important functional domain predicted to encode a silent or missense/in-frame substitution only (without known or predicted impact on splicing), with strength assigned from probability based studies^17^, and VCEP consideration of large-scale case-control findings for missense variants.^20; 21^ PP4 and BP5 were repurposed to capture combined LR estimates towards pathogenicity (PP4) or against pathogenicity (BP5), as derived from calibrated multifactorial likelihood ratio analysis (but excluding any direct statistical measurement of bioinformatic prediction scores, to avoid overlap with other computational codes).

**Figure 3:**
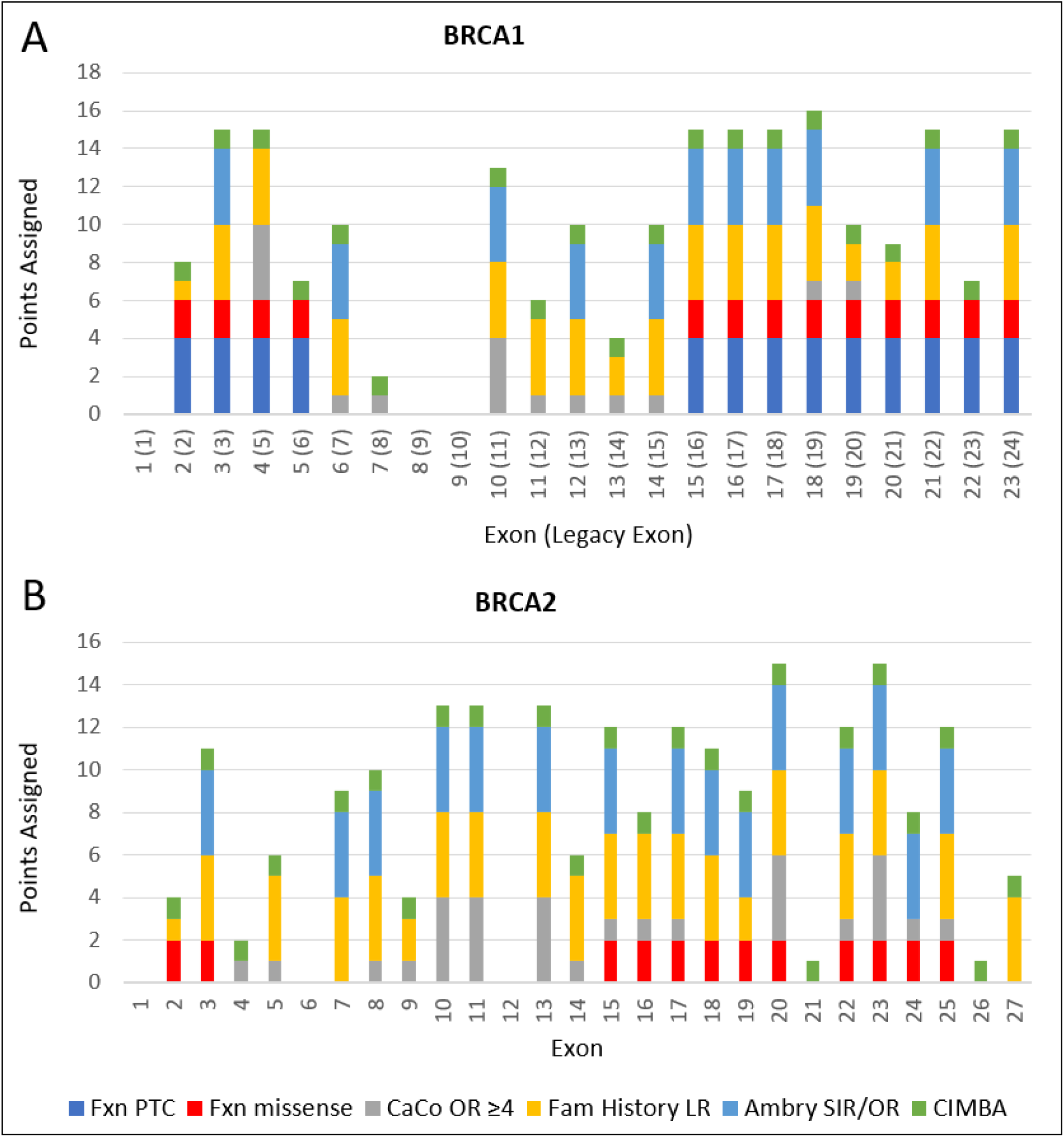
Overview of evidence supporting PM5 exon-specific weights application for premature termination codon variants in *BRCA1 (A) and BRCA2 (B)* Exon-specific points assigned are derived from per-exon evidence, as follows: observed experimental impact on function for at least one premature termination codon (PTC) variant in an exon, to infer code PS3 for another PTC variant in the same exon (Fxn PTC, assigned 4 points); observed experimental impact on function for at least one missense substitution variant in the exon indicates no rescue of pathogenic variants due to alternative splicing, and that a PTC variant leading to nonsense mediated decay would have at minimum the same impact - used to infer code PS3_moderate for another PTC variant in the same exon (Fxn Missense, assigned 2 points); Case-control Odds Ratio (OR) ≥4.0 estimated for PTC variants observed in a given exon, to infer code PS4 for any PTC variant in that exon (CaCo OR ≥4), assigned at full strength for a statistically significant association (4 points), and at supporting strength for non-significant estimates (1 point); family history likelihood ratio (LR) estimates from heterogeneity analysis, using personal and family history profile as a predictor of pathogenicity in a clinical dataset^29^ (Fam History LR, assigned points based on LR); standardized incidence ratio (SIR) ≥4.0, for PTC variants in non-Finnish European (NFE) probands with breast, ovarian and/or pancreatic cancer in a clinical dataset sourced from Ambry Genetics^29^ versus the summed frequency of PTC in gnomAD NFE (Ambry SIR/OR), with SIR ≥ 4, P<0.05 was used to infer code PS4 (4 points), and SIR ≥4 non-significant as PS4_supporting (1 point); identification of PTC variants in a given exon in multiple independent probands in the CIMBA (https://cimba.ccge.medschl.cam.ac.uk/) highly ascertained cohort of *BRCA1* and *BRCA2* pathogenic variant carriers (including patients who historically met strict clinical criteria to undergo diagnostic testing), with observation of ≥5 unique PTCs variant in ≥ 5 CIMBA families used to infer PP4 (CIMBA, 1 point). The per-exon evidence was summed across the different evidence types (Fxn PTC, Fxn Missense, CaCo OR ≥4, Fam History LR, Ambry SIR/OR, CIMBA), to derive an exon-specific evidence strength using a points-based approach (Supporting = 1 point, Moderate = 2 points, Strong = 4 points). Based on the combined evidence, the PM5 (PTC) code can be applied as Strong evidence in favour of pathogenicity for most exons. The PM5 (PTC) code can only be applied to germline variants that meet PVS1 codes, namely nonsense and frameshift changes, including large deletions and tandem duplications. Code weight is determined by the exon in which the predicted termination codon occurs. For example, a frameshift variant in BRCA2 exon 15 that is predicted to result in a PTC within exon 16 would use the code strength of BRCA2 exon 16 (PM5_Strong (PTC)). Variants in the AG-GT splice site positions do not qualify for PM5 (PTC) code.

### Key considerations during development of the *BRCA1* and *BRCA2* specifications

Major points for discussion with key ClinGen SVI WG members before and during documentation of the draft specifications included codes capturing bioinformatic or experimental impact on mRNA splicing; these were later resolved in part by review conducted under the umbrella of the ClinGen SVI Splicing Subgroup. Extensive discussion of additional codes, which at the time were new requests for code adaptations by a VCEP, included:

● downgrading PM2 (absence in population databases) to PM2_supporting, informed by parallel research^17^
● adapting the PVS1 decision tree recommendations for weighting predicted loss of function variants^22^ to consider

♦ the importance of naturally occurring rescue isoforms^23^
♦ functional domains designated as clinically important based on location of known pathogenic missense variants (as per ClinGen-approved external expert panel criteria for *BRCA1* and *BRCA2* (Supplemental Information, Section 1)
♦ duplications that preserve reading frame
♦ splice donor/acceptor ±1,2 dinucleotide variants for exons outside of the coding exons (5’ or 3’ UTRs)
♦ splice donor ±1,2 dinucleotide variants that create *de no*v*o* predicted functional “GC” 5’ splice sites
● repurposing PM5 to capture exon-specific evidence as additional information for classification of predicted loss of function stop and frameshift variants, motivated by existing clinical evidence that PTC variants are highly likely to be pathogenic

♦ variants in the AG-GT splice site positions do not qualify for PM5 (PTC) code, since the mechanism of impact on mRNA transcripts may introduce variability in proportion of loss of function transcripts produced
● considering that protein domain combined with missense bioinformatic predictions, and clinical data from published case-control data, achieves greater predictive value than bioinformatic missense prediction alone, and acceptance that an upweighted repurposed BP1_Strong code for missense and synonymous variants outside of a known (likely) clinically important protein domain is sufficient to achieve likely benign classification for *BRCA1* and *BRCA2* variants
● repurposing PP4 and BP5 to capture likelihood data for multiple evidence types calibrated to predict pathogenicity

♦ expanding potential to provide evidence against pathogenicity for data types previously only considered as positive predictors of pathogenicity e.g. case-control OR can be applied as PS4 only, but case-control LR estimates could be applied as PP4 or BP5
♦ increasing the breadth of information types that might be used for variant interpretation, even if not explicitly or directly captured by existing ACMG/AMP criteria e.g. breast tumor pathology features predictive of *BRCA1* or *BRCA2* variant pathogenicity are not specific to individuals with a pathogenic variant in these genes
♦ allowing combined likelihoods to be captured under a single code
♦ facilitating alignment with pre-existing variant classifications based on multifactorial likelihood analysis

There was also resolution to align code combinations to achieve variant classes following recommendations arising from Bayesian modelling of the ACMG/AMP classification system^13^, and agreement to use the points approach^24^ to justify expansion of benign code combinations to include benign codes at moderate strength level, and use of the points approach to resolve classifications for variants with discordant benign and pathogenic code application.

### Pilot application of specifications for *BRCA1* and *BRCA2* classification

An overview of the classifications during and after the pilot curation process is shown in Figure 4. Classifications assigned to the 40 pilot variants at the first and second curation phases, and the final classification assigned (with codes applied) are detailed in Table S1. Pre-existing ClinVar classification for the pilot variants, based on all submitter variant assertions at the time of extraction, was as follows: 13 VUS/conflicting, 11 P, 3 LP/P, 1 LB/B, and 12 B. After initial review, 32/40 variants achieved classification within a confidence band (LP/P or LB/B). Review of the classifications identified several reasons for the differences: new pieces of unpublished internal information used by one biocurator only; unfamiliarity with data presentation; need for clarification of code use. Between-curator differences often involved: recoding of published multifactorial likelihood data to ACMG/AMP codes PP4 and BP5 (23/40); use of frequency information (16/40); use of bioinformatic data (17/40); weighting of splicing data and use of functional data (15/40). Biocurator feedback indicated the need for more specific advice for some codes, simplified tables and figures in a single “specifications” document, and more detailed recommendations for interpretation of mRNA splicing data. Documentation was revised accordingly, including development and inclusion of an RNA rubric for weighting of mRNA assay data. At this point, additional calibration analysis was undertaken to reassess BayesDel score cut points (as per Table 1), and results incorporated into the revised specifications.

**Figure 4:**
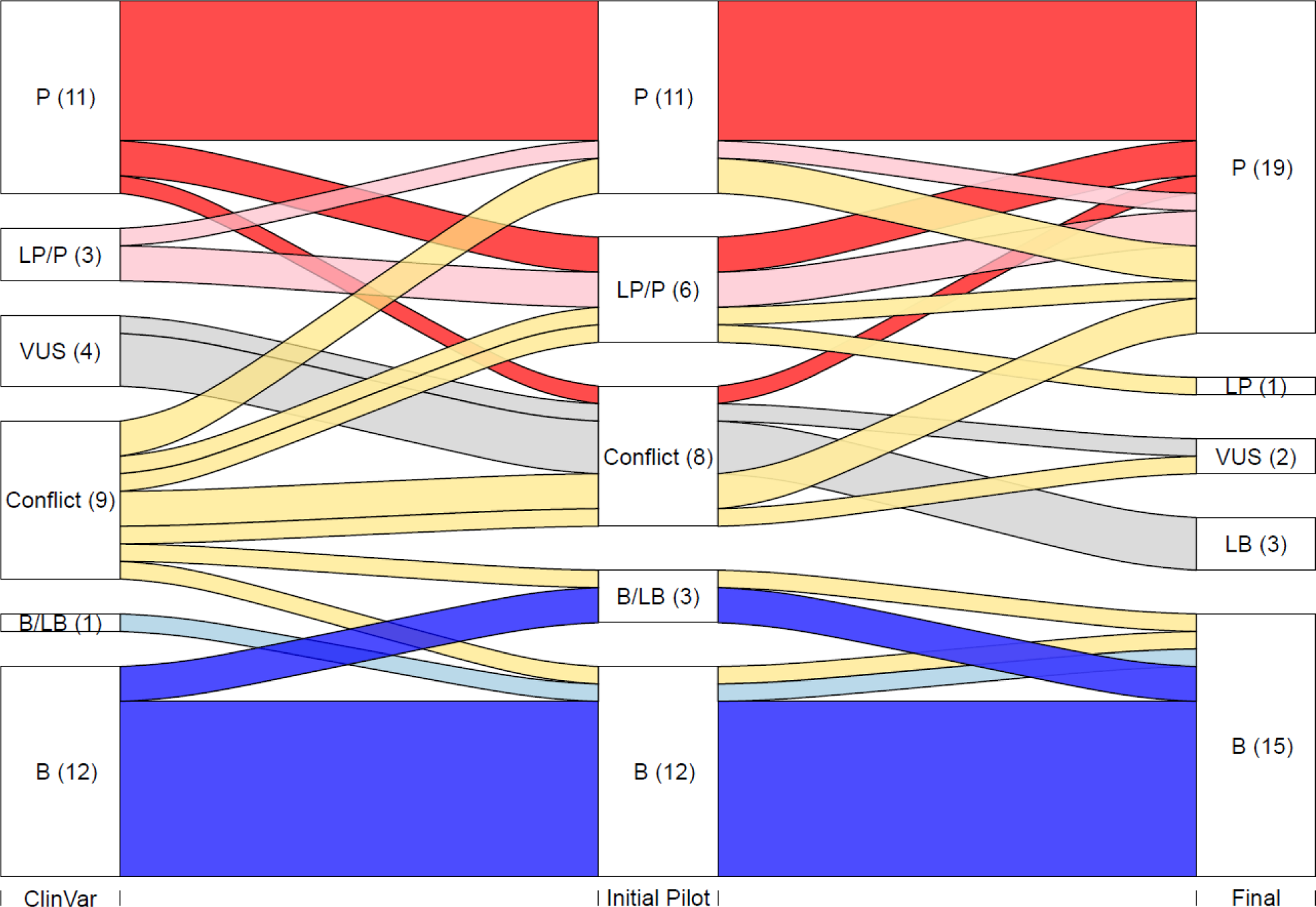
Overview of variant classifications assigned during pilot of VCEP specifications. Sankey diagram shows transition in classification categories for pilot variants, from initial ClinVar classification to final classification assigned by the VCEP. The left column represents the initial ClinVar classification category/grouping of pilot variants. The central column represents classification/s by biocurators after the initial pilot phase. The right column represents the final classifications of pilot variants. Classification categories: P=Pathogenic; LP=Likely Pathogenic; VUS=Variant of uncertain significance; Conflict=conflicting; LB=Likely Benign; B=Benign. For each category/grouping, the number of variants is shown in parentheses. Final VCEP curation resulted in increased certainty in classification for all variants with initial LP/P or B/LB category, movement of three of four variants with initial VUS classification outside of this category, and resolution in classification for the nine variants with initial conflicting classification - eight reaching classification other than VUS.

After re-review in the second curation phase, six of eight variants assigned to “VUS/other” group in the first phase were resolved to a single classification. For variants with classifications that differed in confidence, 5/6 LP/P and all 3 LB/B resolved to a more certain classification (i.e. P or B). Compared to the original ClinVar class, classification was resolved for 11/13 VUS/conflicting variants (5 P, 1 LP, 2 VUS, 3 LB, 2 B). All variants with pre-existing ClinVar class P (11 variants) or B (12 variants) retained class. Of the remainder, 3 LP/P variants were upgraded to P, and a single LB/B variant was classified as B. As expected, variants annotated with missense or intronic molecular consequences showed greater classification uncertainty and variability (considering conflicts and confidence differences) compared to premature termination codon and synonymous variants, in both initial ClinVar classification and at the first VCEP curation step (see Table S1 for details).

Further minor revisions of the specifications and appendices were introduced following biocurator feedback, and after final review from the ClinGen SVI WG.

### Conclusions and future directions

Alignment of pre-existing *BRCA1* and *BRCA2* ENIGMA classification processes with ACMG/AMP classification criteria highlighted several gaps in both the pre-existing processes and in the baseline ACMG/AMP criteria. Statistical calibration of different evidence types was key to justify acceptance – or rejection - of the utility of different ACMG/AMP evidence codes for classification by VCEP members, and also the ClinGen SVI WG overseeing VCEP approval. Functional evidence was lacking from the pre-existing ENIGMA external panel criteria for *BRCA1* and *BRCA2* (Supplemental Information, Section 1), and the requirement to align with ACMG/AMP processes provided motivation and mechanism for the VCEP to define suitable data sources and reach consensus on specifications for this evidence type. Regarding the codes/criteria deemed not applicable, VCEP member individual opinion concerning the utility of proband-counting criterion was sufficiently contentious that a separate sub study was conducted. This study demonstrated that proband counting with comparison to population datasets is not sufficiently robust for generic application for *BRCA1* and *BRCA2*, given that these genes lead to relatively common diseases.^16^ Major items for discussions with key members of the ClinGen SVI WG revolved around the need for ACMG/AMP criteria to be adapted or repurposed to capture more evidence types (and strengths), in particular to provide evidence against pathogenicity. Agreement by the ClinGen SVI WG to adapt an existing code PM5 provided a mechanism for additional exon-specific weighting so that pre-existing diagnostic laboratory classification practices for *BRCA1* and *BRCA2* PTC variants would not be reversed on introduction of ACMG/AMP classification system (unless indicated by evidence in this process).

The alignment of pre-existing ENIGMA classification methods with ACMG/AMP processes has led to benefits beyond interpretation of variants in *BRCA1* and *BRCA2*. The research-driven consideration of evidence types and calibration by the ENIGMA *BRCA1* and *BRCA2* VCEP informed the activities of the SVI Splicing Subgroup, and has already led to uptake of some of the “*BRCA1/BRCA2”* specifications for ACMG/AMP criteria by other ClinGen VCEPs. These have included: introduction of bioinformatic tiers for splicing prediction; consideration of RNA data under the PVS1 decision process; consideration of read depth for annotation of frequency codes;^25; 26^ alignment of weights (for recessive disease) with *PALB2*, another gene associated with Fanconi Anemia; uptake of the PM5 code use for PTC variants in multiple other genes (including *ATM*, *CDH1*, *PALB2*, and *RUNX1*). It is also notable that some of these adaptations have been taken forward for the draft iteration of the next version of the ClinGen-promoted classification guidelines for application to any Mendelian gene.

The ClinGen ENIGMA *BRCA1* and *BRCA2* VCEP has now initiated ACMG/AMP-aligned review of *BRCA1* and *BRCA2* variants in ClinVar. As a priority the VCEP is reviewing variants with conflicting assertions and will shortly re-assess pre-existing external expert panel curations to highlight any variants expected to alter in classification after application of the VCEP specifications. VCEP review of all *BRCA1* and *BRCA2* variants in ClinVar will be an extensive and time-consuming effort, both due to the enormity of the task (11,932 *BRCA1* and 17,109 *BRCA2* variants as at 1^st^ November 2023) and constraints associated with the rigorous FDA-aligned ClinGen protocol. For example, after biocurator assessment of information and classification by code assignment and manual entry into the ClinGen Variant Curation Interface,^27^ three core approvers are required to review and agree with the classification. The VCEP aims to introduce additional efficiencies to ease the load of biocurators in variant review, such as algorithmic code assignment based on frequency and computational information in the BRCA Exchange portal (https://brcaexchange.org/)^28^, as a means to prioritize variants for additional data collection and review. This portal will also provide a mechanism for public dissemination of VCEP-aligned ACMG/AMP codes for *BRCA1* and *BRCA2* variants ahead of formal VCEP review.

In summary, this work has provided extensive evidence-based specifications to enable standardized and improved classification of variants in *BRCA1* and *BRCA2*. Further, it has more widely informed improvements in both gene-specific and generic application of the ACMG/AMP classification guidelines.

## Supporting information

Supplemental Section 2 Table 1

Supplemental Section 2 Table 2

Supplemental Section 2

Supplemental Section 1

Supplemental Section 3

## Data Availability

All data produced in the present work are contained in the manuscript

## Acknowledgments

Refer to Supplemental Information Section 2.

## Author contributions

All authors contributed to conceptualization and writing (review and editing). Funding acquisition by F.J.C. and A.B.S. Data curation by M.T.P., E.T., L.C.W., M.d.l.H., and A.B.S. Formal analysis conducted by D.E.G., M.T.P, B-J.F., G.A.R.W, L.C.W., M.d.l.H., V.N., M.E.R, and A.B.S. Investigation by M.T.P., W.B-F., R.C.C., M.d.l.H., S.H., M.H., H.S.L., A.R.M., R.O., M.E.R., G.S., I.S.P., B.A.T., M.P.G.V., L.Z., C.F. and J.H. All authors read and approved the final manuscript.

## Web resources

URLs are provided in the text where appropriate.

## Data and code availability

The published article includes all datasets generated or analyzed during this study.

