## Supplemental Section 2 for "Evidence-based recommendations for gene-specific ACMG/AMP variant classification from the ClinGen ENIGMA BRCA1 and BRCA2 Variant Curation Expert Panel"

**Supplemental Information Section 2: Description of supplemental tables and additional bioinformatic calibration analysis**

### **Acknowledgements**

We acknowledge Susan Domchek, Barbara Wappenschmidt and David Tran for their contributions to ENIGMA BRCA1 and BRCA2 expert panel activities. The work of M.T.P. was supported in part by National Institutes of Health grant U24 5U24CA258058-02. M.C. was supported by the Basser Center for BRCA Research and the National Cancer Institute (U24CA258058). F.J.C. was supported by NIH grants U24CA258058, R35CA253187 and P50CA116201 and the Breast Cancer Research Foundation. M.d.l.H. was supported by a grant from the Spanish Ministry of Science and Innovation, Plan Nacional de I+D+I 2013-2016, ISCIII (PI20/00110) co-funded by FEDER from Regional Development European Funds (European Union) and by NIH grant 5U24CA258058-02. M.H. was supported by NIH grant 5U24CA258058. A.N.M. is a Breast Cancer Research Foundation investigator. P.R. was supported by a grant of the Italian Association for Cancer Research (AIRC - IG 22093). M.S. was supported by an Australian National Health and Medical Research Council L3 Investigator Grant (GNT2017325). M.P.G.V was supported by The Dutch Cancer Society grant KWF 12754. L.C.W. and G.A.R.W. were supported by the New Zealand Health Research Council (22/187). Cristina Fortuno was supported in part by NHMRC project funding (APP1161589), and a grant from the National Breast Cancer Foundation, Australia (IIRS-21-102). ABS was supported by an NHMRC Investigator Fellowship (APP177524). U24CA258058 for in-part support of VCEP activities from 2023. C.T. acknowledges funding from a Cancer Research UK catalyst award (C61296/A27223). This publication was supported in part by the National Institute of General Medical Sciences R01 GM134731 and Department of Defense W81XWH1810269. The content is solely the responsibility of the author and does not necessarily represent the official views of the National Institutes of Health.

### **Supplemental Tables provided as separate excel files**

#### **Table S1**: Pilot variants assessed using ACMG/AMP criteria specified for *BRCA1* and *BRCA2*

#### **Table S2:** General description of *BRCA1* and *BRCA2* VCEP specifications for ACMG/AMP baseline criteria and evidence codes

Note, specifications are expected to be updated over time; the most up-to-date specifications are available via the ClinGen C-spec registry (<https://cspec.genome.network/cspec/ui/svi/affiliation/50087>).

### **BayesDel recalibration for the ClinGen ENIGMA *BRCA1* and *BRCA2* Variant Curation Expert Panel specifications**

Initial analyses used to select the optimal bioinformatic prediction tool/score for missense impact prediction followed a multi-pronged approach. See Appendices for *BRCA1* and *BRCA2* VCEP Specifications Version 1.0, provided in Supplemental Information, Section 3; also available via <https://cspec.genome.network/cspec/ui/svi/affiliation/50087>. Results indicated that location of a missense variant outside of a (potentially) clinically important functional domain provided strong evidence against pathogenicity, irrespective of bioinformatics prediction of missense impact using several different tools. Functional assay results for BRCA1 and BRCA2 variants were used to define assumed Benign (no functional impact) and assumed Pathogenic (functional impact) reference set variants (total 1608 variants), and based on calibration results from this analysis, BayesDel (binary score cutoffs <0.3, ≥0.30) was selected for application of BP4 and PP3.

During the course of the pilot phase, Receiver Operating Curve (ROC) analysis was conducted using an expanded dataset comprising functional assay results for 2159 missense variants (1734 BRCA1, 425 BRCA2) that all map to (potentially) clinically important functional protein domains, previously defined in VCEP documentation as follows: BRCA1 RING aa (amino acid) 2-101; BRCA1 coiled-coil aa 1391-1424; BRCA1 BRCT repeats aa 1650-1857; BRCA2 PALB2 binding domain aa 10-40; and BRCA2 DNA binding aa 2481-3186. Further, for this analysis, genes were considered separately. As for the initial calibration analysis, variants with full impact on function were designated as an assumed pathogenic reference set, and those with no functional impact as an assumed benign reference set. The optimal binary cutpoint differed somewhat between the two genes (Figure S1). Based on these findings, further calibration analysis was done separately for *BRCA1* and *BRCA2*. LR analysis was conducted to select optimal score ranges for three categories (evidence towards pathogenicity, against pathogenicity, and no evidence as the middle score range category). Results are shown in Table 1. Optimal score ranges differed slightly for the two genes.

Alongside the work of this *BRCA1/BRCA2* VCEP, a Subgroup of the ClinGen Sequence Variant Interpretation Working Group conducted extensive analysis to compare the utility of different bioinformatic tools for generic use in missense pathogenicity prediction, under the codes PP3 and BP4 (Pejaver et al., PMID: 36413997). The optimal BayesDel score ranges now recommended for generic use by ClinGen differ from those derived from our calibration analysis focused on BRCA1 and BRCA2 missense variant prediction. We thus conducted LR analysis using our reference datasets of 1734 BRCA1 and 425 BRCA2 missense variants to assess the performance of the generic score thresholds for *BRCA1* and *BRCA2*. The results, shown in Table S3, indicate that the weights recommended for generic analyses are inappropriate for both BRCA1 and BRCA2 missense variants. The No Evidence category recommended for generic use by Pejaver et al. would be applied to the majority of missense variants in the BRCA1 and BRCA2 benign reference sets (68% BRCA1, 54% BRCA2), variants for which the VCEP gene-specific calibration yields an LR that could be assigned a Moderate Benign weight. Moreover, ~10% of BRCA1 missense variants, and ~20% of BRCA2 missense variants in the benign and pathogenic reference sets would be assigned supporting Pathogenic using the Pejaver et al. scale, instead of no evidence as per the VCEP gene-specific calibration. Overall, use of the Pejaver et al. scale would amount to misleading bioinformatic category being assigned for 78-85% of the benign reference set variants, and 17-30% of the pathogenic reference set variants.

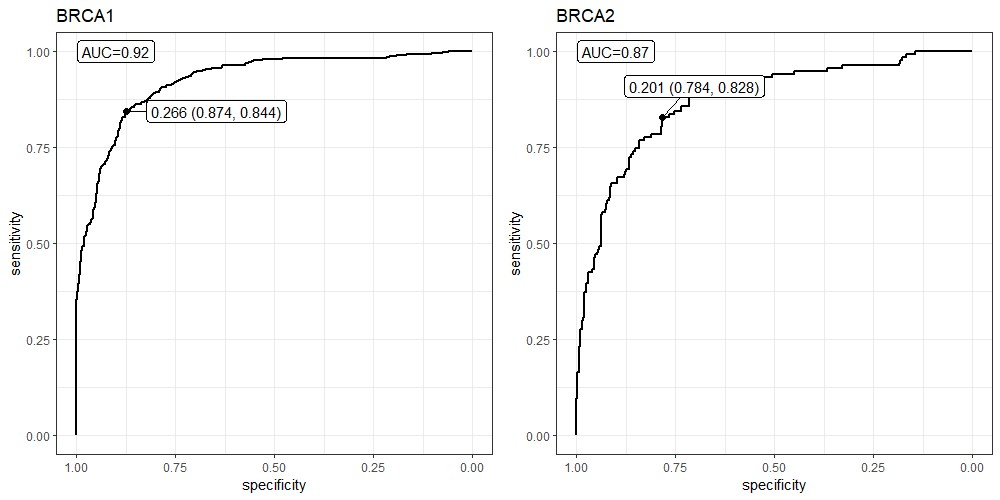

#### **Figure S1:** ROC analysis to define optimal BayesDel binary cutpoint for expanded reference sets of missense variants.

Analysis included information for 1745 BRCA1 and 320 BRCA2 missense variants. The optimal binary cutpoint was 0.266 (sensitivity 0.874, specificity 0.844) for BRCA1 and 0.201 (sensitivity 0.784, specificity 0.828) for BRCA2.

#### **Table S3:** Estimated LR towards pathogenicity for BRCA1 and BRCA2 missense variants using BayesDel score categories recommended for generic use by the ClinGen Sequence Variant Interpretation Working Group.

|  | **Code weight recommended from Pejaver et al.^1^** | **Score Category**  **Pejaver et al. ^1^** | **Benign** | | **Pathogenic** | | **LR towards Pathogenicity** | **95% Confidence Interval** | **Weights for similar score ranges based on LR estimates from BRCA1 and BRCA2 dataset analysis^2^** |
| --- | --- | --- | --- | --- | --- | --- | --- | --- | --- |
|  |  |  | **n** | **%** | **n** | **%** |  |  |  |
| **BRCA1** | Moderate Benign | ≤ -0.36 | 4 | 0.35% | 0 | 0.11% | 0.311 | (0.016-5.87) | Moderate Benign for BayesDel score ≤ 0.15 |
|  | Supporting Benign | ≤ -0.18 | 41 | 3.24% | 0 | 0.11% | 0.034 | (0.002-0.547) |  |
|  | **N/A** | >-0.18  < 0.13 | 853 | 66.74% | 25 | 5.48% | 0.082 | (0.056-0.12) |  |
|  | Supporting Pathogenic | ≥ 0.13 | 221 | 17.29% | 48 | 10.53% | 0.609 | (0.454-0.82) | No evidence |
|  | Moderate Pathogenic | ≥ 0.27 | 159 | 12.44% | 226 | 49.56% | 3.984 | (3.35-4.73) | Moderate Pathogenic for BayesDel score ≥ 0.28 |
|  | Strong Pathogenic | ≥ 0.50 | 0 | 0.04% | 157 | 34.46% | 881.59 | (55.32-14049) |  |
|  |  | **Total** | **1278** |  | **456** |  |  |  |  |
| **BRCA2** | Moderate Benign | ≤ -0.36 | 4 | 1.54% | 0 | 0.37% | 0.240 | (0.013-4.51) | Moderate Benign for BayesDel score ≤ 0.18 |
|  | Supporting Benign | ≤ -0.18 | 25 | 8.73% | 0 | 0.37% | 0.042 | (0.003-0.692) |  |
|  | **N/A** | >-0.18  < 0.13 | 157 | 53.95% | 12 | 8.96% | 0.166 | (0.096-0.288) |  |
|  | Supporting Pathogenic | ≥ 0.13 | 69 | 23.71% | 30 | 22.39% | 0.944 | (0.648-1.376) | No evidence |
|  | Moderate Pathogenic | ≥ 0.27 | 36 | 12.37% | 80 | 59.70% | 4.826 | (3.45-6.75) | Moderate Pathogenic for BayesDel score ≥ 0.30 |
|  | Strong Pathogenic | ≥ 0.50 | 0 | 0.17% | 12 | 9.26% | 54.07 | (3.21-908) |  |
|  |  | **Total** | **291** |  | **134** |  |  |  |  |

^1^ Pejaver, V. *et al.* Calibration of computational tools for missense variant pathogenicity classification and ClinGen recommendations for PP3/BP4 criteria. *Am J Hum Genet* **109**, 2163-2177 (2022). PMID: 36413997.

^2^ As shown in Table 1 in the main paper.
