## Supplemental Section 1 for "Evidence-based recommendations for gene-specific ACMG/AMP variant classification from the ClinGen ENIGMA BRCA1 and BRCA2 Variant Curation Expert Panel"

### **Supplemental Information Section 1:** Pre-existing ENIGMA external expert panel classification criteria for *BRCA1* and *BRCA2*.

Please see following pages for the classification criteria developed by the ENIGMA *BRCA1/2* external expert panel (Version 2.5.1 29 June 2017), and applied to variants for display in ClinVar up to Feb 2020.

**These external expert panel criteria are NO LONGER IN ACTIVE USE.**

**They have been superseded by the specifications developed by the ClinGen ENIGMA *BRCA1* and *BRCA2* Variant Curation Expert Panel (initial approved version provided in Supplemental Information Section 3, most up-to-date version available via https://cspec.genome.network/cspec/ui/svi/affiliation/50087).**

### ENIGMA *BRCA1/2* Gene Variant Classification Criteria

ENIGMA (Evidence-based Network for the Interpretation of Germline Mutant Alleles) is an international consortium of investigators focused on determining the clinical significance of sequence variants in breast cancer genes. Information about the consortium purpose, membership criteria and operation can be found at http://www.enigmaconsortium.org/.

#### Rules for Variant Classification:

Rules describing the 5 class system for classification of *BRCA1/2* gene variants were initially devised and documented by ENIGMA Steering committee members, and revised with input from ENIGMA collaborators. These ENIGMA criteria provide a baseline for standardized clinical classification of *BRCA1/2* gene sequence variation that may be linked to patient and family management in the genetic counseling arena according to published guidelines (Plon et al., 2008). At the present time, the proposed classification criteria are intended to differentiate germline high risk variants (associated with cancer risk equivalent to classical pathogenic variants that are known or predicted encode a premature termination codon i.e. nonsense or frameshift) from variants with low or no risk. These guidelines are not intended for the evaluation and classification of variants associated with an intermediate or moderate level of risk e.g. *BRCA1* c.5096G>A p.Arg1699Gln (Spurdle et al., 2012). It should also be noted that, based on observations in mouse model and *ex-vivo* analyses (Drost et al., 2011, Meeks et al., 2016, Drost et al., 2016), it is currently unclear what the correlation is between germline variant classification for cancer risk and germline/somatic variant classification for response to therapy eg with PARP inhibitors.

The ENIGMA criteria are based on a combination of the following:

- The 5 class system described for quantitative assessment of variant pathogenicity in Plon et al.(2008) using a multifactorial likelihood model(Goldgar et al., 2004, Easton et al., 2007, Goldgar et al., 2008, Tavtigian et al., 2008) (see Appendix, Table 1);
- The 5 class system for interpretation of possible spliceogenic variants and splicing alterations developed by ENIGMA collaborators (Walker et al., 2013);
- Generic elements of the 5 class quantitative/qualitative scheme for mismatch repair gene variant classification developed by InSiGHT (Thompson et al., 2014);
- Generic elements of the ACMG guidelines for interpretation of sequence variations (Richards et al., 2008);
- Classification criteria developed by individual sites participating in ENIGMA, including established country networks;
- The classification of sequence changes according to common clinical practice – that is, description of variants generally considered pathogenic (clinically relevant in a genetic counseling setting such that germline variant status is used to inform patient and family management) or non-pathogenic (significant evidence against being a dominant high-risk pathogenic variant).

Appendix, Table 2 summarises the rationale supporting the criteria.

The interpretation of variant clinical significance in relation to functional domains is assisted by definition of clinically important functional domains and residues (Appendix, Tables 3 and 4).

An overview of the ENIGMA classification process is shown in Appendix, Figure 1.

NO LONGER IN USE

Use of the ENIGMA variant interpretation guidelines is subject to user discretion and responsibility. Guidelines are subject to change with availability of new information and interpretation processes, and we thus recommend date-stamping for all variant classifications. Interpretation of variants using these criteria does not exclude the very low probability that there is a pathogenic variant *in cis* undetected by the testing protocol, which may confound interpretation of variant pathogenicity.

**For a given class, a bullet point "●" represents an "OR" statement, whereas the symbol "✓" represents an "AND" statement. That is, a variant is required to satisfy all the criteria listed for at least one bullet-point that falls within that class.**

NO LONGER IN USE

**Class 5 – Pathogenic**

*There is significant evidence to suggest that this variant is a dominant high-risk pathogenic variant and no substantial conflicting evidence.*

- Variant with probability of pathogenicity >0.99 using a multifactorial likelihood model[1].
- Coding sequence variant that encodes a premature termination codon i.e. nonsense or frameshift alteration predicted to disrupt expression of known clinically important functional protein domain(s)/residues[2] and for which there is low bioinformatic likelihood[3] to disrupt normal splicing (e.g. disruption of native donor/acceptor sites, creation of *de novo* donor). *Note: Predicted nonsense or frameshift variants with high bioinformatic likelihood[3] to disrupt normal splicing require mRNA assays to assess the nature and possible clinical consequence of aberrant transcripts and/or clinical studies to confirm pathogenicity.*
- Variant allele tested for mRNA aberrations using *in vitro* assays of patient RNA[4] that assesses allele-specific transcript expression, and is found to produce only transcript(s) carrying a premature termination codon, <u>or</u> an in-frame deletion disrupting expression of one or more known clinically important residues[2].
- Copy number deletion variant that removes one or more exons spanning a known clinically important functional [2] or is proven by laboratory studies to result in a frameshift alteration predicted to disrupt expression of one or more known clinically important functional residues[2].
- Copy number duplication variant of any size that duplicates one or more exons and is proven by laboratory studies to result in a frameshift alteration predicted to disrupt expression of one or more known clinically important residues[2].

NO LONGER IN USE

**Class 4 – Likely pathogenic**

*There is evidence that this variant is a dominant high-risk pathogenic variant and no substantial conflicting evidence.*

- Variant with probability of pathogenicity between 0.95-0.99 using a multifactorial likelihood model

- In the absence of clinical evidence to assign an alternative classification, a sequence variant that alters the *BRCA1* or *BRCA2* translation initiation methionine site.

- Variant considered extremely likely to alter splicing based on position, namely IVS±1 or IVS±2, or G>non-G at last base of exon if first 6 bases of the intron are <u>not</u> GTRRGT *(listed in Table 5)*, and
  - ✓ is untested for splicing aberrations using *in vitro* assays of patient RNA that assesses **allele-specific** transcript expression
  - ✓ is predicted bioinformatically to alter use of the native donor/acceptor site
  - ✓ is not predicted or known to alter production of (naturally occurring) in-frame RNA isoforms that may rescue *3 (uncertain) unless proven otherwise).*

- A variant demonstrating all these features:
  - ✓ encodes the same amino acid change as a previously established Class 5 pathogenic **missense** variant with a different underlying nucleotide change[2]
  - ✓ no evidence of mRNA aberration (splicing or expression) from *in vitro* mRNA assays[4].
  - ✓ the variant is absent from outbred control reference groups[5]

- A small in-frame deletion variant demonstrating these features:
  - ✓ removes codon for which a missense substitution Class-5 variant has been described[2]
  - ✓ low bioinformatic likelihood to disrupt normal splicing[3]
  - ✓ the variant is absent from outbred control reference groups[5]

NO LONGER IN USE

**Class 3 – Uncertain**

*There is insufficient evidence to place this variant in Class 1 (not pathogenic), 2 (likely not pathogenic), 4 (likely pathogenic) or 5 (pathogenic).*

- Variant with probability of pathogenicity between 0.05-0.949 using a multifactorial likelihood model.

- In the absence of clinical evidence to assign an alternative classification, variant allele tested for mRNA aberrations using *in vitro* assays of patient RNA[4] that assess allele-specific transcript expression, and is found to produce mRNA transcript(s) predicted to encode intact full-length protein and/or isoforms that do not disrupt expression of one or more known clinically important residues[2].

- Variant that has insufficient evidence (molecular or otherwise) to be classified as a high-risk pathogenic variant or as a variant of little clinical significance.

- Variants located at positions listed in Appendix, Table 6, unless proven to fall in another class based on additional evidence.

- Variant considered possibly resistant to classification i.e. where there are multiple apparently conflicting points of evidence regarding variant pathogenicity, and which thus requires further investigation as a possible intermediate risk variant using alternative study design(s).

NO LONGER IN USE

### Class 2 – Likely not pathogenic/little clinical significance

*There is evidence against this variant being a dominant high-risk pathogenic variant and no substantial conflicting evidence.*

- Variants with probability of pathogenicity between 0.001-0.049 using a multifactorial likelihood model

- An exonic variant, that encodes the same amino acid change as a previously established Class 1 not pathogenic missense variant with a different underlying nucleotide change, and for which there is low bioinformatic likelihood[3] to disrupt normal splicing

- Synonymous substitution variant with low bioinformatic likelihood to disrupt normal splicing[3], with combined prior probability[6,7] of pathogenicity of ≤0.02 from clinically calibrated bioinformatic analyses.

NO LONGER IN USE

**Class1 – Not pathogenic/low clinical significance**

*There is significant evidence against this variant being a dominant high-risk pathogenic variant and no substantial conflicting evidence.*

- Variants with probability of pathogenicity <0.001 using a multifactorial likelihood model.

- Variants reported to occur in large outbred control reference groups at an allele frequency ≥1% (MAF ≥ 0.01)[5].

- An exonic variant, that encodes the same amino acid change as a previously established Class 1 not pathogenic missense variant with a different underlying nucleotide change, and for which there is no evidence of mRNA aberration (splicing or allelic imbalance) as determined using *in vitro* laboratory assays[4].

- Variants demonstrating all these features:
  - ✓ Exonic variant encoding a missense substitution or resulting in a small in-frame insertion/deletion or synonymous substitution with prior probability of pathogenicity ≤0.02 from clinically calibrated bioinformatic analyses[6], OR intronic variant
  - ✓ Low bioinformatic likelihood[3] to disrupt normal splicing
    OR

    Increased bioinformatic likelihood[3] to disrupt normal splicing **but** no associated mRNA aberration (splicing or allelic imbalance) as determined using *in vitro* laboratory assays[4].
  - ✓ Co-occurrence *in trans* with a known pathogenic sequence variant in the same gene in an individual who has no obvious additional clinical phenotype other than BRCA-associated cancer[8].
    OR

    Allele frequency ≥0.001 and <0.01 in large outbred control reference groups[9]

NO LONGER IN USE

### Footnotes

[1] To ensure robust variant classification based on multifactorial likelihood analysis results, the following caveats and recommendations should be noted before finalising variant classification. Only independent lines of evidence should be included. Tumour pathology information for a proband cannot be considered if the proband was selected for testing on the basis of breast tumour pathology criteria. Further investigation is necessary for any variant with extreme prior probability and minimal additional evidence, and it is currently recommended that clinical or laboratory evidence should contribute an LR of <0.5 (to reach final Class 2 (likely not pathogenic) or Class 1(not pathogenic)), or >2.0 (to reach final Class 4 (likely pathogenic) or Class 5 (pathogenic)). A variant which displays an obvious discordance between the predicted prior probability and additional clinical or laboratory evidence should be re-investigated to establish the veracity of results, to assess the possibility that it may be a hypomorph exhibiting intermediate or moderate penetrance relative to high-risk observed for classical pathogenic variants that encode a premature termination codon (nonsense or frameshift), and/or to exclude the very low probability that there is a pathogenic variant *in cis.* As per published recommendations (Plon et al., 2008), further research segregation testing in family members is recommended for variants in Class 2 (likely not pathogenic), Class 3 (uncertain) or Class 4 (likely pathogenic) to assist variant classification. For variants where no adequate prior has been estimated, eg UTR, intronic variants located at positions between +6 of one exon and -20 of the next exon, a prior of 0.02 will be applied; this assumes very conservatively that 2/100 of such variants might be associated with a high risk of cancer.

[2] Clinically important residues are defined by the location of sequence variants that introduce deleterious changes to protein function (via missense alteration, protein sequence deletion, or protein truncation in the last exon, AND are also associated with high risk of cancer. A clinically important functional protein domain is a recognized protein functional domain reported to harbor one or more clinically important residues. Physical boundaries for functional domains, and reported risk-associated variants to establish regions and residues of clinical importance, are described in the Appendix, Table 3 (BRCA1) and Table 4 (BRCA2).

[3] Bioinformatic likelihood of altering splicing is currently based on the MaxEntScan algorithm alone, in consideration of the following factors: several studies (Houdayer et al., 2012, Vallee et al., 2016) have reported that this tool has high sensitivity and specificity to detect splicing aberrations caused by abrogation of native donors and acceptors, or creation of de novo donors; there are published findings (Jian et al., 2014) and unpublished data (Spurdle lab, see Appendix, Supplementary Text) to indicate that use of multiple tools, in addition to MaxEntScan, does not improve prediction of spliceogenicity; MaxEntScan is publicly available and thus can be accessed by any clinical testing lab internationally. Empirical data were used to derive cutpoints for stratification of variants according to likelihood of spliceogenicity, taking into consideration score for the nearest native site. Further details, including score cut-points, are provided in the Appendix, Supplementary Text and Figures 1 and 2. Note: prediction of additional motifs relevant for normal splicing, such as Exonic Splicing Enhancers (ESEs), is not yet incorporated in these guidelines (see Appendix, Supplementary Text for justification). These may be considered in future iterations of these guidelines, when tools with sufficiently high sensitivity and specificity to predict severely altered splicing are available.

[4] Recommendations for the conduct and interpretation of mRNA assay data for variant classification are drawn from (Walker et al., 2013). Assessment assumes assays on mRNA from patient germline tissue samples (fresh blood, cultured lymphocytes, lymphoblastoid cell lines etc), compared with assays performed in tandem on mRNA from the same tissue type for ≥10 reference controls. Transcripts identified at similar levels in controls are considered to be naturally occurring isoforms and not mRNA aberrations. A variant is considered to be pathogenic due to an effect on mRNA transcript if it produces only transcript(s) carrying a premature stop codon or an in-frame deletion disrupting known functional domain(s)/residue(s), determined by semi-quantitative or quantitative methods. Sequencing of the full length transcript for the variant allele (if exonic), or a common polymorphism in *cis* (if variant is intronic), is currently considered adequate to assess if variant allele contributes to production of wild-type transcript. A variant may be reported as not associated with an mRNA aberration (splicing or expression) if the variant allele produces transcript patterns comparable to that of controls using assays conducted with nonsense-mediated decay inhibition. At this point in time, results from construct-based mRNA assays alone are not considered sufficiently robust to be used as evidence for variant classification on the basis of aberrant mRNA transcript profile.

NO LONGER IN USE

[5] Outbred control reference groups used for this purpose include datasets from the 1000 Genomes project (http://www.1000genomes.org), The Exome Aggregation Consortium (*ExAC*) – excluding cancer-related information from The Cancer Genome Atlas (TCGA) (http://exac.broadinstitute.org), the Exome Variant Server (http://evs.gs.washington.edu/EVS), the Genome Aggregation Database (gnomAD; http://gnomad.broadinstitute.org) – preferably excluding cancer-related information from the TCGA when available, Fabulous Ladies Over Seventy (FLOSSIES; https://whi.color.com).

[6] Prior probability of pathogenicity derived from calibration of Align-Grantham Variation Grantham Deviation score against *BRCA1/2* clinical features of variant pathogenicity (http://priors.hci.utah.edu/PRIORS/).

[7] Prior probability of pathogenicity derived from calibration of MaxEntScan score against *BRCA1/2* clinical features of variant pathogenicity (http://priors.hci.utah.edu/PRIORS/).

[8] Note: assuming a prior probability of 0.02, **the detection rate for class 5 pathogenic variants in that gene is required to be 0.025 in the sample set tested** to ensure that a single observation of co-occurrence *in trans* equates to a co-occurrence LR of 0.04, and consequently class 1 (not pathogenic) classification. This criterion also requires that the patient is assessed to exclude Fanconi-like or other features (Domchek et al., 2013, Sawyer et al., 2015) that suggest the variant leads to loss of function *in vivo*. If necessary, consider referral for examination by a clinical geneticist and/or additional *in vitro* diagnostic tests for molecular features of Fanconi phenotype.

[9] The lower bound of this allele frequency range was selected such that it would exclude the upper 99% confidence interval (binomial Exact) of the frequency observed for the most common pathogenic allele in multiple large reference datasets (termed maximum pathogenic allele frequency). Frequencies recorded 30 October 2016 are denoted below:

ExAC - non-Finnish European
* BRCA1 c.5329dupC p.Gln1777ProfsTer74 - 19/66740 alleles = 0.0002858 (99% CI 0.00014-0.00050)
* BRCA2 c.5946delT p.Ser1982ArgfsTer22 - 32/66540 alleles = 0.00048 (99% CI 0.00029-0.00075)
gnomAD - non-Finnish European (excludes Ashkenazi)
* BRCA1 5382insC c.5329dupC p.Gln1777ProfsTer74 - 19/126828 alleles = 0.00015 (99% CI 0.00008-0.00026)
* BRCA2 c.9097dupA p.Thr3033AsnfsTer11 & c.9097delA p.Thr3033LeufsTer29 - 13/109062 alleles = 0.00012 (99% CI 0.00005-0.00023)
gnomAD - most common other populations
* BRCA1 c.2025T>A p.Cys675Ter - 16/35712 Latino = 0.00044 (0.00024–0.00084)
* BRCA2 c.9097delA p.Thr3033LeufsTer29 - 2/16986 East Asian = 0.00012 (0.00005-0.00055)
* BRCA2 c.9097dupA p.Thr3033AsnfsTer11 - 4/35156 Latino = 0.00011 (0.00005-0.00036)

**Additional note regarding nomenclature**:

*BRCA1* and *BRCA2* variants are described using HGVS preferentially, with the simplified 1 letter presentation of amino acid substitutions and/or complete BIC designation also provided for comparison to historical reports.

Reference sequences are as follows:

- ***BRCA1.*** Coding DNA reference sequence from genomic refseq NG_005905.2 (same as LRG 292, Ensembl ENSG00000012048) covering BRCA1 transcript NM_007294.3 (Ensembl transcript ENST00000357654.7). Exon boundary numbering is from GenBank U14680.1, that is, exon 4 is missing due to a correction made after the initial description of the gene.
- ***BRCA2.*** Coding DNA reference sequence from genomic refseq NG_012772.3 (same as LRG 293, Ensembl ENSG00000139618), covering BRCA2 transcript NM_000059.3 (Ensembl transcript ENST00000544455.5).

NO LONGER IN USE

### APPENDIX

#### Table 1: IARC 5-tiered classification system with accompanying recommendations for family management[a]

| Class | Quantitative Measure: Probability of Pathogenicity | Predictive Testing of At-Risk Relatives | Surveillance for At-Risk Relatives | Research Testing of Relatives |
|---|---|---|---|---|
| 5: Pathogenic | >0.99 | Yes | Full high-risk guidelines for variant carriers | Not indicated |
| 4: Likely pathogenic | 0.95-0.99 | Yes[b] | Full high-risk guidelines for variant carriers | Yes |
| 3: Uncertain | 0.05-0.949 | No[b] | Based on family history & other risk factors | Yes |
| 2: Likely not pathogenic or of little clinical significance | 0.001-0.049 | No[b] | Based on family history & other risk factors - treat as "no *BRCA1/2* pathogenic variant detected" for this disorder | Yes |
| 1: Not pathogenic or of no clinical significance | <0.001 | No[b] | Based on family history & other risk factors - treat as "no *BRCA1/2* pathogenic variant detected" for this disorder disorder | Not indicated |

[a]Adapted for clarity from original tabular presentation published (Plon et al., 2008)

[b]Recommend continued testing of proband for any additional available testing modalities available for *BRCA1/2* e.g. rearrangements.

NOT IN USE

NO LONGER IN USE

### Table 2: Rationale for ENIGMA classification criteria

| Class | Criterion | Rationale for Criterion | Rationale/Summary of evidence stated for classification in ClinVar/other. |
|---|---|---|---|
| Class 5: pathogenic | Posterior probability of pathogenicity >0.99 from multifactorial likelihood analysis. | IARC recommendation for Class 5 Pathogenic (Plon et al., 2008) | IARC class based on posterior probability from multifactorial likelihood analysis, thresholds for class as per Plon et al. 2008 (PMID: 18951446). Class 5 Pathogenic based on posterior probability = [insert posterior]. |
| | Coding sequence variant encoding a premature termination codon i.e. nonsense/frameshift predicted to disrupt expression of clinically important functional domain(s)/residue(s). | Treated clinically as pathogenic | Variant allele predicted to encode a truncated non-functional protein. |
| | The variant allele produces only transcripts that lead to a premature stop codon, or in-frame deletion predicted to disrupt clinically important domains, as determined by RNA assays on patient germline tissue that assess allele-specific transcript expression. | Treated clinically as pathogenic | Allele-specific assay on patient-derived mRNA demonstrated that the variant allele produces only predicted non-functional transcripts. Variant allele produces [insert r.#_#del] transcript(s). |
| | Copy number deletion removing exon(s) spanning clinically important residue(s) ***or*** proven to result in a frameshift alteration predicted to interrupt expression of clinically important residue(s). | Treated clinically as pathogenic | Copy number deletion variant allele predicted to encode a non-functional protein. |
| | Copy number duplication proven to result in frameshift alteration predicted to interrupt expression of clinically important residue(s). | Treated clinically as pathogenic | Copy number duplication variant allele predicted to encode a non-functional protein. |
| Class 4: likely pathogenic | Posterior probability of pathogenicity 0.95-0.99 from multifactorial likelihood analysis. | IARC recommendation for Class 4 Likely Pathogenic (Plon et al., 2008) | IARC class based on posterior probability from multifactorial likelihood analysis, thresholds for class as per Plon et al. 2008 (PMID: 18951446). Class 4 Likely Pathogenic based on posterior probability = [insert posterior]. |

| | | | |
|---|---|---|---|
| | In the absence of clinical evidence to assign an alternative classification, sequence variant that alters the *BRCA1* or *BRCA2* translation initiation methionine site. | Bioinformatic and laboratory evidence indicates that a variant disrupting translation from the native initiation methionine site will result in lead to transcript(s) that encoding protein with abrogated function.<br><br>For *BRCA1*, the first in-frame methionine codon (p.M18) falls well within the RING domain, and the resulting N-truncated protein would delete several residues that are important for the BARD1 interaction (Starita et al., 2015).<br><br>For *BRCA2*, *in vitro* evidence indicates that preferential transcription from several out of frame ATGs located in the mRNA upstream of the first in-frame methionine codon (p.M124) indicates that very little protein synthesis will originate from p.M124 (Parsons et al., 2015). | *BRCA1*: A variant disrupting the native initiation start site is expected to lead to use of the first downstream in-frame methionine codon (p.M18), which lies well within the RING domain and will result in an N-truncated protein lacking a clinically important functional protein domain (PMID: 25823446).<br><br>*BRCA2*: A variant disrupting the native initiation start site is expected to lead to transcription from out-of-frame methionine codons, and transcripts predicted to encode a truncated non-functional protein (PMID: 24985344). |
| | Variant at IVS±1 or IVS±2 or G>non-G at last base of exon when adjacent intronic sequence is not GTRRGT that is predicted to alter used of native donor/acceptor site<br><br>AND is untested for splicing aberrations using RNA assays on patient blood that assess allele-specific transcript expression,<br><br>AND is not predicted or known to alter production of (naturally occurring) in-frame RNA isoforms that may rescue gene functionality. | Disruption of highly conserved bases at acceptor and donor splice sites is extremely likely to result in a splicing aberration, with suggested prior probability 0.96 (Walker et al., 2013). Conservative classification is warranted since pathogenicity cannot be assumed for all mRNA profiles arising from a variant allele e.g. incomplete effect on splicing, or potential to lead to in-frame transcripts encoding functional protein | Consensus donor/acceptor site variant allele with high likelihood to result in splicing aberration with pathogenic consequences; G>non-G change at last base of the exon with high bioinformatic likelihood to result in splicing aberration with pathogenic consequences. |
| | A variant that encodes the same amino acid change as a previously established Class 5 pathogenic **missense** variant with a different underlying nucleotide change, is located in a known clinically important functional protein domain, with no evidence of mRNA aberration (splicing or expression) from in vitro mRNA assays on patient RNA, and the variant is absent from outbred control reference groups. | Having excluded possible mRNA defects caused by a nucleotide change, the clinical consequences of a rare missense variant should be equivalent irrespective of the underlying nucleotide change. Absence in controls and location in a functional domain provides additional support for evidence of pathogenicity. | Variant allele encodes same protein change as a proven pathogenic **missense** allele, and does not alter mRNA splicing. |
| | A small in-frame deletion variant that removes a codon for which a missense substitution Class 5 pathogenic variant has been described, is located in a known clinically important functional protein | For a given amino acid, the clinical consequences are equivalent for an in-frame deletion and the most severe missense substitution. Absence in controls and | Variant allele deletes an amino acid critical to function and proven to be associated with disease when altered. |

NO LONGER IN USE

| | | | |
|---|---|---|---|
| | domain, unlikely to result in an alternative aberration via mRNA splicing, and is absent from outbred control reference groups. | location in a functional domain provides additional support for evidence of pathogenicity. | |
| Class 3: uncertain | Posterior probability of pathogenicity 0.05-0.949 from multifactorial likelihood analysis. | IARC recommendation for Class 3 Uncertain (Plon et al., 2008) | IARC class based on posterior probability from multifactorial likelihood analysis, thresholds for class as per Plon et al. 2008 (PMID: 18951446). Class 3 Uncertain based on posterior probability = [insert posterior]. |
| | In the absence of clinical evidence to assign an alternative classification, variant allele tested for mRNA aberrations using *in vitro* assays of patient RNA that assess allele-specific transcript expression, and is found to produce mRNA transcript(s) predicted to encode intact full-length protein and/or isoforms that do not disrupt known clinically important functional residue(s). | Variant leads to transcript profile of equivocal clinical significance. | Insufficient evidence to determine clinical significance. Variant allele produces [insert "full-length transcript" AND/OR "in-frame r.#_#del transcript (encoding potentially functional protein)"] |
| | Insufficient evidence to classify variant. | Does not fit prescribed criteria for other classes | Insufficient evidence to determine clinical significance. |
| | Variant located at position listed in Table 6, unless proven to fall in another class based on additional evidence. | Variant has potential to lead to in-frame (naturally-occurring) transcripts that may rescue gene functionality. | Variant may result in mRNA transcript(s) that encode functional proteins. |
| | Variant with conflicting evidence for pathogenicity. | Variant with modest effect on gene/protein function and modest/intermediate effect on risk may demonstrate some but not all features of a high-risk pathogenic variant, and should be highlighted for assessment of risk using alternative approaches. | Conflicting evidence for pathogenicity; potential intermediate risk variant. |
| Class 2: Likely not pathogenic or of little clinical significance | Posterior probability of pathogenicity 0.001-0.049 from multifactorial likelihood analysis. | IARC recommendation for Class 2 Likely Not Pathogenic (Plon et al., 2008) | IARC class based on posterior probability from multifactorial likelihood analysis, thresholds for class as per Plon et al. 2008 (PMID: 18951446). Class 2 Likely Not Pathogenic based on posterior probability = [insert posterior]. |
| | An exonic variant, that encodes the same amino acid change as a previously established Class 1 not pathogenic missense variant with | With low likelihood of splicing aberration, the clinical consequences of a missense variant should be | Variant allele is predicted to not alter splicing and to encode the same protein change as a **missense** allele already |

NO LONGER IN USE

| | | | |
|---|---|---|---|
| | a different underlying nucleotide change, and for which there is low bioinformatic likelihood to disrupt normal splicing. | equivalent irrespective of the underlying nucleotide change. | proven to be not pathogenic or of little clinical significance. |
| | Synonymous substitution with low bioinformatic likelihood to disrupt normal splicing, determined to have combined prior probability of pathogenicity ≤0.02 from clinically calibrated bioinformatic analyses. | A silent substitution variant that is not bioinformatically predicted to effect mRNA function is extremely unlikely to result in clinical consequences equivalent to a high-risk pathogenic variant, as indicated by prior probability of ≤0.02 for variants in this stratum from analysis calibrating synonymous changes (Tavtigian, unpublished data, 2008) and bioinformatic predictions of variant effect on splicing against clinical information (Vallee et al., 2016). | Synonymous substitution variant, with low bioinformatic likelihood to result in a splicing aberration (Splicing prior probability 0.02; (http://priors.hci.utah.edu/PRIORS/) |
| Class 1: not pathogenic or of no clinical significance | Posterior probability of pathogenicity <0.001 from multifactorial likelihood analysis | IARC recommendation for Class 1 Not Pathogenic (Plon et al., 2008) | IARC class based on posterior probability from multifactorial likelihood analysis, thresholds for class as per Plon et al. 2008 (PMID: 18951446). Class 1 Not Pathogenic based on posterior probability = [insert posterior]. |
| | Variant with reported frequency ≥1% in large outbred control reference groups | High-risk variants are not common in the general population, and outbred reference groups exclude the possibility that a selected variant is an undetected founder "mutation" | Not pathogenic based on frequency >1% in an outbred sampleset. Frequency [insert MAF (Population)], derived from [insert dataset (yyyy-mm-dd)]. |
| | Exonic variant that encodes the same amino acid change as a previously established Class 1 Not Pathogenic **missense** variant with a different underlying nucleotide change, and for which there is no evidence of mRNA aberration from in vitro mRNA assays. | Having excluded possible mRNA defects caused by a nucleotide change, the clinical consequences of a missense variant should be equivalent irrespective of the underlying nucleotide change. | Variant allele does not alter splicing and is predicted only to encode the same protein change as a **missense** allele already proven to be not pathogenic or of little clinical significance |
| | Exonic variant encoding a missense substitution or a small in-frame insertion/deletion with prior probability≤0.02 from clinically calibrated bioinformatic analyses OR intronic variant<br><br>AND<br><br>Low bioinformatic likelihood to disrupt normal splicing | Multiple points of evidence indicate the variant is unlikely to be associated with high risk: variant is unlikely to affect protein function (from bioinformatic predictions); variant is predicted bioinformatically or shown from *in vitro* mRNA assays to result in normal splicing (mRNA assays); *in vivo* evidence for proficient function is indicated by co-occurrence *in trans* with a known pathogenic variant in the same gene and with no unusual clinical features, or observation that allele | Variant allele has low bioinformatic likelihood to encode a missense alteration affecting protein function (Missense prior probability 0.02; http://priors.hci.utah.edu/PRIORS/),<br><br>AND low bioinformatic likelihood to alter mRNA splicing (splicing prior 0.xx; http://priors.hci.utah.edu/PRIORS/) OR |

NO LONGER IN USE

| | | | |
|---|---|---|---|
| | OR Increased bioinformatic likelihood to disrupt normal splicing **but** no associated mRNA aberration (splicing or allelic imbalance) as determined using *in vitro* laboratory assays<br><br>AND<br><br>Co-occurrence *in trans* with a known pathogenic sequence variant in the same gene in an individual with no obvious additional clinical phenotype other than BRCA-associated cancer<br><br>OR Allele frequency ≥0.001 and <0.01 in large outbred control reference groups. | frequency in reference groups is greater than would be expected for a single non-founder variant leading to a dominant disorder | does not alter splicing *in vitro* [insert reference]<br><br>AND co-occurrence *in trans* with pathogenic variant [insert pathogenic variant] in patient [insert clinical features, age etc], reported in [insert reference]]. OR<br><br>Frequency [insert MAF (Population)], derived from [insert dataset (yyyy-mm-dd)]. |

NO LONGER IN USE

**Table 3: Catalogue of BRCA1 conserved domains/motifs and currently known clinically important amino acid residues, and relevance for classification of *BRCA1* in-frame and terminal exon sequence variants.**

| Domain/ Motif | AA start | AA end | AA alterations with Demonstrated Clinical Importance[a] | Classification of in-frame deletions targeting domain/motifs | References and summary interpretation[a] |
|---|---|---|---|---|---|
| RING | 1 | 101 | L22S (c.65T>C (p.Leu22Ser))<br>T37K (c.110C>A (p.Thr37Lys))<br>C39R (c.115T>C (p.Cys39Arg))<br>H41R (c.122A>G (p.His41Arg))<br>C44S (c.130T>A (p.Cys44Ser))<br>C44Y (c.131G>A (p.Cys44Tyr))<br>C61G (c.181T>G (p.Cys61Gly)) | Class-5 if at least one clinically relevant residue is removed. Class-3 otherwise. | http://www.ncbi.nlm.nih.gov/protein/15988069;<br><br>http://hci-exlovd.hci.utah.edu; Multifactorial analysis for H41R (c.122A>G (p.His41Arg)) (Whiley et al., 2014). |
| NES | 81 | 99 | None reported | Class-3 | Domain location description (Rodriguez and Henderson, 2000) |
| NLS1 | 503 | 508 | None reported | Class-3 | Domain location description (Chen et al., 1996, Thakur et al., 1997). |
| NLS2 | 607 | 614 | None reported | Class-3 | Domain location description (Chen et al., 1996, Thakur et al., 1997). |
| NLS3 | 651 | 656 | None reported | Class-3 | Domain location description (Chen et al., 1996). |
| COILED-COIL | 1391 | 1424 | None reported | Class-3 | Domain location description (Hu et al., 2000) |
| BRCT DOMAINS | 1650 | 1863 | T1685A (c.5053A>G (p.Thr1685Ala))<br>T1685I (c.5054C>T (p.Thr1685Ile))<br>V1688del (c.5062_5064del (p.Val1688del))<br>R1699W (c.5095C>T (p.Arg1699Trp))<br>G1706E (c.5117G>A (p.Gly1706Glu))<br>A1708E (c.5123C>A (p.Ala1708Glu))<br>S1715R (c.5143A>C (p.Ser1715Arg))<br>G1738R (c.5212G>A (p.Gly1738Arg))<br>L1764P (c.5291T>C (p.Leu1764Pro))<br>I1766S (c.5297T>G (p.Ile1766Ser))<br>M1775K (c.5324T>A (p.Met1775Lys))<br>M1775R (c.5324T>G (p.Met1775Arg))<br>C1787S (c.5359T>A (p.Cys1787Ser))<br>G1788V (c.5363G>T (p.Gly1788Val))<br>V1838E (c.5513T>A (p.Val1838Glu)) | Class-5 if at least one clinically relevant residue is removed. Class-3 otherwise. | Domain boundaries derived from X-ray chrystallography data are aa1646-1863 (1T15, http://www.ncbi.nlm.nih.gov/Structure/mmdb/mmdbsrv.cgi?uid=27907), and ENIGMA functional assay data (Monteiro, unpublished)<br><br>Digestion data indicate aa1860-1863 are dispensable based on susceptibility to digestion (Lee et al., 2010), while pathogenic variant data indicate that 1855-1862 are dispensable (Hayes et al., 2000). Position 1854 is implicated as clinically important by the observation that Y1853X (c.5559C>G (p.Tyr1853Ter) is a recognized high-risk pathogenic variant.<br>These data combined indicate that position 1854 or 1855 is the C-terminal border of the BRCT/BRCA1 relevant to clinical interpretation of sequence variants in exon 24 of BRCA1. That is, a variant predicted to |

NO LONGER IN USE

| | | | | | |
|---|---|---|---|---|---|
| | | | | | disrupt expression of protein sequence only downstream* of position 1855 would not be considered clinically important. |

[a] Missense substitutions in denoted functional domains that are designated as Class 5 pathogenic based on multifactorial likelihood posterior probability of pathogenicity > 0.99 (listed in http://hci-exlovd.hci.utah.edu, or individual references noted), and for which there is no/little effect on mRNA transcript profile, ***unless*** the variant results in an aberrant transcript that encodes a discrete in-frame deletion considered informative to definition of clinically important domains.
*Typo was corrected in version 2.5.1

Note - The following pathogenic exonic variants known to alter mRNA splicing have been excluded from Table 3 above, as justified below:

| Variant | mRNA Change | Predicted Protein Change | Reason for exclusion |
|---|---|---|---|
| *BRCA1* R1495M (c.4484G>T (p.Arg1495Met)) | r.[4358_4484del, 4358_4675del] | p.(Ala1453GlyfsTer10) - predominant transcript | Predominant alternate transcript is out-of-frame. Loss of function assumed due to loss of full length transcript from variant allele (Houdayer et al., 2012, Colombo et al., 2013, Santos et al., 2014). |
| *BRCA1* E1559K (c.4675G>A (p.Glu1559Lys)) | r.[4665_4675del] | p.(Gln1366AlafsTer13) | Alternate transcript is out-of-frame. Level of full length transcript not assessed (Wappenschmidt et al., 2012). |
| *BRCA1* A1623G (c.4868C>G (p.Ala1623Gly)) | r.[4868_4986del] | p.(Ala1623AspfsTer16) | Alternate transcript is out-of-frame. Variant allele produces some full length transcript (Walker et al., 2010) |
| *BRCA1* D1692N (c.5074G>A (p.Asp1692Asn)) | r.[4987_5074del, 5074_5075ins5074+1_5074+153] | p.(Val1665SerfsTer8) – predominant transcript | Predominant alternate transcript, based on minigene assay (Ahlborn et al., 2015), is out of frame. |

NO LONGER IN USE

**Table 4: Catalogue of BRCA2 conserved domains/motifs and currently known clinically important amino acid residues, and relevance for classification of *BRCA2* in-frame and terminal exon sequence variants**

| Domain/ Motif | AA start | AA end | AA alterations with Demonstrated Clinical Importance[a] | Classification of in-frame deletions targeting domain/motifs | References and summary interpretation[a] |
|---|---|---|---|---|---|
| PALB2 Binding | 10 | 40 | None reported | Class-3 | Domain location description (Oliver et al., 2009, Xia et al., 2006) |
| BRC-1 | 1002 | 1036 | None reported | Class-3 | http://www.ncbi.nlm.nih.gov/protein/NP_000050.2 |
| BRC-2 | 1212 | 1246 | None reported | Class-3 | http://www.ncbi.nlm.nih.gov/protein/NP_000050.2 |
| BRC-3 | 1422 | 1453 | None reported | Class-3 | http://www.ncbi.nlm.nih.gov/protein/NP_000050.2 |
| BRC-4 | 1518 | 1549 | None reported | Class-3 | http://www.ncbi.nlm.nih.gov/protein/NP_000050.2 |
| BRC-5 | 1665 | 1696 | None reported | Class-3 | http://www.ncbi.nlm.nih.gov/protein/NP_000050.2 |
| BRC-6 | 1837 | 1871 | None reported | Class-3 | http://www.ncbi.nlm.nih.gov/protein/NP_000050.2 |
| BRC-7 | 1971 | 2005 | None reported | Class-3 | http://www.ncbi.nlm.nih.gov/protein/NP_000050.2 |
| BRC-8 | 2051 | 2085 | None reported | Class-3 | http://www.ncbi.nlm.nih.gov/protein/NP_000050.2 |
| DBD (DNA/DSS 1 binding domain - helical, OB1, OB2, OB3) | 2481 | 3186 | W2626C (c.7878G>C (p.Trp2626Cys))<br>I2627F(c.7879A>T (p.Ile2627Phe))<br>E2663V (c.7988A>T (p.Glu2663Val))<br>T2722R (c.8165C>G (p.Thr2722Arg))<br>D2723G (c.8168A>G (p.Asp2723Gly))<br>D2723H (c.8167G>C (p.Asp2723His))<br>G2748D (c.8243G>A (p.Gly2748Asp))<br>I2778_Q2829del (c.8332_8487del (p.Ile2778_Gln2829del))<br>R3052W (c.9154C>T (p.Arg3052Trp)) | Class-5 if at least one clinically relevant residue (or all of AA 2778-2829) is removed.<br><br>Class-3 otherwise | http://www.ncbi.nlm.nih.gov/protein/NP_000050.2; http://hci-exlovd.hci.utah.edu.<br><br>Pathogenic variant c.8486G>A (also recorded as Gln2829Arg) results in a transcript encoding an in-frame exon 19 deletion only (Houdayer et al., 2012), indicating that genetic variation encompassing loss of this entire exon (AA2778-2829) should be considered clinically important.The clinical impact of alteration/deletion of individual amino acids in exon 19 is not yet established. |
| NLS1 | 3263 | 3269 | None reported | Class-3 | Domain location description (Guidugli et al., 2014) |
| BRC-9 or TR2 | 3265 | 3330 | None reported | Class-3 | Note, although amino acids 3270-3305 within this fragment is reported to bind RAD51-DNA filaments (Davies and Pellegrini, 2007), there is no sequence conservation with the BRC repeats located between aa1002 and |

NO LONGER IN USE

| | | | | | |
|---|---|---|---|---|---|
| | | | | | aa2014. Domain boundaries are derived from x-ray chrystallography data are aa3265-3330 (Esashi et al., 2005, Esashi et al., 2007).<br><br>Case-control and frequency data indicate that BRCA2 c.9976A>T (p.Lys3326Ter) does not confer a high risk of cancer (OR 1.3-1.5, dependent on breast or ovarian cancer subtype (Meeks et al., 2016), demonstrating that residues at and downstream of 3327 are likely dispensable.<br><br>Position 3308 is implicated as clinically important by the observation that a nonsense variant c.9924C>G (p.Tyr3308Ter) is recognized as a high-risk pathogenic variant with known functional relevance ((Vallee et al., 2016); Bayes score 1122:1 from a single large kConFab family, Spurdle unpublished data). There is currently no publicly available clinical information to support pathogenicity of nonsense or frameshift variants located between positions 3309 and 3325.<br><br>These data combined suggest that the C-terminal border of the BRC-9 relevant to the clinical interpretation of sequence variants in exon 27 of BRCA2 lies between 3309 and 3325. That is, a variant predicted to disrupt expression only of protein sequence downstream of position 3325 would be considered unlikely to be clinically important. Further functional and clinical studies are underway to refine risk, if any, for predicted nonsense or frameshift variants downstream of position 3326. |
| NLS2 | 3381 | 3385 | No | Class-3 | Domain location description (Guidugli et al., 2014).<br>This domain is considered unlikely clinically relevant since it lies downstream of position 3326. |

[a] Missense substitutions in denoted functional domains that are designated as Class 5 pathogenic based on multifactorial likelihood posterior probability of pathogenicity > 0.99, and for which there is no/little effect on mRNA transcript profile - ***unless*** the variant results in an aberrant transcript that encodes a discrete in-frame deletion considered informative to definition of clinically important domains. (Splicing aberrations are reported for *BRCA2* c.7988A>T (p.Glu2663Val) and c.8168A>G (p.Asp2723Gly) (Walker et al., 2010), but these did not lead to complete loss of function of the full length transcript), and missense alterations showed abrogation of functional activity using multiple assays (Walker et al., 2010). An additional conserved region not commonly recognized as a BRCA2 domain/motif is located AA 1110-1183, but no pathogenic missense substitutions have been recorded for this region.

Note – The following pathogenic exonic variants known to alter mRNA splicing have been excluded from Table 4 above, as justified below:

| Variant | mRNA Change | Predicted Protein Change | Reason for exclusion |
|---|---|---|---|
| *BRCA2* R2659K (c.7976G>A (p.Arg2659Lys)) | r.[7806_7976del] | p.(Ala2603_Arg2659del) | Alternate transcript is in-frame but level of full length transcript not assessed (Farrugia et al., 2008) |

NO LONGER IN USE

| | | | |
|---|---|---|---|
| *BRCA2* R2659T (c.7976G>C (p.Arg2659Thr)) | r.[7806_7976del] | p.(Ala2603_Arg2659del) | Alternate transcript is in-frame but level of full length transcript not assessed (Farrugia et al., 2008) |
| *BRCA2* P3039P (c.9117G>A (p.Pro3039Pro)) | r.[8954_9117del] | p.(Val2985Glyfs*4) | Allele-specific assay shows out-of-frame transcript (Houdayer et al., 2012) |

**Table 5: *BRCA1* and *BRCA2* donor site intronic sequence, as relevant for inferring spliceogenicity of variants at the last base of the exon:** G>non-G substitutions at the last base of the exons noted in red font are considered Class 4 Likely Pathogenic in the absence of additional clinical/ laboratory data.*

| Gene | Exon | First 6 bases of the intron | Donor GTRRGT? | Polymorphism in first 6 bp?* | Max allele frequency* |
|---|---|---|---|---|---|
| BRCA1 | Exon 1 | GTAGTA | N | | |
| BRCA1 | Exon 2 | GTAAGT | Y | | |
| BRCA1 | Exon 3 | GTAAGT | Y | | |
| BRCA1 | Exon 5 | GTATAT | N | | |
| BRCA1 | Exon 6 | GTAAGT | Y | | |
| BRCA1 | Exon 7 | GTAAAA | N | rs200358748 (+5A>G) | 0.001 EUR |
| BRCA1 | Exon 8 | GTAAGG | N | | |
| BRCA1 | Exon 9*** | GTGAGT | Y | rs80358013 (+3G>A) | 0.001 AFR |
| BRCA1 | Exon 10*** | GTAATG | N | | |
| BRCA1 | Exon 11*** | GTATTG | N | | |
| BRCA1 | Exon 12 | GTAAAA | N | | |
| BRCA1 | Exon 13 | GTGTGT | N | | |
| BRCA1 | Exon 14 | GTAAGA | N | | |
| BRCA1 | Exon 15 | GTAATA | N | | |

NO LONGER IN USE

Version 2.5.1 29 June 2017

| | | | |
|---|---|---|---|
| BRCA1 | Exon 16 | GTGAGT | Y |
| BRCA1 | Exon 17 | GTATAC | N |
| BRCA1 | Exon 18 | GTAAGT | Y |
| BRCA1 | Exon 19 | GTAAGT | Y |
| BRCA1 | Exon 20 | GTAAAG | N |
| BRCA1 | Exon 21 | GTAAGA | N |
| BRCA1 | Exon 22 | GTAAGT | Y |
| BRCA1 | Exon 23 | GTAAGG | N |
| BRCA2 | Exon 1 | GTTAGT | Y |
| BRCA2 | Exon 2 | GTATTG | N |
| BRCA2 | Exon 3 | GTAAGT | Y |
| BRCA2 | Exon 4 | GTATGA | N |
| BRCA2 | Exon 5 | GTATGA | N |
| BRCA2 | Exon 6 | GTAAAT | N |
| BRCA2 | Exon 7 | GTAATA | N |
| BRCA2 | Exon 8 | GTAAGT | Y |
| BRCA2 | Exon 9 | GTAAGT | Y |
| BRCA2 | Exon 10 | GTACCT | N |
| BRCA2 | Exon 11 | GTAAGT | Y |

NO LONGER IN USE

| | | | | | |
|---|---|---|---|---|---|
| BRCA2 | Exon 12*** | GTAAAA | N | | |
| BRCA2 | Exon 13 | GTAAGA | N | | |
| BRCA2 | Exon 14 | GTATTG | N | rs81002852 (+6G>A) | 0.004 AFR |
| BRCA2 | Exon 15 | GTATGT | N | | |
| BRCA2 | Exon 16 | GTACTC | N | rs81002819 (+6C>G) | 0.006 AFR |
| BRCA2 | Exon 17 | GCAAGT | N | | |
| BRCA2 | Exon 18 | GTAAAT | N | | |
| BRCA2 | Exon 19 | GTATGA | N | | |
| BRCA2 | Exon 20 | GTAAAA | N | | |
| BRCA2 | Exon 21 | GTGAGA | N | | |
| BRCA2 | Exon 22 | GTAAGT | Y | | |
| BRCA2 | Exon 23 | GTACAA | N | | |
| BRCA2 | Exon 24 | GTAATG | N | | |
| BRCA2 | Exon 25 | GTAAGG | N | | |
| BRCA2 | Exon 26 | GTAAGT | Y | | |

* It is advised that the spliceogenicity/pathogenicity of any variant in an individual should take into consideration bioinformatic prediction that takes into consideration local sequence context (eg use of cryptic sites to produce in-frame products), and other variants reported in that individual that may lie *in cis* to the variant under review.

** Variants with minor allele frequency ≥ 0.0001 in 1000 Genomes, Exome Variant Server and EXAC as at April 2016. EUR=European, AFR=African sub-populations. None of these polymorphisms alter the GTRRGT annotation for *BRCA1*/2 introns.

*** Although variants at this position are proven/highly likely to alter splicing, they are also predicted to lead to production of in-frame mRNA isoforms that may rescue functionality (See Table 6), and thus should be considered class 3 uncertain in the absence of additional clinical information.

NO LONGER IN USE

**Table 6: *BRCA1* and *BRCA2* exon boundary variants predicted or known to lead to naturally occurring in-frame RNA isoforms that may rescue gene functionality.** *Variants at these positions should be considered Class 3 Uncertain unless proven otherwise.**

| Gene | Alternative Splicing Event | Variants Implicated | Rationale |
|---|---|---|---|
| *BRCA1* | $\Delta$8p | c.442-1 (IVS7-1)<br>c.442-2 (IVS7-2) | *BRCA1* exon 8 acceptor site is an experimentally validated tandem acceptor site (NAGNAG) subject to alternative splicing (Colombo et al., 2014). c.442-1,-2 variants are predicted to inactivate the 5' acceptor site, but not the 3' acceptor site, thus producing $\Delta$8p transcripts. |
| | $\Delta$9,10 | c.548-1 (IVS8-1)<br>c.548-2 (IVS8-2)<br>c.593 to non-G<br>c.593+1 (IVS9+1)<br>c.593+2 (IVS9+2)<br>c.594-1 (IVS9-1)<br>c.594-2 (IVS9-2)<br>c.670 to non-G<br>c.670+1 (IVS10+1)<br>c.670+2 (IVS10+2) | Carriers of variants at these positions are predicted to produce normal (or increased) levels of *BRCA1* $\Delta$(9,10), a major in-frame alternative splicing event (Colombo et al., 2014).<br><br>T*he BRCA1* variant c.594-2A>C (shown from ENIGMA research to co-occur in cis with c.641A>G), has been reported to demonstrate clinical characteristics inconsistent with a high risk of cancer expected for a pathogenic *BRCA1* variant (Rosenthal et al., 2015) . The haplotype of c.[594-2A>C; 641A>G] has been shown from mRNA analysis in human samplesto produce high levels of $\Delta$10 transcripts (˜70% of the overall expression, and **has been designated as Class 1 Not Pathogenic** by the ENIGMA Consortium using multifactorial likelihood analysis that includes genetic (segregation, case-control analysis) and pathology data (de la Hoya et al., 2016). |
| | $\Delta$11q, $\Delta$11 | c.4096 to non-G<br>c.4096+1 (IVS11+1)<br>c.4097+2 (IVS11+2) | Data collected by the ENIGMA consortium demonstrates that the *BRCA1* c.4096+1G>A variant, proven to result in production of naturally occurring in-frame transcripts Δ11q (Bonatti et al., 2006) and also Δ11 (Radice, unpublished data), may not exhibit the clinical characteristics of a standard high-risk pathogenic *BRCA1* variant (Spurdle, unpublished data). |
| | $\Delta$13p | c.4186-1 (IVS12-1)<br>c.4186-2 (IVS12-2) | *BRCA1* exon 13 acceptor site is an experimentally validated tandem acceptor site (NAGNAG) subject to alternative splicing (Colombo et al., 2014). c.4186-1,-2 variants are predicted to inactivate the 5' acceptor site, but not the 3' acceptor site, thus producing $\Delta$13p transcripts |
| | $\Delta$14p | c.4358-1 (IVS13-1)<br>c.4358-2 (IVS13-2) | *BRCA1* exon 14 acceptor site is an experimentally validated tandem acceptor site (NAGNAG) subject to alternative splicing (Colombo et al., 2014). c.4358-1,-2 variants are predicted to inactivate the 5' acceptor site, but not the 3' acceptor site, thus producing $\Delta$14p transcripts |
| *BRCA2* | $\Delta$12 | c.6842-1 (IVS11-1)<br>c.6842-2 (IVS11-2)<br>c.6937 to non-G<br>c.6937+1 (IVS12+1)<br>c.6937+2 (IVS12+2) | Carriers of these variants are predicted to produce exon12 skipping. BRCA2 $\Delta$12 is a naturally occurring in-frame splicing event (Fackenthal et al., 2016). BRCA2 exon12 is functionally redundant (Li et al., 2009) |

NO LONGER IN USE

*This summary table does not yet capture the possibility of acceptor site changes leading to small in-frame deletions > 3bp e.g. due to NAG $(NNN)_n$ NAG sites. It is recommended that bioinformatic prediction analysis is carried out for variation in/near *all* donor and acceptor sites to assess the likelihood that a variant will or will not alter native splicing.
Note - It could be argued that nonsense or frameshift variants in *BRCA1* exon 9, *BRCA1* exon 10, or *BRCA2* exon 12 may not be associated with high risk of cancer due to rescue by expression of in-frame naturally occurring isoforms that bypass the premature termination codon, and thus encode a functional protein. Review of multiple clinical and control datasets for frequency of unique nonsense or frameshift variants - with adjustment for exon size - does not provide strong support for this hypothesis at the present time (Spurdle, de la Hoya, unpublished data). Additional research is underway to further investigate the functional/clinical importance of germline nonsense or frameshift variants in these exons.
Moreover, further work is planned within ENIGMA (led by Paolo Radice) to document variants that have undergone splicing assays and are proven to be "leaky" variants, to provide a record of all spliceogenic variants for which additional research is necessary. This resource will identify variants that have already been classified using clinical data, as positive and negative controls for future quantitative mRNA studies.

NO LONGER IN USE

**Supplementary Text: Splicing algorithms for qualitative classification**

Bioinformatic prediction of splicing, outside of application of a prior probability, is required to implement qualitative criteria that include a statement about bioinformatic likelihood to disrupt normal splicing, Detailed analysis was undertaken in the Spurdle laboratory to inform selection of bioinformatic splicing tool(s), score cut-offs, and define the overarching scheme for selecting potentially spliceogenic variants and the most appropriate classification depending on their likely consequences.

The dataset was based on exonic *BRCA1* and *BRCA2* variants for which splicing assay results had been reported in the literature to the end of 2015; there were currently 650 distinct assay results for *BRCA1/2* variants recorded (353 no aberration). Of these 291 are intronic, and 359 exonic (237 no aberration). Some variants have been assayed multiple times, and where results were not the same, the most recent/appropriate results were recorded. A subset of 176 assayed exonic variants were selected for bioinformatic scoring, including all variants with a reported aberration (n=60 unique variants), and a subset of those with no aberration observed (n=76 unique variants). Bioinformatic predictions were generated for native donor and acceptors, and these selected exonic variants, using 5 tools commonly used in the clinical setting: SSF, MaxEntScan, NNSPLICE, GeneSplicer, and HSF. GeneSplicer did not provide a score for 10/44 native sites for *BRCA1* (6 donor, 4 acceptor), and 22/52 native sites for *BRCA2* (17 donor, 5 acceptor); because of this poor prediction of native sites, GeneSplicer was dropped from further analysis. For each variant, scores were recorded for (possible) donor or acceptor sites: for a variant with a reported aberration the score was selected as appropriate to the type and position of the aberration reported; for a variant with no splicing aberration, the scores recorded were the highest donor and acceptor predictions at/near the variant (within 15bp). For variants where no score was returned (not unexpected since the variant is not causing an aberration), a score of 0 was assigned. In addition, distance from position scored to nearest native donor or acceptor was calculated, as was the difference in score between native donor/acceptor and the variant score. The value of % change in score was considered, however it became clear that some variants with no predicted effect on splicing could nevertheless exhibit a big change in score e.g. scores of 0>>0 with 250% change were identified to be due to miniscule changes in score beyond the second decimal point. Variants were also scored for possible effect on an ESS/ESE using the ESRseq algorithm described by Gaildrat *et al* (Gaildrat et al., 2012)

The final dataset used for the main analysis - assessing prediction of variant effect on donor/acceptor site usage - included variants with the following molecular outcomes: 29 donor site loss, 2 acceptor site loss, 9 donor site gain, 1 acceptor site gain (a result which could be a mis-interpretation of a naturally occurring isoform), and 76 variants with no aberration (scored for effect on donor and acceptor sites). Another 18 variants that appeared to lead to aberrations as a result of effect on exonic splice enhancers or silencers (ESE/ESS) were included only in secondary analysis assessing sensitivity and specificity of predicting variant effect on ESE/ESS, with the 76 variants. demonstrating no aberration as reference set. Forward logistic regression

NO LONGER IN USE

analysis was used to investigate which tools, scores and other factors best predicted presence/absence of a splicing aberration due to loss or creation of a donor/acceptor.

MaxEnt Scan was the best predictor of aberration due to donor site gain (89% sensitivity, 97% specificity), and inclusion of scores for additional tools did not improve predictions. These results are consistent with findings from Jian et al (Jian et al., 2014), using a larger but relatively "uncurated" dataset. Raw MaxEntScan scores were then compared to the empirical splicing results (aberration or not), and also to MaxEntScan scores used to define cutpoints for prior probability of pathogenicity (due to spliceogenicity) as defined by Vallee et al for single nucleotide substitution variants (Vallee et al., 2016).

Irrespective of the tool used, aberration for exonic variants due to donor or acceptor site loss was best predicted by distance, reflecting the bias in selection of these variants for mRNA assays and for reporting in publication form. As for donor gains, raw MaxEntScan scores were compared to empirical splicing results to inform selection of cutpoints. These comparisons, together with relationship between variant score and nearest native score, were used to inform selection of appropriate cut-offs for prioritizing variants likely to be spliceogenic. The outcome was a 3 tier scheme for predicting capacity to cause altered splicing based on raw score, with promotion of a moderate category variant to a higher spliceogenicity category if native site score indicates capacity of the variant to outcompete the nearest native site.

ESRseq score alone was a poor predictor of splicing aberrations induced by alteration of an ESE/ESS site (22% sensitivity), although ESRseq score could be considered as a negative predictor of aberration (96% specificity). Other factors such as difference in score to wildtype sequence and distance to the nearest donor/acceptor, added no value to the prediction. ESE prediction was thus not included in schemes for variant classification.

The practical use of splicing predictions for assessing spliceogenicity of coding sequence variants assumed to encode a premature termination codon is summarized in Figure 2.

The suggested use of splicing predictions for assessing spliceogenicity of coding sequence variants assumed to encode a non-synonymous or missense substitution, or intronic non-coding variants will be documented in future iterations of these rules.

NO LONGER IN USE

**Figure 1: Overview of ENIGMA classification process.**

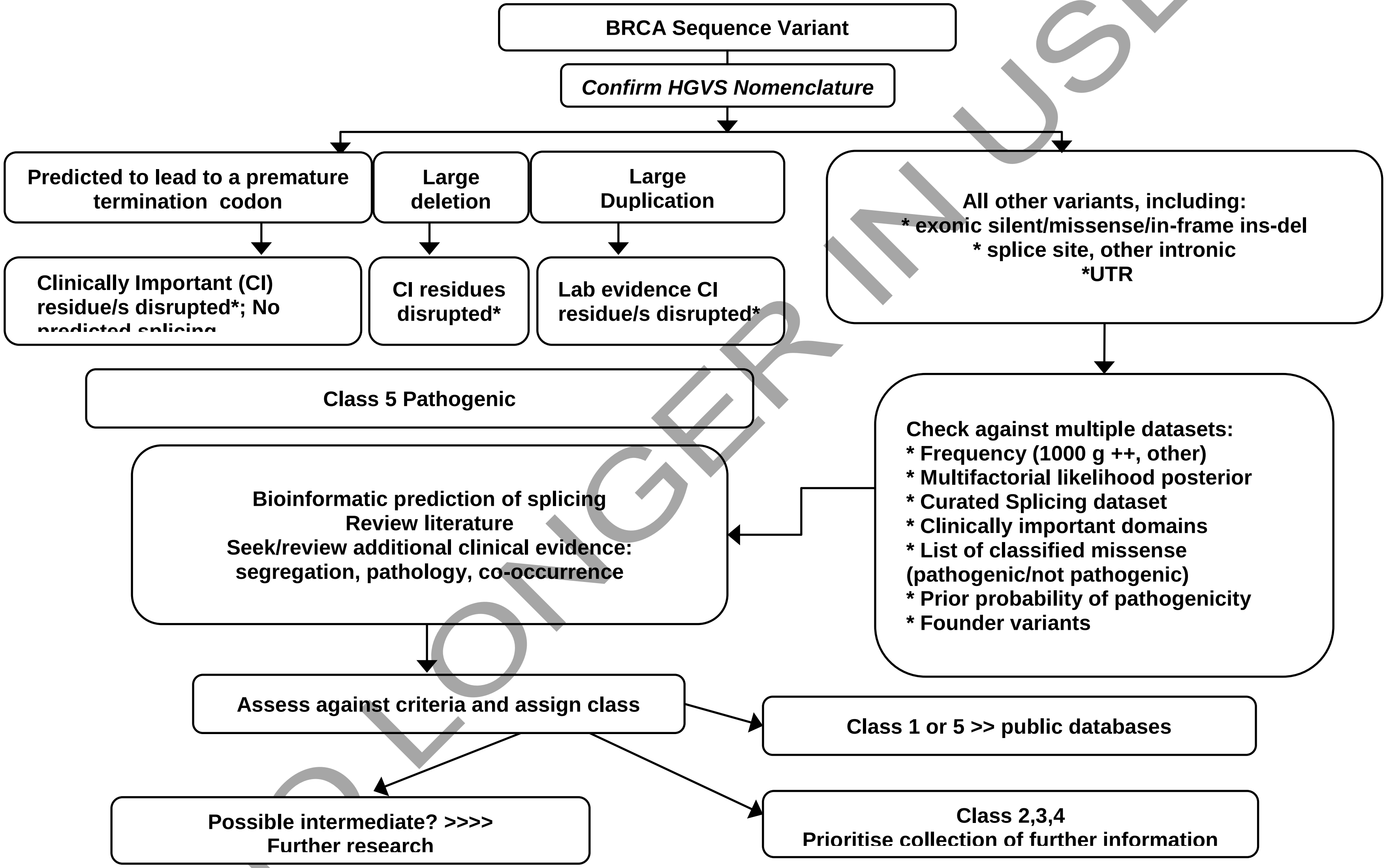

NO LONGER IN USE

**Figure 2: Classification of assumed premature termination variants incorporating bioinformatic prediction of effect on splicing.**

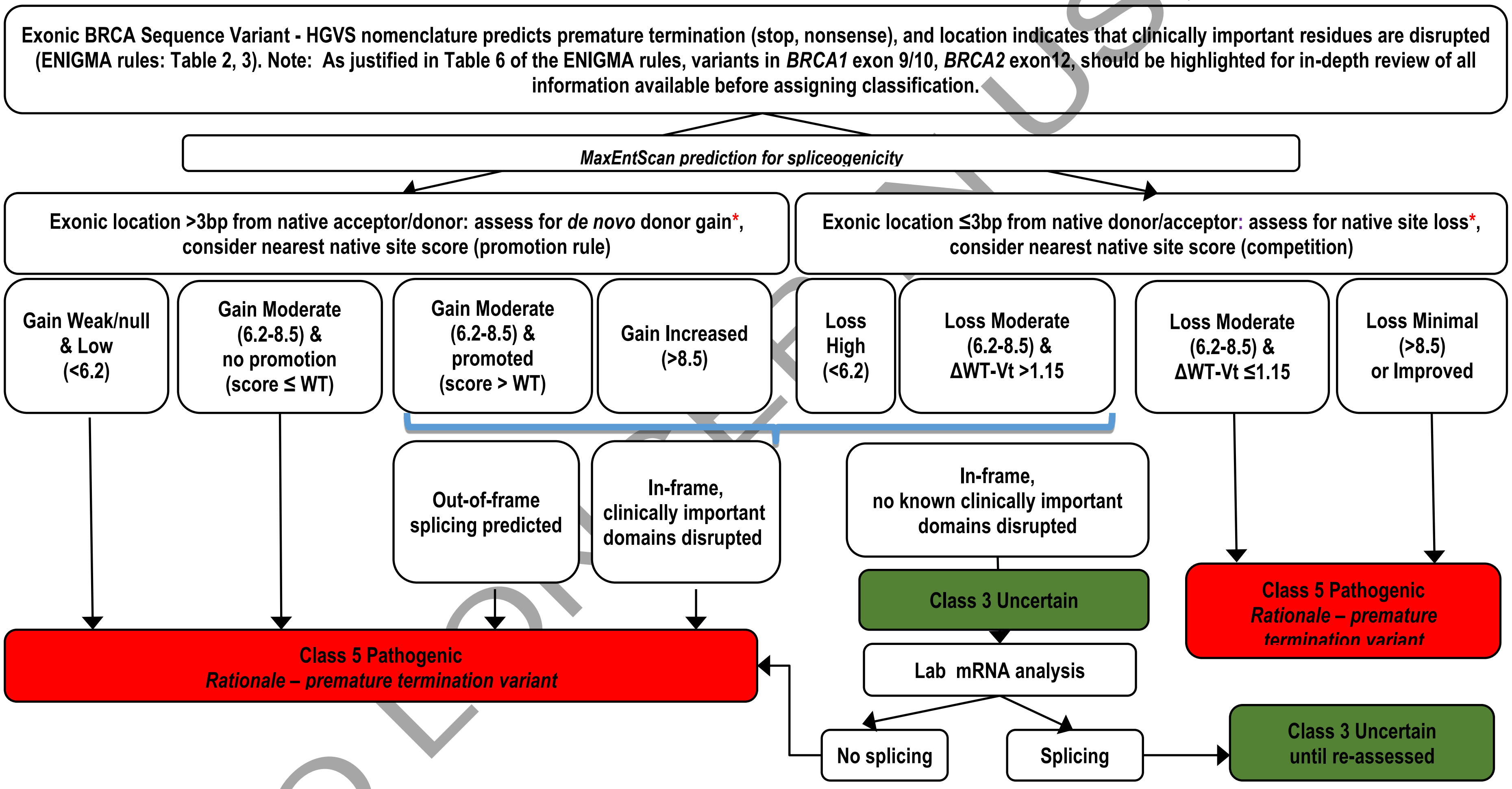

*** Raw score cutpoints are the same but have different annotations for *de novo site* gain versus native site loss. When using the Alamut platform, low scores may be returned as 0 or null. WT=wildtype, Vt = variant.**

NO LONGER IN USE

DOMCHEK, S. M., TANG, J., STOPFER, J., LILLI, D. R., HAMEL, N., TISCHKOWITZ, M., MONTEIRO, A. N., MESSICK, T. E., POWERS, J., YONKER, A., COUCH, F. J., GOLDGAR, D. E., DAVIDSON, H. R., NATHANSON, K. L., FOULKES, W. D. & GREENBERG, R. A.

NO LONGER IN USE

2013. Biallelic deleterious BRCA1 mutations in a woman with early-onset ovarian cancer. *Cancer Discov,* 3**,** 399-405.

DROST, R., BOUWMAN, P., ROTTENBERG, S., BOON, U., SCHUT, E., KLARENBEEK, S., KLIJN, C., VAN DER HEIJDEN, I., VAN DER GULDEN, H., WIENTJENS, E., PIETERSE, M., CATTEAU, A., GREEN, P., SOLOMON, E., MORRIS, J. R. & JONKERS, J. 2011. BRCA1 RING function is essential for tumor suppression but dispensable for therapy resistance. *Cancer Cell,* 20**,** 797-809.

DROST, R., DHILLON, K. K., VAN DER GULDEN, H., VAN DER HEIJDEN, I., BRANDSMA, I., CRUZ, C., CHONDRONASIOU, D., CASTROVIEJO-BERMEJO, M., BOON, U., SCHUT, E., VAN DER BURG, E., WIENTJENS, E., PIETERSE, M., KLIJN, C., KLARENBEEK, S., LOAYZA-PUCH, F., ELKON, R., VAN DEEMTER, L., ROTTENBERG, S., VAN DE VEN, M., DEKKERS, D. H., DEMMERS, J. A., VAN GENT, D. C., AGAMI, R., BALMANA, J., SERRA, V., TANIGUCHI, T., BOUWMAN, P. & JONKERS, J. 2016. BRCA1185delAG tumors may acquire therapy resistance through expression of RING-less BRCA1. *J Clin Invest,* 126**,** 2903-18.

EASTON, D. F., DEFFENBAUGH, A. M., PRUSS, D., FRYE, C., WENSTRUP, R. J., ALLEN-BRADY, K., TAVTIGIAN, S. V., MONTEIRO, A. N., IVERSEN, E. S., COUCH, F. J. & GOLDGAR, D. E. 2007. A systematic genetic assessment of 1,433 sequence variants of unknown clinical significance in the BRCA1 and BRCA2 breast cancer-predisposition genes. *Am J Hum Genet,* 81**,** 873-83.

ESASHI, F., CHRIST, N., GANNON, J., LIU, Y., HUNT, T., JASIN, M. & WEST, S. C. 2005. CDK-dependent phosphorylation of BRCA2 as a regulatory mechanism for recombinational repair. *Nature,* 434**,** 598-604.

ESASHI, F., GALKIN, V. E., YU, X., EGELMAN, E. H. & WEST, S. C. 2007. Stabilization of RAD51 nucleoprotein filaments by the C-terminal region of BRCA2. *Nat Struct Mol Biol,* 14**,** 468-74.

FACKENTHAL, J. D., YOSHIMATSU, T., ZHANG, B., DE GARIBAY, G. R., COLOMBO, M., DE VECCHI, G., AYOUB, S. C., LAL, K., OLOPADE, O. I., VEGA, A., SANTAMARINA, M., BLANCO, A., WAPPENSCHMIDT, B., BECKER, A., HOUDAYER, C., WALKER, L. C., LOPEZ-PEROLIO, I., THOMASSEN, M., PARSONS, M., WHILEY, P., BLOK, M. J., BRANDAO, R. D., TSERPELIS, D., BARALLE, D., MONTALBAN, G., GUTIERREZ-ENRIQUEZ, S., DIEZ, O., LAZARO, C., KCONFA, B. I., SPURDLE, A. B., RADICE, P. & DE LA HOYA, M. 2016. Naturally occurring BRCA2 alternative mRNA splicing events in clinically relevant samples. *J Med Genet*.

FARRUGIA, D. J., AGARWAL, M. K., PANKRATZ, V. S., DEFFENBAUGH, A. M., PRUSS, D., FRYE, C., WADUM, L., JOHNSON, K., MENTLICK, J., TAVTIGIAN, S. V., GOLDGAR, D. E. & COUCH, F. J. 2008. Functional assays for classification of BRCA2 variants of uncertain significance. *Cancer Res,* 68**,** 3523-31.

GAILDRAT, P., KRIEGER, S., DI GIACOMO, D., ABDAT, J., REVILLION, F., CAPUTO, S., VAUR, D., JAMARD, E., BOHERS, E., LEDEMENEY, D., PEYRAT, J. P., HOUDAYER, C., ROULEAU, E., LIDEREAU, R., FREBOURG, T., HARDOUIN, A., TOSI, M. & MARTINS, A. 2012. Multiple sequence variants of BRCA2 exon 7 alter splicing regulation. *J Med Genet,* 49**,** 609-17.

GOLDGAR, D. E., EASTON, D. F., BYRNES, G. B., SPURDLE, A. B., IVERSEN, E. S. & GREENBLATT, M. S. 2008. Genetic evidence and integration of various data sources for classifying uncertain variants into a single model. *Hum Mutat,* 29**,** 1265-72.
NO LONGER IN USE

GOLDGAR, D. E., EASTON, D. F., DEFFENBAUGH, A. M., MONTEIRO, A. N., TAVTIGIAN, S. V. & COUCH, F. J. 2004. Integrated evaluation of DNA sequence variants of unknown clinical significance: application to BRCA1 and BRCA2. *Am J Hum Genet,* 75**,** 535-44.

GUIDUGLI, L., CARREIRA, A., CAPUTO, S. M., EHLEN, A., GALLI, A., MONTEIRO, A. N., NEUHAUSEN, S. L., HANSEN, T. V., COUCH, F. J., VREESWIJK, M. P. & CONSORTIUM, E. 2014. Functional assays for analysis of variants of uncertain significance in BRCA2. *Hum Mutat,* 35**,** 151-64.

HAYES, F., CAYANAN, C., BARILLA, D. & MONTEIRO, A. N. 2000. Functional assay for BRCA1: mutagenesis of the COOH-terminal region reveals critical residues for transcription activation. *Cancer Res,* 60**,** 2411-8.

HOUDAYER, C., CAUX-MONCOUTIER, V., KRIEGER, S., BARROIS, M., BONNET, F., BOURDON, V., BRONNER, M., BUISSON, M., COULET, F., GAILDRAT, P., LEFOL, C., LEONE, M., MAZOYER, S., MULLER, D., REMENIERAS, A., REVILLION, F., ROULEAU, E., SOKOLOWSKA, J., VERT, J. P., LIDEREAU, R., SOUBRIER, F., SOBOL, H., SEVENET, N., BRESSAC-DE PAILLERETS, B., HARDOUIN, A., TOSI, M., SINILNIKOVA, O. M. & STOPPA-LYONNET, D. 2012. Guidelines for splicing analysis in molecular diagnosis derived from a set of 327 combined in silico/in vitro studies on BRCA1 and BRCA2 variants. *Hum Mutat,* 33**,** 1228-38.

HU, Y. F., MIYAKE, T., YE, Q. & LI, R. 2000. Characterization of a novel trans-activation domain of BRCA1 that functions in concert with the BRCA1 C-terminal (BRCT) domain. *J Biol Chem,* 275**,** 40910-5.

JIAN, X., BOERWINKLE, E. & LIU, X. 2014. In silico prediction of splice-altering single nucleotide variants in the human genome. *Nucleic Acids Res,* 42**,** 13534-44.

LEE, M. S., GREEN, R., MARSILLAC, S. M., COQUELLE, N., WILLIAMS, R. S., YEUNG, T., FOO, D., HAU, D. D., HUI, B., MONTEIRO, A. N. & GLOVER, J. N. 2010. Comprehensive analysis of missense variations in the BRCT domain of BRCA1 by structural and functional assays. *Cancer Res,* 70**,** 4880-90.

LI, L., BISWAS, K., HABIB, L. A., KUZNETSOV, S. G., HAMEL, N., KIRCHHOFF, T., WONG, N., ARMEL, S., CHONG, G., NAROD, S. A., CLAES, K., OFFIT, K., ROBSON, M. E., STAUFFER, S., SHARAN, S. K. & FOULKES, W. D. 2009. Functional redundancy of exon 12 of BRCA2 revealed by a comprehensive analysis of the c.6853A>G (p.I2285V) variant. *Hum Mutat,* 30**,** 1543-50.

MEEKS, H. D., SONG, H., MICHAILIDOU, K., BOLLA, M. K., DENNIS, J., WANG, Q., BARROWDALE, D., FROST, D., EMBRACE, MCGUFFOG, L., ELLIS, S., FENG, B., BUYS, S. S., HOPPER, J. L., SOUTHEY, M. C., TESORIERO, A., KCONFAB, I., JAMES, P. A., BRUINSMA, F., CAMPBELL, I. G., AUSTRALIA OVARIAN CANCER STUDY, G., BROEKS, A., SCHMIDT, M. K., HOGERVORST, F. B., HEBON, BECKMAN, M. W., FASCHING, P. A., FLETCHER, O., JOHNSON, N., SAWYER, E. J., RIBOLI, E., BANERJEE, S., MENON, U., TOMLINSON, I., BURWINKEL, B., HAMANN, U., MARME, F., RUDOLPH, A., JANAVICIUS, R., TIHOMIROVA, L., TUNG, N., GARBER, J., CRAMER, D., TERRY, K. L., POOLE, E. M., TWOROGER, S. S., DORFLING, C. M., VAN RENSBURG, E. J., GODWIN, A. K., GUENEL, P., TRUONG, T., COLLABORATORS, G. S., STOPPA-LYONNET, D., DAMIOLA, F., MAZOYER, S., SINILNIKOVA, O. M., ISAACS, C., MAUGARD, C., BOJESEN, S. E., FLYGER, H., GERDES, A. M., HANSEN, T. V., JENSEN, A., KJAER, S. K., HOGDALL, C., HOGDALL, E., PEDERSEN, I. S., THOMASSEN, M., BENITEZ, J., GONZALEZ-NEIRA, A., OSORIO, A., HOYA MDE, L., SEGURA, P. P., DIEZ, O., LAZARO, C., BRUNET, J., ANTON-CULVER, H., EUNJUNG, L., JOHN, E. M., NEUHAUSEN, S. L., DING, Y. C., CASTILLO, D., WEITZEL, J. N., GANZ, P. A., NUSSBAUM, R. L., CHAN, S. B., KARLAN, B. Y., LESTER, J., WU, A., GAYTHER, S.,
NO LONGER IN USE

RAMUS, S. J., SIEH, W., WHITTERMORE, A. S., MONTEIRO, A. N., PHELAN, C. M., TERRY, M. B., PIEDMONTE, M., OFFIT, K., ROBSON, M., LEVINE, D., et al. 2016. BRCA2 Polymorphic Stop Codon K3326X and the Risk of Breast, Prostate, and Ovarian Cancers. *J Natl Cancer Inst,* 108.

OLIVER, A. W., SWIFT, S., LORD, C. J., ASHWORTH, A. & PEARL, L. H. 2009. Structural basis for recruitment of BRCA2 by PALB2. *EMBO Rep,* 10**,** 990-6.

PARSONS, M. T., WHILEY, P. J., BEESLEY, J., DROST, M., DE WIND, N., THOMPSON, B. A., MARQUART, L., HOPPER, J. L., JENKINS, M. A., AUSTRALASIAN COLORECTAL CANCER FAMILY, R., BROWN, M. A., TUCKER, K., WARWICK, L., BUCHANAN, D. D. & SPURDLE, A. B. 2015. Consequences of germline variation disrupting the constitutional translational initiation codon start sites of MLH1 and BRCA2: Use of potential alternative start sites and implications for predicting variant pathogenicity. *Mol Carcinog,* 54**,** 513-22.

PLON, S. E., ECCLES, D. M., EASTON, D., FOULKES, W. D., GENUARDI, M., GREENBLATT, M. S., HOGERVORST, F. B., HOOGERBRUGGE, N., SPURDLE, A. B., TAVTIGIAN, S. V. & GROUP, I. U. G. V. W. 2008. Sequence variant classification and reporting: recommendations for improving the interpretation of cancer susceptibility genetic test results. *Hum Mutat,* 29**,** 1282-91.

RICHARDS, C. S., BALE, S., BELLISSIMO, D. B., DAS, S., GRODY, W. W., HEGDE, M. R., LYON, E., WARD, B. E. & MOLECULAR SUBCOMMITTEE OF THE, A. L. Q. A. C. 2008. ACMG recommendations for standards for interpretation and reporting of sequence variations: Revisions 2007. *Genet Med,* 10**,** 294-300.

RODRIGUEZ, J. A. & HENDERSON, B. R. 2000. Identification of a functional nuclear export sequence in BRCA1. *J Biol Chem,* 275**,** 38589-96.

ROSENTHAL, E. T., BOWLES, K. R., PRUSS, D., VAN KAN, A., VAIL, P. J., MCELROY, H. & WENSTRUP, R. J. 2015. Exceptions to the rule: case studies in the prediction of pathogenicity for genetic variants in hereditary cancer genes. *Clin Genet,* 88**,** 533-41.

SANTOS, C., PEIXOTO, A., ROCHA, P., PINTO, P., BIZARRO, S., PINHEIRO, M., PINTO, C., HENRIQUE, R. & TEIXEIRA, M. R. 2014. Pathogenicity evaluation of BRCA1 and BRCA2 unclassified variants identified in Portuguese breast/ovarian cancer families. *J Mol Diagn,* 16**,** 324-34.

SAWYER, S. L., TIAN, L., KAHKONEN, M., SCHWARTZENTRUBER, J., KIRCHER, M., MAJEWSKI, J., DYMENT, D. A., INNES, A. M., BOYCOTT, K. M., MOREAU, L. A., MOILANEN, J. S., GREENBERG, R. A. & UNIVERSITY OF WASHINGTON CENTRE FOR MENDELIAN, G. 2015. Biallelic Mutations in BRCA1 Cause a New Fanconi Anemia Subtype. *Cancer Discov,* 5**,** 135-42.

SPURDLE, A. B., WHILEY, P. J., THOMPSON, B., FENG, B., HEALEY, S., BROWN, M. A., PETTIGREW, C., KCONFAB, VAN ASPEREN, C. J., AUSEMS, M. G., KATTENTIDT-MOURAVIEVA, A. A., VAN DEN OUWELAND, A. M., DUTCH BELGIUM, U. V. C., LINDBLOM, A., PIGG, M. H., SCHMUTZLER, R. K., ENGEL, C., MEINDL, A., GERMAN CONSORTIUM OF HEREDITARY, B., OVARIAN, C., CAPUTO, S., SINILNIKOVA, O. M., LIDEREAU, R., FRENCH, C. G. C., COUCH, F. J., GUIDUGLI, L., HANSEN, T., THOMASSEN, M., ECCLES, D. M., TUCKER, K., BENITEZ, J., DOMCHEK, S. M., TOLAND, A. E., VAN RENSBURG, E. J., WAPPENSCHMIDT, B., BORG, A., VREESWIJK, M. P., GOLDGAR, D. E. & CONSORTIUM, E. 2012. BRCA1 R1699Q variant displaying ambiguous functional abrogation confers intermediate breast and ovarian cancer risk. *J Med Genet,* 49**,** 525-32.
NO LONGER IN USE

STARITA, L. M., YOUNG, D. L., ISLAM, M., KITZMAN, J. O., GULLINGSRUD, J., HAUSE, R. J., FOWLER, D. M., PARVIN, J. D., SHENDURE, J. & FIELDS, S. 2015. Massively Parallel Functional Analysis of BRCA1 RING Domain Variants. *Genetics,* 200**,** 413-22.

TAVTIGIAN, S. V., GREENBLATT, M. S., LESUEUR, F., BYRNES, G. B. & GROUP, I. U. G. V. W. 2008. In silico analysis of missense substitutions using sequence-alignment based methods. *Hum Mutat,* 29**,** 1327-36.

THAKUR, S., ZHANG, H. B., PENG, Y., LE, H., CARROLL, B., WARD, T., YAO, J., FARID, L. M., COUCH, F. J., WILSON, R. B. & WEBER, B. L. 1997. Localization of BRCA1 and a splice variant identifies the nuclear localization signal. *Mol Cell Biol,* 17**,** 444-52.

THOMPSON, B. A., SPURDLE, A. B., PLAZZER, J. P., GREENBLATT, M. S., AKAGI, K., AL-MULLA, F., BAPAT, B., BERNSTEIN, I., CAPELLA, G., DEN DUNNEN, J. T., DU SART, D., FABRE, A., FARRELL, M. P., FARRINGTON, S. M., FRAYLING, I. M., FREBOURG, T., GOLDGAR, D. E., HEINEN, C. D., HOLINSKI-FEDER, E., KOHONEN-CORISH, M., ROBINSON, K. L., LEUNG, S. Y., MARTINS, A., MOLLER, P., MORAK, M., NYSTROM, M., PELTOMAKI, P., PINEDA, M., QI, M., RAMESAR, R., RASMUSSEN, L. J., ROYER-POKORA, B., SCOTT, R. J., SIJMONS, R., TAVTIGIAN, S. V., TOPS, C. M., WEBER, T., WIJNEN, J., WOODS, M. O., MACRAE, F., GENUARDI, M. & INSIGHT 2014. Application of a 5-tiered scheme for standardized classification of 2,360 unique mismatch repair gene variants in the InSiGHT locus-specific database. *Nat Genet,* 46**,** 107-15.

VALLEE, M. P., SERA, T. L., NIX, D. A., PAQUETTE, A. M., PARSONS, M. T., BELL, R., HOFFMAN, A., HOGERVORST, F. B., GOLDGAR, D. E., SPURDLE, A. B. & TAVTIGIAN, S. V. 2016. Adding In Silico Assessment of Potential Splice Aberration to the Integrated Evaluation of BRCA Gene Unclassified Variants. *Hum Mutat*.

WALKER, L. C., WHILEY, P. J., COUCH, F. J., FARRUGIA, D. J., HEALEY, S., ECCLES, D. M., LIN, F., BUTLER, S. A., GOFF, S. A., THOMPSON, B. A., LAKHANI, S. R., DA SILVA, L. M., KCONFAB, I., TAVTIGIAN, S. V., GOLDGAR, D. E., BROWN, M. A. & SPURDLE, A. B. 2010. Detection of splicing aberrations caused by BRCA1 and BRCA2 sequence variants encoding missense substitutions: implications for prediction of pathogenicity. *Hum Mutat,* 31**,** E1484-505.

WALKER, L. C., WHILEY, P. J., HOUDAYER, C., HANSEN, T. V., VEGA, A., SANTAMARINA, M., BLANCO, A., FACHAL, L., SOUTHEY, M. C., LAFFERTY, A., COLOMBO, M., DE VECCHI, G., RADICE, P., SPURDLE, A. B. & CONSORTIUM, E. 2013. Evaluation of a 5-tier scheme proposed for classification of sequence variants using bioinformatic and splicing assay data: inter-reviewer variability and promotion of minimum reporting guidelines. *Hum Mutat,* 34**,** 1424-31.

WAPPENSCHMIDT, B., BECKER, A. A., HAUKE, J., WEBER, U., ENGERT, S., KOHLER, J., KAST, K., ARNOLD, N., RHIEM, K., HAHNEN, E., MEINDL, A. & SCHMUTZLER, R. K. 2012. Analysis of 30 putative BRCA1 splicing mutations in hereditary breast and ovarian cancer families identifies exonic splice site mutations that escape in silico prediction. *PLoS One,* 7**,** e50800.

WHILEY, P. J., PARSONS, M. T., LEARY, J., TUCKER, K., WARWICK, L., DOPITA, B., THORNE, H., LAKHANI, S. R., GOLDGAR, D. E., BROWN, M. A. & SPURDLE, A. B. 2014. Multifactorial likelihood assessment of BRCA1 and BRCA2 missense variants confirms that BRCA1:c.122A>G(p.His41Arg) is a pathogenic mutation. *PLoS One,* 9**,** e86836.

XIA, B., SHENG, Q., NAKANISHI, K., OHASHI, A., WU, J., CHRIST, N., LIU, X., JASIN, M., COUCH, F. J. & LIVINGSTON, D. M. 2006. Control of BRCA2 cellular and clinical functions by a nuclear partner, PALB2. *Mol Cell,* 22**,** 719-29.
NO LONGER IN USE
