## Supplemental Section 3 for "Evidence-based recommendations for gene-specific ACMG/AMP variant classification from the ClinGen ENIGMA BRCA1 and BRCA2 Variant Curation Expert Panel": CSpec_BRCA12ACMG-Rules-Appendices_V1.0_2023-04-27.docx

Appendices-*BRCA1* & *BRCA2 ACMG/AMP rules specifications, V1.0 2023-04-27.*

### Appendix A: Assignment of code weights based on odds of pathogenicity

Designation of likelihood ratios (LRs) to ACMG/AMP rule code strengths were based on LR ranges recently proposed as consistent with ACMG/AMP qualitative rule strengths for future classification in a Bayesian framework (Tavtigian et al 2018, PMID: 29300386). The benign category intervals are calculated as inverse odds to the pathogenic category intervals. These odds ranges assume a global prior probability of pathogenicity of 0.10.

##### Table 1: Pathogenic and Benign code strength assignment based on measured odds in favour of pathogenicity

| **Code Strength** | **Odds range for Pathogenic codes** | **Odds range for Benign Codes** |
| --- | --- | --- |
| Strong | >18.70 to 350 | <0.05 to 0.00285 |
| Moderate | >4.30 to 18.70 | <0.23 to 0.05 |
| Supporting | >2.08 to 4.30 | <0.48 to 0.23 |
| No evidence | 1.00 to 2.08 | 0.48 to 1.00 |

### Appendix B: Multifactorial likelihood analysis methodology and assignment of ACMG/AMP code weights for expanded set of evidence types

The current iteration of multifactorial likelihood model for *BRCA1/2* variant interpretation used by the ENIGMA *BRCA1/2* expert panel allows for inclusion of likelihood ratios (LRs) for pathogenicity estimated from clinical data, such as co-segregation with disease, co-occurrence with a pathogenic variant in the same gene, reported family history, breast tumor pathology, and case-control data (Goldgar et al 2004, PMID:15290653; Thompson et al 2005, PMID:12900794; Easton et al 2007, PMID:17924331; Spurdle et al 2014, PMID:25857409; de la Hoya et al 2016, PMID: 27008870; Parsons et al 2019, PMID:31131967; Li et al 2020, PMID: 31853058). To date, LRs have been combined with a clinically calibrated prior probability of pathogenicity based on bioinformatic predictions of variant effect on protein sequence or mRNA splicing (Tavtigian et al 2008, PMID: 8972225; Vallee et al 2016, PMID: 26913838) to assign class based on posterior probability. The following caveats and recommendations are noted in relation to multifactorial likelihood analysis methodology. Due to selection of the reference sets used to derive LR estimates and prior probabilities, the model is designed to assess if a variant demonstrates the clinical features observed for a classical “high risk” variant. Only independent lines of evidence should be included. Tumour pathology information cannot be considered if the individual was selected for testing based only on that phenotype.

Given the breadth of data types included in multifactorial likelihood analysis, not all of which are captured in the current ACMG/AMP scheme (in particular benign codes), evidence weights for clinical data derived from calibrated multifactorial likelihood analysis approaches will be assigned using PP4 (combined clinical evidence towards pathogenicity) or BP5 (combined evidence against pathogenicity), with weight defined by combined LR as in Table 1. This approach is consistent with the recently described points system approach (Tavtigian et al 2020, PMID: 32720330). [Bioinformatic prior probabilities will **not be considered**, since bioinformatic data is captured under other ACMG/AMP codes]. These codes may be applied for both previously published data, and also unpublished data where there is no appropriate ACMG/AMP code. See Table 2 for examples of code assignment.

##### Table 2: Examples of ACMG/AMP code assignment based on multifactorial likelihood clinical data points

| **Example Variant** | **Previously Published** | **Source(s)** | **Co-Segregation LR** | **Tumour Pathology LR** | **Co-occurrence LR** | **Family History LR** | **Combined Clinical LR for PP4 or BP5** ^a^ | **Code/s to apply** | **Text Description** |
| --- | --- | --- | --- | --- | --- | --- | --- | --- | --- |
| 1 | Yes | Easton et al 2007 | - | - | 2.231 | 1508.137 | 3364.654 | PP4_Very Strong | PP4_Very Strong: Published clinical data for multifactorial likelihood analysis (PMID: 17924331); Combined LR for clinical data indicative of very strong evidence for pathogenicity (LR 3364) based on co-occurrence (LR 2.23), family history (LR 1508). |
|  | No | VCEP | 1.961 | - | - | - | - | PP1 not met | PP1 not met: co-segregation LR 1.96 (1 family, internal VCEP contributor). |
| 2 | Yes | Parsons et al 2019, Li et al 2020 | 730.630 | 2.406 | - | 1.556 | 2735.286 | PP4_Very Strong | PP4_Very Strong: Published clinical data for multifactorial likelihood analysis (PMID: 31131967, 31853058); Combined LR for clinical data indicative of very strong evidence towards pathogenicity (LR 2735) based on co-segregation (LR 730.63, 12 families), Tumour pathology (LR 2.40, 6 tumours), family history (LR 1.56). |
|  | No | VCEP | 5.240 | - | - | - | - | PP1_  Moderate | PP1_mod: co-segregation LR 5.24 (1 family, internal VCEP contributor). |
| 3 | No | ENIGMA unpublished | - | 0.2138 | 1.102 | 2.362 | 0.557 | PP4/BP5 not met | PP4 and BP5 not met: Unpublished clinical data for multifactorial likelihood analysis (drawn from datasets used for PMID: 17924331, and PMID 31853058); Combined LR 0.557 based on Tumour pathology (LR 0.214), co-occurrence (LR 1.102), family history (LR 2.362). |
|  | No | VCEP | 74321 | - | - | - | - | PP1_Very Strong | PP1_Very Strong: co-segregation LR 74321 (10 families, internal VCEP contributors). |
| 4 | None | N/A | - | - | - | - | - | PP4/BP5 not met | PP4 and BP5 not met: no data. |
|  | No | VCEP | 14.421 |  |  |  |  | PP1_  Moderate | PP1_Moderate: co-segregation LR 14.42 (analysis 2 families, internal VCEP contributors). |
| 5 | Yes | Parsons et al 2019 |  |  | 1.177 | 0.0127 | 0.015 | BP5_Strong | BP5_Strong: Published clinical data for multifactorial likelihood analysis (PMID: 31131967); Combined LR for clinical data indicative of strong evidence towards benign (LR 0.015) based on o-occurrence (LR 1.177), family history (LR 0.0127). |
|  | No | VCEP | 0.234 |  |  |  |  | BS4_  Supporting | BS4_Supporting: co-segregation LR 0.234 (1 family, internal VCEP contributors). |

^a^ Combined clinical LR is calculated by multiplying individual LRs e.g. 2.231x 1508.137. Internal co-segregation data from VCEP contributors is captured under PP1 or BS4, whereas internal tumour pathology data is combined with other clinical LRs. Weights are assigned as per Tavtigian et al 2018 (PMID: 29300386). For these examples, data and codes drawn from internal VCEP contributions are denoted in blue font.

### Appendix C: *BRCA1* and *BRCA2* exon maps and functional domains

BRCA1 and BRCA2 exon maps are used to define **Single-cassette and Multi-Cassette exon losses, and 10% protein loss calculation.**

##### Figure 1: *BRCA1* Exon map.

Coding DNA reference sequence from genomic refseq [NG_005905.2](http://www.ncbi.nlm.nih.gov/nuccore/NG_005905.2), covering *BRCA1* transcript [NM_007294.4](http://www.ncbi.nlm.nih.gov/nuccore/NM_007294.4). Exons are sequentially numbered to match the exon descriptions of the MANE transcript (NM_007294.4). Exon numbering of *BRCA1* has historically been according to GenBank [U14680.1](http://www.ncbi.nlm.nih.gov/nuccore/U14680), with exon 4 missing due to a correction made after the initial description of the gene, termed in this document as legacy exon numbering. Nucleotide (nt) position relative to cDNA (c.), and amino acid position for the encoded protein (p.) for the start of each exon are noted above the exon (shown as a box). The lines between each exon represent the frame at which the exon ends. Forward slash: a two-nt overhang, Backslash: a one-nt overhang, Vertical bar: in-frame end of the exon (i.e. the last base of the exon is the third base of a codon). If the lines on both sides of an exon are parallel, deletion of this exon would be in-frame. See figure key for more explanation. Table 3 provides more details regarding proven and potentially clinically important residues within functional domains.

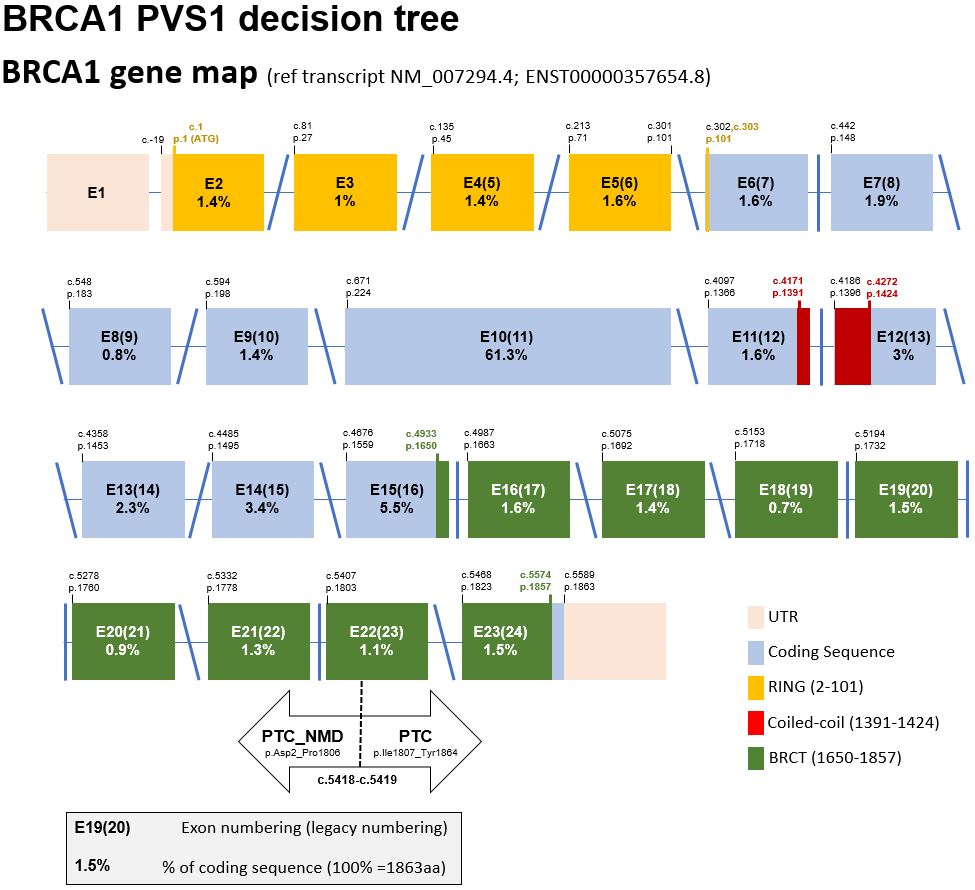

##### Figure 2: *BRCA2* Exon map.

Coding DNA reference sequence from genomic refseq [NG_012772.3](http://www.ncbi.nlm.nih.gov/nuccore/NG_012772.3) covering *BRCA2* transcript [NM_000059.4](https://www.ncbi.nlm.nih.gov/nuccore/NM_000059.4). Nucleotide (nt) position relative to cDNA (c.), and amino acid position for the encoded protein (p.) for the start of each exon are noted above the exon (shown as a box). The lines between each exon represent the frame at which the exon ends. Forward slash: a two-nt overhang, Backslash: a one-nt overhang, Vertical bar: in-frame end of the exon (i.e. the last base of the exon is the third base of a codon). If the lines on both sides of an exon are parallel, deletion of this exon would be in-frame. See figure key for more explanation. Table 4 provides more details regarding proven clinically important residues within functional domains.

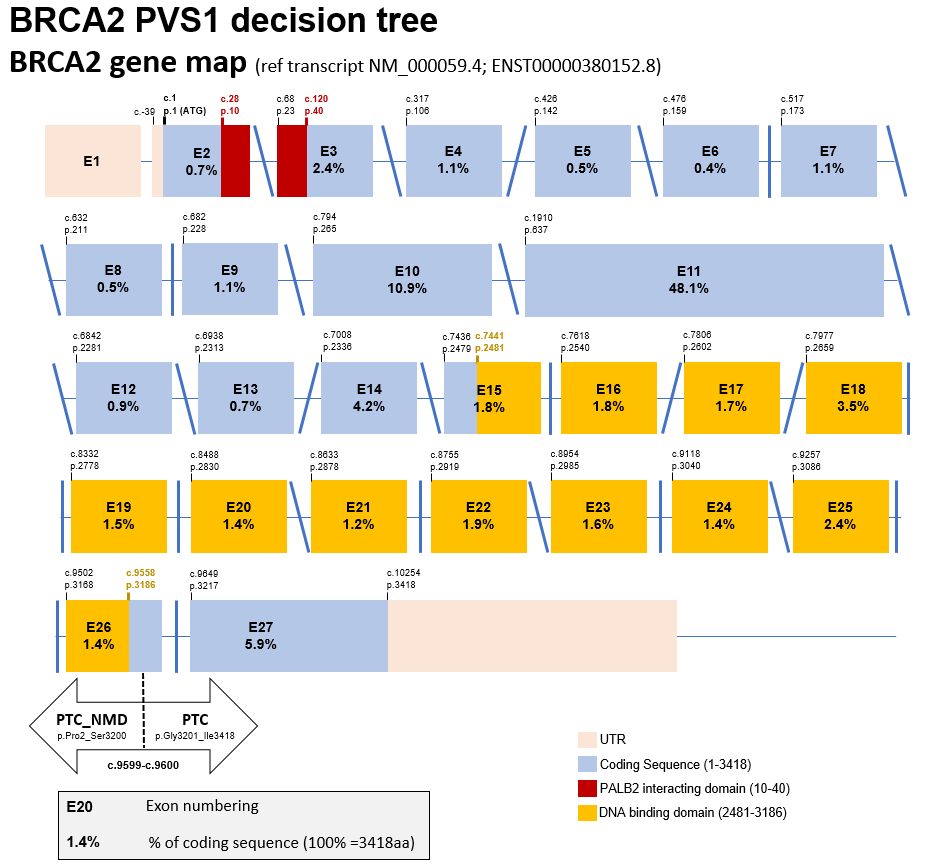

#### Clinically important residues and functional protein domains of BRCA1 and BRCA2:

Clinically important residues are defined by the location of sequence variants that introduce deleterious changes to protein function (via missense alteration, protein sequence deletion, or protein truncation in the last exon), AND association with high cancer risk. A clinically important functional protein domain is a recognized protein functional domain reported to harbor one or more clinically important residues.

##### Table 3: Catalogue of BRCA1 conserved domains/motifs and currently known clinically important amino acid residues, and relevance for classification of *BRCA1* in-frame and terminal exon sequence variants.

| **Domain/**  **Motif** | **AA start** | **AA end** | **AA alterations with Demonstrated Clinical Importance^a^** | **References and summary interpretation^a^** |
| --- | --- | --- | --- | --- |
| RING | 2 | 101 | M18T (c.53T>C (p.Met18Thr))  L22S (c.65T>C (p.Leu22Ser))  T37K (c.110C>A (p.Thr37Lys))  C39R (c.115T>C (p.Cys39Arg))  C39Y (c.116G>A (p.Cys39Tyr))  H41R (c.122A>G (p.His41Arg))  C44S (c.130T>A (p.Cys44Ser))  C44Y (c.131G>A (p.Cys44Tyr))  C44F (c.131G>T (p.Cys44Phe))  C47Y (c.140G>A (p.Cys47Tyr))  C61G (c.181T>G (p.Cys61Gly))  C61S (c.181T>A (p.Cys61Ser))  C64Y (c.191G>A (p.Cys64Tyr))  T77_F79del (c.230_238del (p.Thr77_Phe79del)) | http://www.ncbi.nlm.nih.gov/protein/15988069;  http://hci-exlovd.hci.utah.edu; Multifactorial analysis for H41R (c.122A>G (p.His41Arg)) (Whiley et al 2014, PMID 24489791).  Intiation codon changes excluded since these reflect a different mechanism of action. |
| NES | 81 | 99 | None reported | Domain location description (Rodriquez and Henderson, 2000, PMID: 10991937). |
| NLS1 | 503 | 508 | None reported | Domain location description (Chen et al 1996, PMID 8955125; Thakur et al 1997, PMID: 8972225). |
| NLS2 | 607 | 614 | None reported | Domain location description (Chen et al 1996, PMID 8955125). |
| NLS3 | 651 | 656 | None reported | Domain location description (Chen et al 1996, PMID 8955125). |
| COILED-COIL | 1391 | 1424 | Domain considered potentially clinically important. | Domain location description (Hu et al 2000, PMID: 11067843). Note, non-hydrophobic changes at key positions in the alpha-helix (M1400T/R/K;L1404P/R/Q; L1407P,R,H; M1411T,R,K; L1414P,R,Q; and L1418S and possibly H1421), and other proline substitutions in the alpha-helix (Q1401P, H1420P, Q1408P, Q1409P, A1412P, and A1416P), might be expected to impact BRCA1 interaction with PALB2 - and potentially be risk-associated. Missense substitutions M1411T and L1407P have been demonstrated to show partial impact on protein function (Woods et al 2016. PMID: 28781887). The *BRCA1* c.4231T>C M1411T substitution has been identified in multiple probands clinically ascertained for testing due to reported breast-ovarian cancer family history. Although no variants in the Coiled-Coil domain are considered formally classified as Pathogenic, given the suspicion of cancer risk association for at least some missense substitutions, it is termed a potentially clinically important functional domain. |
| BRCT DOMAINS | 1650 | 1863 | T1685A (c.5053A>G (p.Thr1685Ala))  T1685I (c.5054C>T (p.Thr1685Ile))  V1688del (c.5062_5064del (p.Val1688del))  C1697R (c.5089T>C (p.Cys1697Arg))  R1699W (c.5095C>T (p.Arg1699Trp))  G1706E (c.5117G>A (p.Gly1706Glu))  S1715R (c.5143A>C (p.Ser1715Arg))  S1715N (c.5144G>A (p.Ser1715Asn))  V1736A (c.5207T>C (p.Val1736Ala))  V1736G (c.5207T>G (p.Val1736Gly))  G1738R (c.5212G>A (p.Gly1738Arg))  G1738E (c.5213G>A (p.Gly1738Glu))  D1739V (c.5216A>T (p.Asp1739Val))  G1748D (c.5243G>A (p.Gly1748Asp))  L1764P (c.5291T>C (p.Leu1764Pro))  I1766S (c.5297T>G (p.Ile1766Ser))  G1770V (c.5309G>T (p.Gly1770Val))  M1775K (c.5324T>A (p.Met1775Lys))  M1775R (c.5324T>G (p.Met1775Arg))  L1786P (c.5357T>C (p.Leu1786Pro))  C1787S/G1788V (c.[5359T>A; 5363G>T] (p.Cys1787_Gly1788delinsSerVal))  W1837R (c.5509T>C (p.Trp1837Arg))  V1838E (c.5513T>A (p.Val1838Glu))  L1839S (c.5516T>C (p.Leu1839Ser))  Y1853C (c.5558A>G (p.Tyr1853Cys)) | Digestion of a bacterially expressed BRCA1 C-terminal region (aa 1528-1863) resulted in a proteolytically stable fragment (aa 1646-1863), and further deletion of residues 1860 to 1863 yielded a soluble fragment used for structural determination, thus residue positions [1646 to 1859] can be considered as delimiting the start [aa1646] and the end [aa1859] of the tandem BRCT domains in BRCA1 (Williams et al 2001, PMID: 11573086).    Other observations further refine the start and end of the structural and functional unit of the tandem BRCT domains. The first secondary structure in the BRTC-N, a β -sheet (β1), starts at M1650 indicating that this residue position is the start [aa1650].  Y1853, L1854 and I1855 represent a cluster of hydrophobic residues present at the end of Motif II in several BRCT domains (Bork et al 1997, PMID: 9034168). Position 1854 is implicated as clinically important by the observation that Y1853X (c.5559C>G (p.Tyr1853Ter); it was identified in an early onset bilateral breast cancer case (Friedman et al 1994, PMID: 7894493). Additional data indicate that Q1857 is required for BRCT stability in mammalian cells (Fernandes et al 2019, PMID: 30765603).  We propose that for clinical variant annotation the BRCT domains in BRCA1 should be considered [1650-1853], with equivocal evidence for functional impact for variants that lead to a premature termination codon from 1855 to 1857 (Fernandes et al 2019, PMID: 30765603). A missense or nonsense variant predicted to truncate protein sequence at residues from position p.1858-onwards is considered highly unlikely to be clinically important.  However, the impact of variants that cause a frameshift before position 1855 but that may end after 1857 is not certain, and a hypomorphic effect cannot be excluded.  **In summary, the BRCT region considered (potentially) clinically important is denoted as 1650-1857.** |
| **^a^** Missense substitutions in denoted functional domains that are designated as Class 5 pathogenic based on multifactorial likelihood posterior probability of pathogenicity > 0.99 (listed in a web-based portal of published multifactorial likelihood studies - http://hci-exlovd.hci.utah.edu, or individual references noted), and for which there is no/little effect on mRNA transcript profile, ***unless*** the variant results in an aberrant transcript that encodes a discrete in-frame deletion considered informative to definition of clinically important domains. Note - The following pathogenic exonic “predicted missense” variants known to alter mRNA splicing have been excluded from Table 3 above, as justified below: Variant \| mRNA Change \| Predicted Protein Change \| Reason for exclusion; *BRCA1* R71G (c.211A>G (p.Arg71Gly)) \| r.[135_212del, 191_212del] \| p.[Lys45_Arg71del, Cys64Ter) \| Two alternate transcripts (out-of-frame and in-frame). Level of full length transcript not assessed (Houdayer et al 2012, PMID: 22505045; Menendez et al 2012, PMID: 21735045; Sanz et al 2010, PMID: 20215541; Vega et al 2001, PMID: 11385711); *BRCA1* R1495M (c.4484G>T (p.Arg1495Met)) \| r.[4358_4484del, 4358_4675del] \| p.(Ala1453GlyfsTer10) - predominant transcript \| Predominant alternate transcript is out-of-frame. Loss of function assumed due to loss of full length transcript from variant allele (Houdayer et al 2012, PMID: 22505045; Colombo et al 2013, PMID: 24569164; Santos et al 2014, PMID: 24607278).; *BRCA1* E1559K (c.4675G>A (p.Glu1559Lys)) \| r.[4665_4675del] \| p.(Gln1366AlafsTer13) \| Alternate transcript is out-of-frame. Level of full length transcript not assessed (Wappenschmidt et al 2012, PMID: 23239986).; *BRCA1* A1623G (c.4868C>G (p.Ala1623Gly)) \| r.[4868_4986del] \| p.(Ala1623AspfsTer16) \| Alternate transcript is out-of-frame. Variant allele produces some full length transcript (Walker et al 2010, PMID: 23893897); *BRCA1* D1692N (c.5074G>A (p.Asp1692Asn)) \| r.[4987_5074del, 5074_5075ins5074+1_5074+153] \| p.(Val1665SerfsTer8) – predominant transcript \| Predominant alternate transcript, based on minigene assay (Ahlborn et al 2015, PMID: 25724305), is out of frame.; *BRCA1* D1692N (c.5074G>C (p.Asp1692Asn)) \| r.[4987_5074del, 5074_5075ins5074+1_5074+153] \| p.(Val1665SerfsTer8) – predominant transcript \| Predominant alternate transcript, based on minigene assay (Ahlborn et al 2015, PMID: 25724305), is out of frame.; *BRCA1* A1708E (c.5123C>A (p.Ala1708Glu)) \| r.[5075_5152del] \| p.(Asp1692_Trp1718delinsGly) \| Three studies published this transcript, 2 minigene (Sanz et al 2010, PMID: 20215541; Millevoi et al 2010, PMID: 19404736) and 1 patient material (Quiles et a, 2016, PMID: 26780556). Millevoi et al report transcript at 30%. | | | | |

##### Table 4: Catalogue of BRCA2 conserved domains/motifs and currently known clinically important amino acid residues, and relevance for classification of *BRCA2* in-frame and terminal exon sequence variants

| **Domain/**  **Motif** | **AA start** | **AA end** | **AA alterations with Demonstrated Clinical Importance^a^** | **References and summary interpretation^b^** |
| --- | --- | --- | --- | --- |
| PALB2 Binding | 10 | 40 | D23_L105del (c.68_316del (p.Asp23_Leu105del)) | Deletion of the complete exon 3 (AA23-105) has been shown to be pathogenic by co-segregation analysis (Caputo et al 2018, PMID: 29707112).  Domain location description (Oliver et al 2009, PMID: 19609323; Xia et al 2006, PMID: 16793542). |
| BRC-1 | 1002 | 1036 | None reported | <http://www.ncbi.nlm.nih.gov/protein/NP_000050.2> |
| BRC-2 | 1212 | 1246 | None reported | <http://www.ncbi.nlm.nih.gov/protein/NP_000050.2> |
| BRC-3 | 1422 | 1453 | None reported | <http://www.ncbi.nlm.nih.gov/protein/NP_000050.2> |
| BRC-4 | 1518 | 1549 | None reported | <http://www.ncbi.nlm.nih.gov/protein/NP_000050.2> |
| BRC-5 | 1665 | 1696 | None reported | <http://www.ncbi.nlm.nih.gov/protein/NP_000050.2> |
| BRC-6 | 1837 | 1871 | None reported | <http://www.ncbi.nlm.nih.gov/protein/NP_000050.2> |
| BRC-7 | 1971 | 2005 | None reported | <http://www.ncbi.nlm.nih.gov/protein/NP_000050.2> |
| BRC-8 | 2051 | 2085 | None reported | <http://www.ncbi.nlm.nih.gov/protein/NP_000050.2> |
| DBD (DNA/  DSS1 binding domain - helical, OB1, OB2, OB3) | 2481 | 3186 | T2607P (c.7819A>C (p.Thr2607Pro))  W2626C (c.7878G>C (p.Trp2626Cys))  I2627F(c.7879A>T (p.Ile2627Phe))  E2663V (c.7988A>T (p.Glu2663Val))  D2723H (c.8167G>C (p.Asp2723His))  G2748D (c.8243G>A (p.Gly2748Asp))  I2778_Q2829del (c.8332_8487del (p.Ile2778_Gln2829del))  P2992_T3033del42 (c.8975_9100del (p.Pro2992_Thr3033del))  R3052W (c.9154C>T (p.Arg3052Trp))  N3124I (c.9371A>T (p.Asn3124Ile)) | <http://www.ncbi.nlm.nih.gov/protein/NP_000050.2>;  http://hci-exlovd.hci.utah.edu.  Pathogenic variant c.8486G>A (also recorded as Gln2829Arg) results in a transcript encoding an in-frame exon 19 deletion only (Houdayer et al 2012, PMID: 22505045), indicating that genetic variation encompassing loss of this entire exon (AA2778-2829) should be considered clinically important. The clinical impact of alteration/deletion of individual amino acids in exon 19 is not yet established. |
| NLS1 | 3263 | 3269 | None reported | Domain location description (Guidugli et al 2014, PMID: 24323938). |
| BRC-9 or TR2 | 3265 | 3330 | None reported | Note, although amino acids 3270-3305 within this fragment is reported to bind RAD51-DNA filaments (Davies and Pellegrini, 2007, PMID: 17515903), there is no sequence conservation with the BRC repeats located between aa1002 and aa2014. Domain boundaries are derived from x-ray chrystallography data are aa3265-3330 (Esashi et al 2005, PMID: 15800615; Esashi et al 2007, PMID: 17515904).  Case-control and frequency data indicate that BRCA2 c.9976A>T (p.Lys3326Ter) does not confer a high risk of cancer (OR 1.3-1.5, dependent on breast or ovarian cancer subtype (Meeks et al 2016, PMID: 26586665), demonstrating that residues at and downstream of 3327 are likely dispensable.  Position 3308 is implicated as clinically important by the observation that a nonsense variant c.9924C>G (p.Tyr3308Ter) is recognized as a high-risk pathogenic variant with reported functional relevance (Kuznetsov et al 2008, PMID 18607349); Bayes score 1122:1 from a single large kConFab family, Spurdle unpublished data). There is currently no publicly available clinical information to support pathogenicity of nonsense or frameshift variants located between positions 3309 and 3325. However, mammalian functional assay results indicate that variation leading to termination from p.3309 (c.9925) onwards would not be high-moderate risk, as inferred from comparison to wildtype–like functional assay results observed for the p.Lys3326Ter variant assayed in parallel (Mesman et al 2018, PMID: 29988080).  These data combined suggest that the C-terminal border of the BRC-9 relevant to the clinical interpretation of sequence variants in exon 27 of BRCA2 lies at p.3308. That is, a missense or nonsense variant predicted to modify or truncate protein sequence at residues from position p.3309 onwards is considered highly unlikely to be clinically important as a high-risk variant.  Caveat: the impact of variants that cause a frameshift before position 3309 but that may end after 3326 is not certain, and a hypomorphic effect cannot be excluded. |
| NLS2 | 3381 | 3385 | No | Domain location description (Guidugli et al 2014, PMID: 15290653).  See above. This domain is considered unlikely clinically relevant since it lies downstream (3’) of position 3326, and termination codon variants downstream of this position do not impact function, as observed for known pathogenic termination codon variants (PMID: 7894493). |

**^a^** Missense substitutions in denoted functional domains that are designated as Class 5 pathogenic based on multifactorial likelihood posterior probability of pathogenicity > 0.99, (listed in a web-based portal of published multifactorial likelihood studies - http://hci-exlovd.hci.utah.edu),and for which there is no/little effect on mRNA transcript profile - ***unless*** the variant results in an aberrant transcript that encodes a discrete in-frame deletion considered informative to definition of clinically important domains. (Splicing aberrations are reported for *BRCA2* c.7988A>T (p.Glu2663Val) and c.8168A>G (p.Asp2723Gly), but these did not lead to complete loss of function of the full length transcript), and missense alterations showed abrogation of functional activity using multiple assays (Walker et al., 2010, PMID: 20513136). An additional conserved region not commonly recognized as a BRCA2 domain/motif is located AA 1103-1183 (with the core of the motif from Leu1103 to Gln1129), but no pathogenic missense substitutions have been recorded for this region.

Note – The following pathogenic exonic variants known to alter mRNA splicing have been excluded from Table 4 above, as justified below:

| Variant | mRNA Change | Predicted Protein Change | Reason for exclusion |
| --- | --- | --- | --- |
| *BRCA2* R2336H (c.7007G>A (p.Arg2336His)) | r.[6938_7007del, 6842_7007del] | p.(Gly2313Alafs*31) – predominant transcript | Predominant alternate transcript (Houdayer et al 2012, PMID: 22505045; Machackova et al 2008, PMID: 18489799; Sanz et al 2010, PMID: 20215541; Thomassen et al 2006, PMID: 16792514), is out of frame. |
| *BRCA2* R2659G (c.7975A>G (p.Arg2659Gly)) | r.[7806_7976del] | p.(Ala2603_Arg2659del) | Alternate transcript is in-frame (Davy et al 2017, PMID: 28905878; Fraile-Bethencourt et al 2017, PMID: 28339459; Houdayer et al 2012, PMID: 22505045), and estimated at 26% (in a minigene assay). |
| *BRCA2* R2659K (c.7976G>A (p.Arg2659Lys)) | r.[7806_7976del] | p.(Ala2603_Arg2659del) | Alternate transcript is in-frame but level of full length transcript not assessed (Farrugia et al 2008, PMID: 18451181). |
| *BRCA2* R2659T (c.7976G>C (p.Arg2659Thr)) | r.[7806_7976del] | p.(Ala2603_Arg2659del) | Alternate transcript is in-frame but level of full length transcript not assessed (Farrugia et al 2008, PMID: 18451181). |
| *BRCA2* p.I2675V (c.8023A>G (p.Ile2675Val)) | r.[8023_8331del] | p.(Met2676_Ile2778del) | Predominant alternate transcript, based on minigene assay (Fraile-Bethencourt et al 2017, PMID: 28339459; Bonnet et al 2008, PMID: 18424508), is in frame. |
| *BRCA2* p.I2675V (c.8035G>T (p.Ile2675Val)) | r.[8034_8331del] | p.(Asp2679Phefs*43) | Alternate transcript is out of frame and estimated by minigene assay to be >90% of transcripts produced (Fraile-Bethencourt et al 2017, PMID: 28339459; Sanz et al 2010, PMID: 20215541). |
| *BRCA2* T2722R (c.8165C>G (p.Thr2722Arg)) | r.[7977_8331]del | p.(Tyr2660Phefs*43) | Alternate transcript level is not assessed (Fackenthal et al 2002, PMID: 12145750). |
| *BRCA2* D2723G (c.8168A>G (p.Asp2723Gly)) | r.[8168_8331]del | p.(Gly2724Phefs*3) | Six studies report alternate splicing (Houdayer et al 2012, PMID: 22505045; Fraile-Bethencourt et al 2017, PMID: 28339459; Sanz et al 2010, PMID: 20215541; Thery et al 2011, PMID: 21673748; Walker et al 2010,PMID: 20513136; Rodríguez-Balada et al 2016, PMID: 27886673), estimated at 26% in minigene by Fraile-Bethencourt et al (PMID: 28339459). |

### Appendix D: PVS1 and justification for variable weighting and considerations for code application

#### Justification for PVS1 code weights according to variant location in the gene

Loss of function of *BRCA1* or *BRCA2* is a well established mechanism of disease. Pathogenic variants in these genes are designated as predisposing to “Hereditary breast and ovarian cancer syndrome”, but they are also associated with increased risk of other cancer types (relative to the general population), as described in section on segregation (see **Appendix J**). This committee considers female breast, ovarian, male breast and pancreatic cancer as the most relevant phenotypes associated with autosomal dominant *BRCA1* and *BRCA2* disease. In addition, prostate cancer diagnosis can be used in segregation analyses that take into account age-specific penetrance.

An overview of the interpretation of predicted loss-of-function variants in *BRCA1* and *BRCA2*, adapted from the scheme of Tayoun et al 2018 (PMID: 30192042), is shown in Figures 5-8. These interpretations are based on the following considerations:

- *BRCA1* transcript NM_007294.3 and *BRCA2* transcript NM_000059.3 are denoted as the predominant and biologically relevant transcripts for variant interpretation, and build on previous research publications (Colombo et al 2014, PMID: 24569164; Fackenthal et al 2016, PMID: 27060066).
- ATG re-initiation sites that have been shown experimentally to lead to in-frame transcripts (*BRCA1* c.142, Buisson et al 2006, PMID: 16941470; *BRCA2* c.370, Parsons et al 2015, PMID: 24302565) also result in loss of important functional domains; further ATG re-initiation is not considered relevant for pathogenicity interpretation since multiple predicted termination codon variants 5' to these positions have been reported clinically as pathogenic.
- Information in Tables 3 and 4 that guides interpretation of variants predicted to lead to in-frame insertion-deletions or in-frame exon skipping predicted to lead to expression of a functional protein, and the most 3' termination position considered pathogenic considering functional and clinical data combined.
- Information in Table 5 that guides interpretation of variants predicted to impact mRNA splicing via disruption of donor and acceptor sites.
- Dosage sensitivity analysis, to justify StandAlone weight for full gene deletion

*BRCA1*: <https://dosage.clinicalgenome.org/clingen_gene.cgi?sym=brca1&subject=>

*BRCA2*: <https://dosage.clinicalgenome.org/clingen_gene.cgi?sym=brca2&subject=>

On the basis of the triage as denoted in Figures 3-6, predicted “loss of function” *BRCA1* or *BRCA2* variants may be assigned codes PVS1_StandAlone, PVS1, PVS1_Strong, PVS1_Moderate, PVS1_Supporting, or PVS1_N/A.

##### Figure 3: *BRCA1* PVS1 decision tree – initiation, nonsense/frameshift, deletion, duplication

PTC from position c.5419 onwards are not predicted to result in NMD.

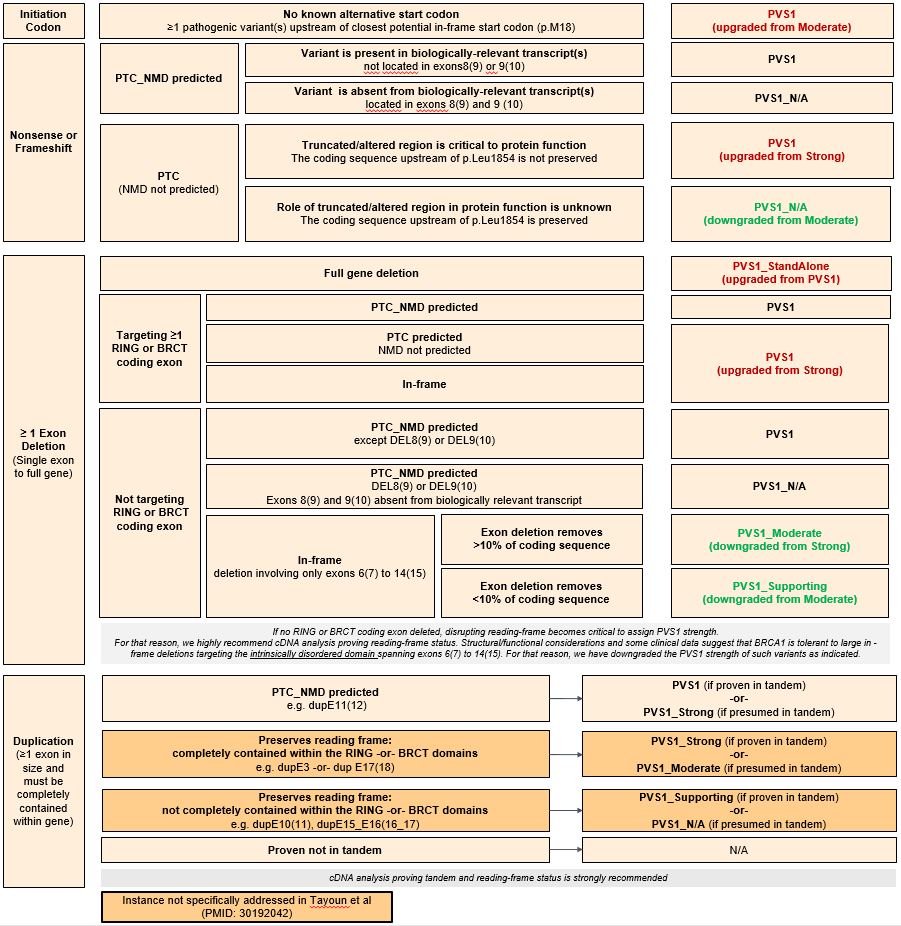

##### Figure 4: *BRCA1* PVS1 decision tree – splice sites

As detailed in Appendix E, apply as PVS1_CodeStrength for variants without in vitro splicing data, or as PVS1_CodeStrength (RNA) where variant impact is confirmed by mRNA assay results. See following page for more information. Splice sites in this context refers to the donor and acceptor ±1,2 intron positions.

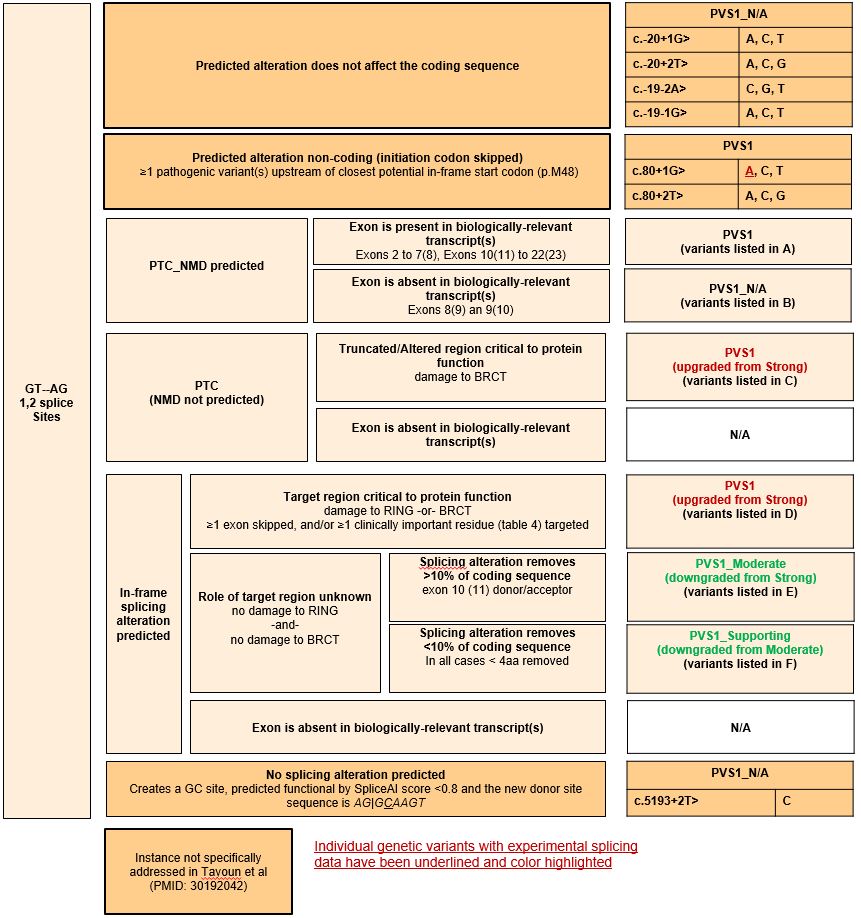

**Figure 4 continued *BRCA1* PVS1 decision tree – splice sites**

Additional information, lists of variants as noted in the decision tree above.

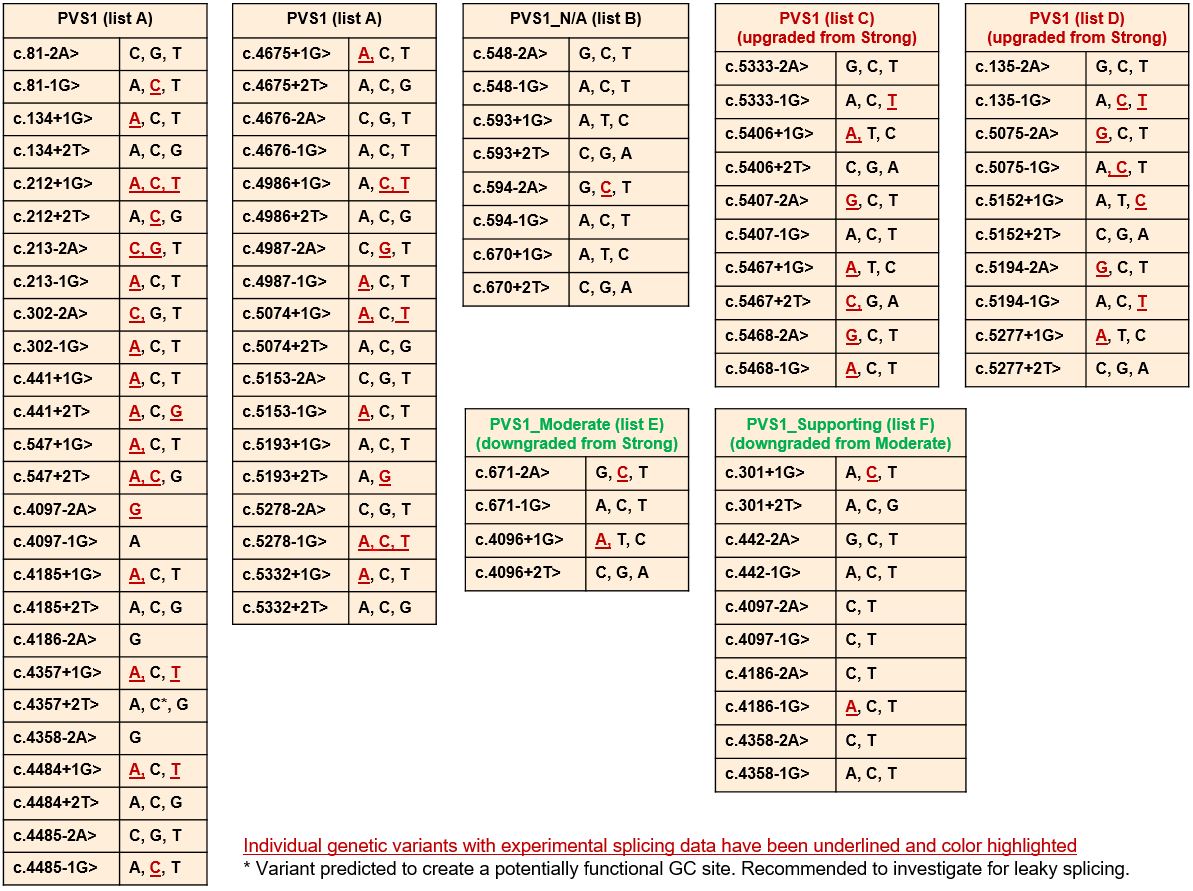

##### Figure 5: *BRCA2* PVS1 decision tree – initiation, nonsense/frameshift, deletion, duplication

PTC from position c.9600 onwards are not predicted to result in NMD.

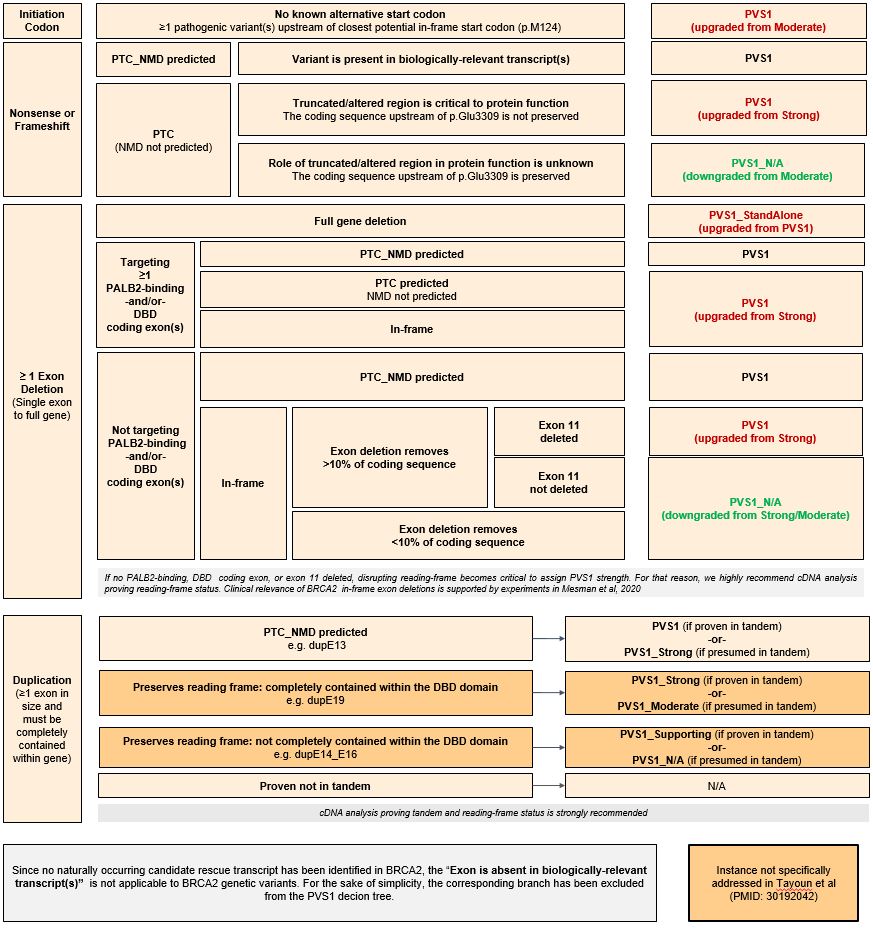

##### Figure 6: *BRCA2* PVS1 decision tree – splice sites

As detailed in Appendix E, apply as PVS1_CodeStrength for variants without in vitro splicing data, or as PVS1_CodeStrength (RNA) where variant impact is confirmed by mRNA assay results. See following page for more information. Splice sites in this context refers to the donor and acceptor ±1,2 intron positions.

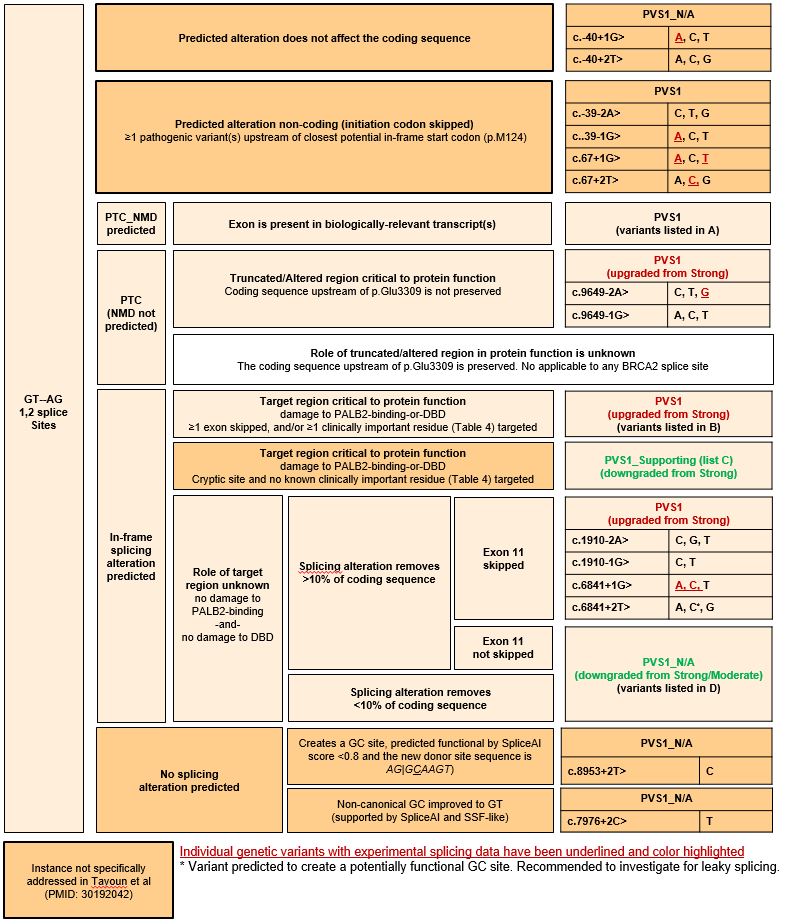

**Figure 6 continued *BRCA2* PVS1 decision tree – splice sites**

Additional information, lists of variants as noted in the decision tree above.

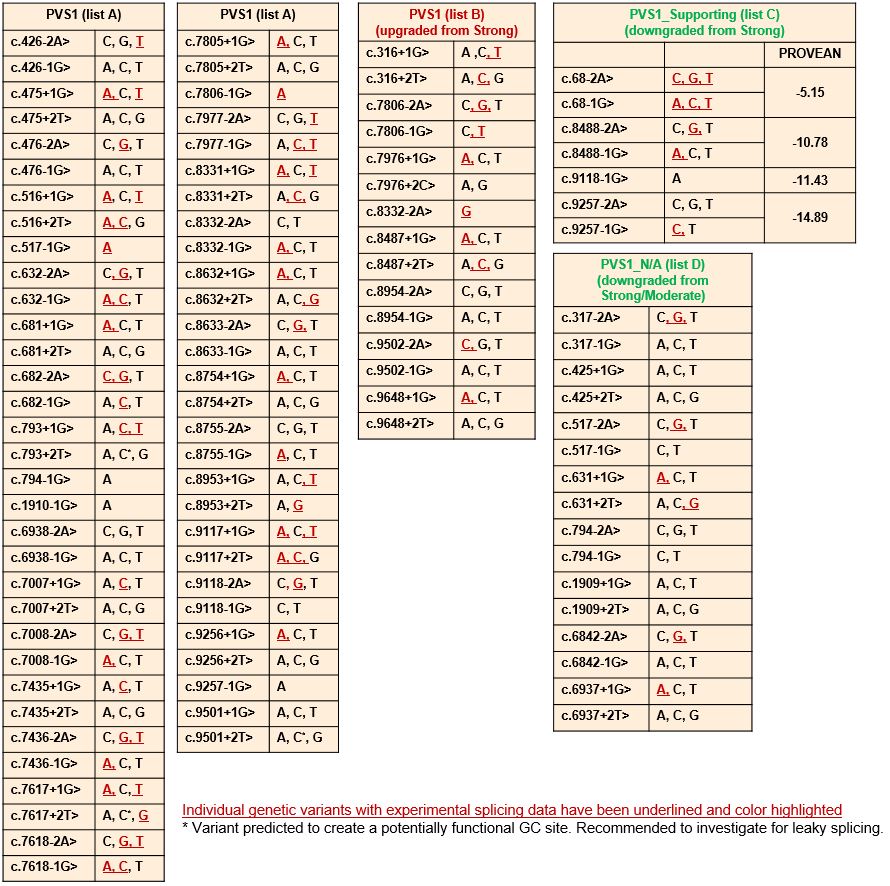

##### **Table 5: *BRCA1* and *BRCA2* exon boundary donor/acceptor ±1,2 variants with altered PVS1 evidence strength due to (predicted) functional in-frame transcripts that (may) rescue gene functionality.**

| **Donor/Acceptor ±1,2 positions** | **Predicted rescue mechanism** | | | **Rationale** |
| --- | --- | --- | --- | --- |
|  | **Normal levels of**  **naturally occurring**  **rescue**  **transcript^*^** | **Upregulated**  **levels of**  **naturally occurring**  **rescue**  **transcript^*^** | **Variant**  **induced rescue transcript** |  |
| ***BRCA1***** | | | | |
| c.301+1  c.301+2 | - | - | **Δ5(6)q9** | **Exon 5(6) donor site.** In silico, all six variants score very similar, with SpliceAI predicting PTC-NMD transcripts Δ5(6) and MES identifying an in-frame cryptic donor site. Experimental data for c.301+1G>C (Paxgene RNA) do not detect Δ5(6) transcripts, but confirms use of the cryptic site to produce in-frame Δ5(6)q9 (p.Gly98_Tyr101delinsAsp) transcripts (Leman et al 2018, PMID: 29750258). Based on that, we have assigned a pathogenic supporting evidence (**PVS1_Supporting**). Of note, all six donor site variants score damaging in a functional assay (Findlay et al 2018, PMID:30209399), but c.301+1G>A, also producing Δ5(6)q9, has been detected in trans with a stop-gain variant in one healthy individual without Fanconi Anemia features (Ambry Genetics internal data, ClinVar VCV000142004.13).  *These apparently conflicting evidence types do not prove but might be compatible with an intermediate risk variant.* |
| c.442-1  c.442-2 | - | **Δ7(8)p3** | - | **Exon 7(8) donor site.** Experimentally validated tandem acceptor site (NAGNAG). Variant alleles are predicted to inactivate the 5’ but not the 3’ tandem site, resulting in upregulation of Δ7(8)p3, a short in-frame alteration not targeting clinically relevant domains. Upregulation has been observed for the nearby c.442-3T>G variant (Houdayer et al 2012, PMID: 22505045). Based on Δ7(8)p3 **(**p.Gln148del), we have assigned a pathogenic supporting evidence (**PVS1_Supporting**). |

| c.548-1  c.548-2  c.593+1  c.593+2  c.594-1  c.594-2  c.670+1  c.670+2 | **Δ8,9 (Δ9,10)** | - | - | **Exon 7(8) and exon 9(10) acceptor and donor sites.** Combined clinical and splicing data on c.591C>T and c.594-2A>C (shown from ENIGMA research to co-occur in cis with c.641A>G), suggests that alleles expressing normal levels of naturally occurring in-frame Δ8,9(9,10) transcripts do not increase risk, even if splicing alterations and/or stop-gain variants are observed in exons 8(9) or 9(10) (Dosil et al 2010, PMID:19892845; Rosenthal et al 2015, PMID:25639900; de la Hoya et al 2016, PMID:27008870).  Alleles carrying donor or acceptor splice site variants targeting exons 8(9) or 9(10) are predicted to produce exon 8(9) or exon 9(10) skipping (both PTC-NMD), but also normal (or upregulated) levels of Δ8,9(9,10) transcripts. Based on that, we have not assigned pathogenic evidence at any strength (**PVS1_N/A**). |
| --- | --- | --- | --- | --- |
| c.671-2  c.671-1 | - | **Δ10(11)**  **Δ8-10(9-11)** | - | **Exon 10(11) acceptor site.** All six variants score similar with SpliceAI predictions. Experimental splicing data shows that c.671-2A>C produces PTC-NMD transcripts Δ9-10(10-11) and in-frame transcripts Δ8-10(9-11) and particularly Δ10(11) (Keaton et al 2003, PMID:14513821; Lattimore et al 2018, PMID:29774201). Based on similarities with exon 10(11) donor site variants (see below), we have assigned pathogenic with moderate evidence strength (**PVS1_Moderate**). |
| c.4096+1  c.4096+2 | - | **Δ10(11)q3309**  **Δ10(11)** | - | **Exon 10(11) donor site.** Data collected by the ENIGMA consortium demonstrates that the *BRCA1* c.4096+1G>A variant, proven to result in upregulation of naturally occurring in-frame transcripts Δ10q3309 (Bonatti et al 2006, PMID:17011978) and also Δ10 (Radice, unpublished data), may not exhibit the clinical characteristics of a standard high-risk pathogenic *BRCA1* variant (Spurdle, unpublished data). Based on that, we have assigned pathogenic with moderate evidence strength (**PVS1_Moderate**).  *Existing data on exon 11 donor site variants do not prove but might be compatible with an intermediate risk variant* |
| c.4097-2  c.4097-1 | - | **-** | **Δ11(12)p6** | **Exon 11(12) acceptor site.** In silico, four of the six variants at this acceptor site (c.4097-2A>C/T, c.4097-1G>C/T) are predicted to use an alternate acceptor site 6nt into exon 11(12). This creates an in-frame transcript and is assigned supporting evidence for pathogenicity (**PVS1_Supporting**). The other two variants, c.4097-2A>G and c.4097-1G>A, are predicted to create 1nt insertion and deletion, respectively. We have assigned **PVS1** to these variants. |
| c.4186-1  c.4186-2 | - | **Δ12(13)p3** | - | **Exon 12(13) acceptor site.** Experimentally validated tandem acceptor site (NAGNAG). Variant alleles are predicted to inactivate the 5’ but not the 3’ tandem site, producing Δ12(13)p3 transcripts only (i.e. upregulating Δ12(13)p3), a short in-frame alteration not targeting clinically relevant domains. Based on Δ12(13)p3 p.(Gln1396del), we have assigned a pathogenic supporting evidence (**PVS1_Supporting**) to exon 12(13) acceptor site variants. The only exception being c.4186-2A>G, predicted to damage both tandem acceptor sites, causing exon 12(13) skipping (a PTC_NMD alteration). Based on that, we have assigned a pathogenic very strong evidence (**PVS1)** to this specific variant. |

| c.4358-1  c.4358-2 | - | **Δ13(14)p3** | - | **Exon 13(14) acceptor site.** Experimentally validated tandem acceptor site (NAGNAG). Variant alleles are predicted to inactivate the 5’ but not the 3’ tandem site, producing Δ13(14)p3 transcripts only (i.e. upregulating Δ13(14)p3), a short in-frame alteration not targeting clinically relevant domains. Based on Δ13(14)p3 p.(Ala1453del), we have assigned a pathogenic supporting evidence to exon 13(14) acceptor site variants (**PVS1_Supporting**).The only exception being c.4358-2A>G, predicted to damage both tandem acceptor sites, causing exon 13(14) skipping (a PTC_NMD alteration). Based on that, we have assigned a pathogenic very strong evidence (**PVS1**) to this specific variant. |
| --- | --- | --- | --- | --- |
| ***BRCA2*** | | | | |
| c.68-2  c.68-1 | - | - | **∆E3p6** | **Exon 3 acceptor site.** In silico, all six variants score very similar, with SpliceAI predicting acceptor loss and acceptor gain 6nt into exon 3, resulting in an in-frame deletion ∆(E3p6). No known pathogenic missense variants are within this 6nt region, however the BayesDel prediction of this in-frame deletion is above the threshold for PP3. We have assigned pathogenic evidence at the supporting strength (**PVS1_Supporting**) to exon 3 acceptor site variants. |
| c.317-2  c.317-1 | - | - | **Δ4,5** | **Exon 4 acceptor site.** In silico, all six variants score very similar, with SpliceAI predicting exon 4 skipping (PTC-NMD). Experimental splicing data indicates that c.317-2A>G produces Δ4, but also a significant proportion of Δ4,5 (Fraile-Bethencourt et al 2019, PMID: 30883759). Based on similarity with exon 4 donor site variants, we have not assigned pathogenic evidence at any strength (**PVS1_N/A**) to exon 4 acceptor site variants. |
| c.425+1  c.425+2 | - | **Δ4,7** | **Δ4,5** | **Exon 4 donor site.** In silico, all six variants score very similar, with SpliceAI predicting exon 4 skipping (PTC-NMD). Clinical, functional, and splicing data indicates that c.425G>T (with very similar SpliceAI predictions) is (partially) functional and benign, presumable due to production (in addition to Δ4) of in-frame Δ4,5 and/or Δ4,7 transcripts (Nix et al, 2021, PMID: 33469799) (Mesman et al 2020, PMID: 32398771). Based on that, we have not assigned pathogenic evidence at any strength (**PVS1_N/A**) to exon 4 donor site variants. |
| c.517-1  c.517-2 | - | **Δ4,7** | - | **Exon 7 acceptor site.** With the exception of c.517-1G>A, all variants score very similar, with SpliceAI predicting Δ7 (PTC_NMD). Functional, and splicing data (Mesman et al 2020, PMID: 32398771) indicates that c.517-2A>G is (partially) functional, presumable due to production of in-frame Δ4,7 transcripts. Based on that (and similarity with exon 4 donor sites), we have not assigned pathogenic evidence at any strength (**PVS1_N/A**) to exon 7 acceptor site variants. Variant c.517-1G>A is predicted (and observed) to produce Δ7 and Δ7p1 (both PTC_NMD) (Menéndez et al 2016, PMID: 21735045; Fraile-Bethencourt et al 2019, PMID: 30883759). Based on that, we have assigned **PVS1 (RNA)** to this specific variant. |
| c.631+1  c.631+2 | - | **Δ4,7** | - | **Exon 7 donor site.** In silico, all six variants score very similarly, with SpliceAI predicting PTC-NMD Δ7 transcripts. Functional, and splicing data (Mesman et al 2020, PMID: 32398771) indicates that c.631+2T>G is (partially) functional, presumably due to production of in-frame Δ4,7 transcripts. Based on that (and similarity with exon 4 donor sites), we have not assigned pathogenic evidence at any strength (**PVS1_N/A**) to exon 7 donor site variants, except for c.631+1G>A. For c.631+1G>A, experimental data has detected Δ7 but not the candidate rescue Δ4,7 (Fraile-Bethencourt et al 2019, PMID: 30883759). Based on that, we have assigned **PVS1 (RNA)** to this specific variant. |
| c.794-2  c.794-1  c.1909+1  c.1909+2 |  |  | **Δ10** | **Exon 10 acceptor and donor sites.** No good SpliceAI predictions. Lacking predictions or experimental data, Δ10 assumed. Functional data indicates that Δ10 codes for a (partially) functional protein (Mesman et al 2020, PMID: 32398771). Based on that, we have not assigned pathogenic evidence at ant strength (**PVS1_N/A**). The only exception is c.794-1G>A, which is predicted by SpliceAI to create a new acceptor site 1nt into the exon, expected to result in a frame-shifting transcript. We have assigned **PVS1** to this specific variant. |
| c.6842-1  c.6842-2  c.6937+1  c.6937+2 |  | **Δ12** |  | **Exon 12 acceptor and donor sites**. Variant alleles are predicted to produce exon12 skipping only (i.e. upregulating naturally occurring Δ12). BRCA2 exon12 is functionally redundant (Li et al 2009, PMID: 19795481) (Meulemans et al 2020, PMID: 32046981)(Mesman et al 2020, PMID: 32398771). Based on that, we have not assigned pathogenic evidence at any strength (**PVS1_N/A**) to exon 12 donor and acceptor site variants. |
| c.8488-2  c.8488-1 |  |  | **ΔE20p12** | **Exon 20 acceptor site.** Variants at this site are reported to produced partial intron retention (12nt) as well as exon skipping and intron retention (Acedo et al 2015, PMID: 25382762; Howlett et al 2002, PMID: 12065746; Santos et al 2014, PMID: 24607278; Acedo et al 2012, PMID: 22632462). This deletion removes part of the DNA-binding domain but no known pathogenic missense variants (Table 4). Based on this information we have assigned supporting strength pathogenic evidence (**PVS1_Supporting**) to exon 20 acceptor site variants. |
| c.9118-1G>A |  |  | **ΔE24p24** | **Exon 24 acceptor site variant.** In silico, the c.9118-1G>A variant is predicted by SpliceAI to create an acceptor site 24nt into exon 24, resulting in an in-frame transcript. This deletion removes part of the DNA-binding domain but no known pathogenic missense variants (Table 4). Based on this information we have assigned supporting strength pathogenic evidence (**PVS1_Supporting**) to this specific variant. |
| c.9257-2  c.9257-1 |  |  | **ΔE25p27** | **Exon 25 acceptor site.** In silico, all variants (except for -1G>A) have similar predictions to create an acceptor site 27nt into exon 25, resulting in an in-frame transcript. This deletion removes part of the DNA-binding domain but no known pathogenic missense variants (Table 4). Based on this information we have assigned supporting strength pathogenic evidence (**PVS1_Supporting**). The only exception is c.9257-1G>A, which is predicted by SpliceAI to create a new acceptor site 1nt into the exon, expected to result in a frame-shifting transcript. We have assigned **PVS1** to this specific variant. |
| ^*^ Colombo et al 2014 (PMID: 24569164); Fackenthal et al 2016 (PMID: 30890586),  ** For *BRCA1*, legacy exon numbering is in parenthesis. | | | | |

#### Considerations for PVS1 code application for predicted termination codon variants

*BRCA1* and *BRCA2* pathogenic variants are considered clinically actionable. Dependent on sex, age and cancer status of carriers, risk management options include: high risk breast surveillance; eligibility for risk reducing surgery, including bilateral risk-reducing management and bilateral salpingo-oophorectomy; precision management of cancer-affected individuals with PARP inhibitors and platinum based chemotherapy; cascade genetic testing for relatives. The penetrance estimates and age-specific cancer incidence tables used to define these clinical management strategies have largely been derived from a combined analysis of 22 population-based studies (Antoniou et al 2003, PMID: 12677558), and more recently from an analysis of prospective data drawn from 3 large cohort studies capturing data from the USA, Canada, UK, Europe and Australia (Kuchenbaecker et al 2017, PMID: 28632866).

The population-based analysis (PMID: 12677558) described “mutations” included as nonsense, frame-shift, deletion, consensus splice site, large deletion and large duplication variants i.e. variants considered generically as (likely) loss of function. The cohort study (PMID: 28632866) provided no details regarding predicted mechanism for pathogenicity of “mutations” included in the analysis, but it is presumed that these were predominantly (likely) loss of function variants. These studies provided convincing evidence that carriers of a predicted loss of function *BRCA1* or *BRCA2* variant have a high absolute risk of breast and ovarian cancer, and that risk is considerably elevated relative to population-specific incidence. See Table 6 below for details. In addition, recent population-based case-control analysis (Dorling et al 2021, PMID: 33471991) showed that risk of breast cancer is >4-fold for predicted termination codon variants specifically (excluding variants in the last exon) in *BRCA1* and in *BRCA2*.

##### Table 6: Breast and ovarian cancer risk estimated from pooled analysis of *BRCA1* and *BRCA2* pathogenic variant carriers

| **Gene** | **Cancer Type** | **Antoniou et al 2003**  **Population-based analysis**  **Cumulative risk to**  **age 70y% (95% CI)** | **Kuchenbaecker et al 2017**  **Prospective analysis**  **Cumulative risk to**  **age 80y% (95% CI)** | **Kuchenbaecker et al 2017**  **Prospective analysis**  **Standardized Incidence Rate (95%CI)** |
| --- | --- | --- | --- | --- |
| *BRCA1* | Breast | 65% (51%-75%) | 72% (65%-79%) | 16.6 (14.7-18.7) |
|  | Ovarian | 39% (22%-51%) | 44% (36%-53%) | 49.6 (40.0-61.5) |
| *BRCA2* | Breast | 45% (33%-54%) | 69% (61%-77%) | 12.9 (11.1-15.1) |
|  | Ovarian | 11% (4.1%-18%) | 17% (11%-25%) | 13.7 (9.1-20.7) |

While these data combined justify assignment of a Likely Pathogenic assertion as a group for variants that meet ACMG/AMP PVS1 following the conservative scheme outlined in Figure 3-6, we propose to apply exon-specific weights for protein termination codon (PTC) variants based on prior evidence for other protein termination codons in that exon. Following recommendations from the ClinGen SVI, the code applied will be a modification of the PM5 code, which we term PM5_PTC. The data to justify exon-specific weights is provided in Supplementary Table 1, and assumes that the molecular effect of (and risk associated with) PTC variants in a given exon will be the same for variants predicted to lead to NMD.

#### Justification for PM5_PTC exon-specific weights

Exon-specific weights are based on:

(i) The observation of impact on function for at least one PTC variant in an exon, where this variant is not predicted (or shown) to alter splicing with potential to rescue function, can be used to infer code PS3 for another PTC variant in the same exon.
(ii) The observation of impact on function for at least one missense substitution variant in the exon, indicating that there is no rescue of pathogenic variants due to alternative splicing, and that a PTC variant leading to NMD, would have at minimum the same impact. Can be used to infer code PS3_moderate for another PTC variant in the same exon.
(iii) Case-control OR ≥4.0 estimated for PTC variants observed in a given exon, from well-designed case-control studies, can be used to infer code PS4 for any PTC variant in that exon. OR ≥4 non-significant considered sufficient to apply as PS4_supporting.
(iv) Heterogeneity analysis using personal and family history profile as a predictor of pathogenicity, stratifying PTC variants by exon, using dataset and methods described in Li et al 2020 (PMID: 31853058), with LRs converted to appropriate weight.
(v) Standardized incidence ratios (SIR) for PTC variants in a given exon, comparing frequency of PTCs in non-Finnish European (NFE) probands with breast, ovarian, pancreatic cancer versus the summed frequency of PTC in gnomAD NFE, using the same dataset as in Li et al 2020 (PMID: 31853058). SIR ≥ 4, P<0.05 was used to infer code PS4, and SIR ≥4 non-significant as PS4_supporting.
(vi) Identification of PTC variants in a given exon in multiple independent probands in the CIMBA *BRCA1/2* cohort, based on the knowledge that individuals in this highly ascertained cohort had met strict clinical criteria to undergo diagnostic testing. Conservatively, observation of ≥5 unique PTCs variant in ≥ 5 families was used to infer PP4, supporting evidence in favour of pathogenicity.

The per-exon evidence was then summed using a points-based approach (Supporting = 1, Moderate = 2, Strong = 4). Based on the combined evidence, the PM5_PTC code can be applied as Strong evidence in favour of pathogenicity for most exons. See Figure 7 and Figure 8 below, and Supplementary Table 1 for further details.

PM5_PTC code can only be applied to germline variants that meet PVS1 codes, namely nonsense and frame-shift changes, including large deletions and tandem duplications. Code weight is determined by the exon in which the predicted termination codon occurs. For example, a frame-shift variant in BRCA2 exon 15 that is predicted to result in a termination codon within exon 16 would use the code strength of BRCA2 exon 16 (PM5_PTC_Strong).

Variants at the donor/acceptor ±1,2 positions do not qualify for PM5_PTC code.

##### Figure 7: Overview of evidence supporting PM5_PTC weights for *BRCA1*

BRCA1

PM5_PTC Points Assigned

| Exon (Legacy Exon) | Fxn PTC | Fxn missense | CaCo OR ≥4 | Fam History LR | SIR ≥4 | CIMBA counts |
| --- | --- | --- | --- | --- | --- | --- |
| 2 (2) | 4 | 2 |  | 1 |  | 1 |
| 3 (3) | 4 | 2 |  | 4 | 4 | 1 |
| 4 (5) | 4 | 2 | 4 | 4 |  | 1 |
| 5 (6) | 4 | 2 |  |  |  | 1 |
| 6 (7) |  |  | 1 | 4 | 4 | 1 |
| 7 (8) |  |  | 1 | -1 |  | 1 |
| 10 (11) |  |  | 4 | 4 | 4 | 1 |
| 11 (12) |  |  | 1 | 4 |  | 1 |
| 12 (13) |  |  | 1 | 4 | 4 | 1 |
| 13 (14) |  |  | 1 | 2 |  | 1 |
| 14 (15) |  |  | 1 | 4 | 4 | 1 |
| 15 (16) | 4 | 2 |  | 4 | 4 | 1 |
| 16 (17) | 4 | 2 |  | 4 | 4 | 1 |
| 17 (18) | 4 | 2 |  | 4 | 4 | 1 |
| 18 (19) | 4 | 2 | 1 | 4 | 4 | 1 |
| 19 (20) | 4 | 2 | 1 | 2 |  | 1 |
| 20 (21) | 4 | 2 |  | 2 |  | 1 |
| 21 (22) | 4 | 2 |  | 4 | 4 | 1 |
| 22 (23) | 4 | 2 |  |  |  | 1 |
| 23 (24) | 4 | 2 |  | 4 | 4 | 1 |

##### Figure 8: Overview of evidence supporting PM5_PTC weights for *BRCA2*

BRCA2

PM5_PTC Points Assigned

| Exon | Fxn missense | CaCo OR ≥4 | Fam History LR | SIR ≥4 | CIMBA counts |
| --- | --- | --- | --- | --- | --- |
| 2 | 2 |  | 1 |  | 1 |
| 3 | 2 |  | 4 | 4 | 1 |
| 4 |  | 1 |  |  | 1 |
| 5 |  | 1 | 4 |  | 1 |
| 7 |  |  | 4 | 4 | 1 |
| 8 |  | 1 | 4 | 4 | 1 |
| 9 |  | 1 | 2 |  | 1 |
| 10 |  | 4 | 4 | 4 | 1 |
| 11 |  | 4 | 4 | 4 | 1 |
| 13 |  | 4 | 4 | 4 | 1 |
| 14 |  | 1 | 4 |  | 1 |
| 15 | 2 | 1 | 4 | 4 | 1 |
| 16 | 2 | 1 | 4 |  | 1 |
| 17 | 2 | 1 | 4 | 4 | 1 |
| 18 | 2 |  | 4 | 4 | 1 |
| 19 | 2 |  | 2 | 4 | 1 |
| 20 | 2 | 4 | 4 | 4 | 1 |
| 21 |  |  |  |  | 1 |
| 22 | 2 | 1 | 4 | 4 | 1 |
| 23 | 2 | 4 | 4 | 4 | 1 |
| 24 | 2 | 1 | -1 | 4 | 1 |
| 25 | 2 | 1 | 4 | 4 | 1 |
| 26 |  |  |  |  | 1 |
| 27 |  |  | 4 |  | 1 |

### Appendix E: PS3 and BS3 code use

In consultation with members of the ClinGen SVI Group, and the ClinGen SVI Splicing Subgroup, evidence from splicing assays will be not be assigned PS3 and BS3 codes, but rather noted as additional annotation under the PVS1 or BP7 code. PS3 and BS3 codes are reserved for assigning variant impact on protein function, but are required to take into consideration functional assay design, variant location, and predicted/observed impact on splicing as well as predicted impact on the encoded protein sequence. Figure 9 provides an overview of the sequential approach to assign codes based bioinformatic predictions and results from assays measuring variant effect on mRNA transcripts and/or protein function.

##### Figure 9: Decision tree for assessing predicted and experimentally investigated impact of a variant on mRNA and protein function

See Appendix J and K for details on application of bioinformatic codes. BP7 is applied for intronic variants at or beyond +7 or -21 positions, where BP4 is met, based on low prior probability for alternative mechanism. See Bioinformatic section (Appendix J) for additional justification. Tables 7-9 together summarise findings that inform the suggested application of weights for splicing data. Current data (Table 7) suggests that mRNA results for donor/acceptor ±1,2 positions that are consistent with the expected PVS1 code (Figure 4 and Figure 6) may be applied irrespective of formal allele-specific quantitation. In contrast, quantitation is more important to assess pathogenicity associated with spliceogenicity for variants outside of these positions (Table 8). Table 9 summarises suggesting weighting of mRNA data for variants outside of the donor/acceptor ±1,2 positions, according to assay methodology and proportion of functional transcript retained. Based on current findings, near-complete or substantial expression of aberrant transcripts is defined as ≥ 90% of per-allele expression profile from quantitative assays. See Table 3 and Table 4 for rationale to define (potentially) clinically important functional domains, currently denoted as: BRCA1 RING aa 2-101; BRCA1 coiled-coil aa 1391-1424; BRCA1 BRCT repeats aa 1650-1857; BRCA2 PALB2 binding domain aa 10-40; BRCA2 DNA binding aa 2481-3186.

.

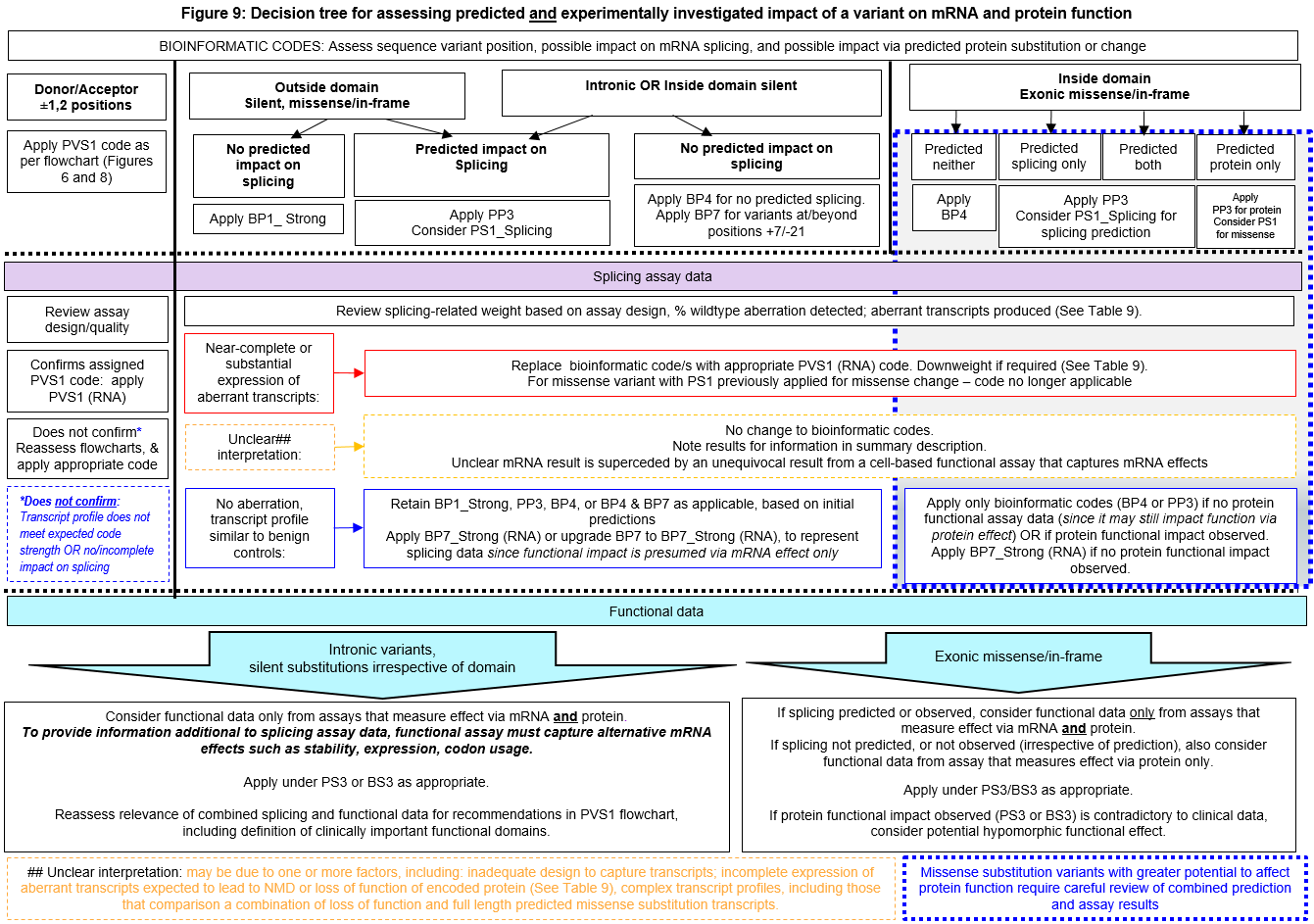

#### Splicing assay data interpretation and evidence weights

Recommendations for the conduct and interpretation of mRNA assay data for variant classification are drawn in part from (Walker et al., 2013, PMID:23893897). Assessment assumes assays on mRNA from patient germline tissue samples (fresh blood, cultured lymphocytes, lymphoblastoid cell lines etc), compared with assays performed in tandem on mRNA from the same tissue type for reference controls, with preference for 5-10 controls. Transcripts identified at similar levels in controls are considered to be naturally occurring isoforms and not mRNA aberrations. A variant is considered spliceogenic predicted loss of function (Spurdle et al., 2019, PMID:30962250) if it results in an altered mRNA transcript profile that is predicted to cause gene loss-of-function i.e. any combination of mRNA transcripts predicted non-coding, predicted protein truncating-NMD, and/or predicted to encode proteins lacking critical structural/functional motifs. Sequencing of the full length transcript for the variant allele (if exonic), or a common polymorphism in *cis* (if variant is intronic), is currently considered adequate to assess if variant allele contributes to production of wild-type transcript. A variant may be reported as not associated with an mRNA aberration (splicing or expression) if the variant allele produces transcript patterns comparable to that of controls.

Experimental design should be considered when assessing the quality of a published mRNA splicing assay. Particularly, primer/location design should be reviewed for ability to detect predicted potential aberrant transcripts, guided by knowledge of naturally occurring isoforms (eg multi-exon skipping previously identified in controls). Use (or not) of NMD inhibitors should be noted, and considered for the interpretation of relative levels of transcripts that are expected to be subject to NMD. Results from specific construct-based mRNA assays may be considered to aid variant interpretation if the transcript profile observed for wildtype sequence from the construct-based assay adequately represents the transcript profile observed for wildtype sequence from blood-based assays AND The construct-based mRNA assay provides estimates of allele-specific extent of aberration e.g. extent of intron exclusion or inclusion, of particular relevance for intronic variants AND/OR Results from a construct-based mRNA assay are not materially different to those observed for construct-based assays for another spliceogenic variant that has been classified using clinical information. Studies with inadequate experimental design and conditions to detect predicted potential aberrations should be noted as excluded (with appropriate explanation) from a finalized summary of variant impact on splicing.

Tables 7-9 below provide data that informs the suggested application of weights for splicing data, based on assay methods commonly used in the literature to date. Weights are based on consideration of splicing assay result category (patient and/or construct data, allele-specific quantitation) for variants previously classified as pathogenic based on multifactorial likelihood analysis – an analysis which did not consider splicing assay data. The coding for variants, and source of data used for analysis is shown in Supplementary Tables 2 and 3. Variants with high potential to impact function via missense change (A-GVGD category C65), were exclude to limit confounding by alternative mechanism of pathogenicity. As shown in Table 7, all variants at the donor and acceptor ±1,2 positions that met PVS1 “bioinformatic” code as per Figures 4 or 6, were shown to lead to loss of function transcripts. Further, all these variants were classified as pathogenic, irrespective of the splicing assay result category. This finding indicated that splicing assay result category alone should not be considered to downweight PVS1 codes applied for variants at the highly conserved donor and acceptor ±1,2 positions.

##### **Table 7: Splicing assay results for variants** at the donor or acceptor ±1,2 positions **classified as (Likely) Benign or (Likely) Pathogenic using multifactorial likelihood** analysis

| **Splicing Assay Result Category*** | **(Likely) Benign (n)** | **(Likely) Pathogenic (n)** |
| --- | --- | --- |
| Patient plus allele-specific; no aberration seen | 0 | 0 |
| Patient not allele-specific OR minigene only; no aberration seen | 0 | 0 |
| Patient not allele-specific; aberrant transcripts consistent with loss of function | 0 | 17 |
| Minigene only; aberrant transcripts consistent with loss of function | 0 | 4 |
| Patient plus allele-specific to confirm not leaky; aberrant transcripts consistent with loss of function | 0 | 22 |
| **All splicing assays (irrespective of assay result category)** | **0** | **43** |

* See above for description of patient material sources. Patient plus allele-specific includes allele-specific patient assay OR non-allele-specific patient assay supplemented with minigene results to assess assay leakiness.

Table 8 shows similar categorization for variants outside the donor and acceptor ±1,2 dinucleotides. All but 1 variant with no impact on splicing was classified as benign; the single exception was BRCA2 c.5144G>A p.S1715N with pathogenicity due to missense alteration (a C45 A-GVGD missense substitution prediction). Classification was (likely) pathogenic for 33/36 variants shown to lead to aberrant transcripts coded as loss of function. The three exceptions were: *BRCA2* c.520C>T p.R174C impacting splicing via ESE impact (3 different minigene assays demonstrating exon 7 deletion, level 60-66%); *BRCA2* c.9501+3A>T (multiple assays showing impact, reported levels ranging from 13% exon skipping to severe/complete impact); *BRCA1* c.5453A>G p.D1818G impacting splicing via ESE impact (two patient assays, one reported as complete). These observations highlight several issues – exclusion of ESE predictions in classification algorithms (due to their poor predictive value); incomplete impact on splicing. This review of obvious discrepancies with existing classifications suggested the following: variants with no impact on splicing can be assigned a strong benign code weight irrespective of assay result category; but for variants that result in aberrant transcripts consistent with loss of function, maximum weight should be only assigned for the aberration type if variant impact is quantified as complete or near complete, with downweighting if minigene only or patient not quantitated.

##### **Table 8: Splicing assay results for variants classified as (Likely) Benign or (Likely) Pathogenic using multifactorial likelihood** analysis - variants excluding those at donor or acceptor ±1,2 positions

| **Splicing Assay Result Category*** | **(Likely) Benign (n)** | **(Likely) Pathogenic (n)** |
| --- | --- | --- |
| Patient plus allele-specific; no aberration seen | 38 | 0 |
| Patient not allele-specific OR minigene only; no aberration seen | 102 | 1 |
| Patient not allele-specific; aberrant transcripts consistent with loss of function | 0 | 9 |
| Minigene only; aberrant transcripts consistent with loss of function | 1 | 3 |
| Patient plus allele-specific; aberrant transcripts consistent with loss of function | 2 | 21 |
| **All splicing assays (irrespective of assay result category)** | ***143*** | ***34*** |
| * See above for description of patient material sources. Patient plus allele-specific includes allele-specific patient assay OR non-allele-specific patient assay supplemented with minigene results to assess assay leakiness. | | |

For variants outside the donor and acceptor ±1,2 positions, further consideration of weights according to % splicing aberration were derived using a Delphi approach, and have been proposed in Table 9. These are based on published findings correlating variant pathogenicity with experimentally derived relative proportion of transcripts (inferred to be) produced by the variant allele, including predicted loss of function, rescue and wildtype transcripts. Specifically:

- Observation that different *BRCA2* spliceogenic alleles that lead to near-complete (94%, 95% or 97%) or complete (100%) expression of ∆E3 in construct based assays have clinical features of pathogenic variants, indicating that some level of leakiness (at least ~5%) may be expected for construct-based assays (PMID:29707112).
- Observation that a common synonymous variant *BRCA2* c.231T>G (maximum minor allele frequency >1%) demonstrated 30% ∆E3 in a minigene assay, and 40% ∆E3 and complementation of the null phenotype in a mESC assay (PMID: 32641407). That is, 60-70% per-allele expression of a functional transcript is sufficient for cell survival.
- Classification as benign (based on clinical evidence) for a *BRCA1* allele producing as much as 70–80% of transcript encoding tumor suppressor deficient protein (and 20-30% of in-frame rescue transcripts), as measured in blood-related samples (PMID: 27008870).

We anticipate that these recommendations will be updated to consider additional methodology and further data correlating % per allele expression of aberrant transcripts with clinical phenotype.

##### Table 9: Rubric for weighting of mRNA data for variants outside of the donor/acceptor ±1,2 positions, according to assay methodology and proportion of functional transcript retained.

| **PVS1 (RNA) or BP7 (RNA) code weight to apply, according to mRNA source, assay design/quantitation**  **AND relative level/s of functional and non-functional transcript/s^$^** | | | | | | |
| --- | --- | --- | --- | --- | --- | --- |
| **Assay results with allele-specific quantitation #** | | | | | | |
| **PVS1 Code Weight as per PVS1 flowcharts*** | **Patient with quantification#** | **Minigene only** | **Patient with quantification#** | **Minigene only** | **Patient with quantification# OR minigene only** | |
| Proportion WT or (assumed) functional transcript^ | ≤10% | ≤10% | >10 & <20% | >10 & <20% | 20-30% | >30% |
| Proportion non-functional transcript | ≥ 90% | ≥ 90% | < 90% & > 80% | < 90% & > 80% | 70-80% | <70% |
| PVS1 | PVS1 (RNA) | PVS1_Strong (RNA) | PVS1_Strong (RNA) | PVS1_Moderate (RNA) | PVS1_N/A (RNA) | BP7_Strong (RNA) |
| PVS1_Strong | PVS1_Strong (RNA) | PVS1_Moderate (RNA) | PVS1_Moderate (RNA) | PVS1_Supporting (RNA) |  |  |
| PVS1_Moderate | PVS1_Moderate (RNA) | PVS1_Supporting (RNA) | PVS1_Supporting (RNA) | PVS1_Supporting (RNA) |  |  |
| PVS1_Supporting | PVS1_Supporting (RNA) | PVS1_Supporting (RNA) | PVS1_Supporting (RNA) | PVS1_Supporting (RNA) |  |  |
| **Assay results using patient mRNA without allele-specific quantitation #** | | | | | | |
| **PVS1 Code Weight as per PVS1 flowcharts*** | **Apparent (near) complete splicing** | | **Apparent incomplete impact** | | | **Apparent no impact (relative to controls)** |
| PVS1 | PVS1_Strong (RNA) | | PVS1_N/A (RNA) | | | BP7_Strong (RNA) |
| PVS1_Strong | PVS1_Moderate (RNA) | |  |  |  |  |
| PVS1_Moderate | PVS1_Moderate (RNA) | |  |  |  |  |
| PVS1_Supporting | PVS1_Moderate (RNA) | |  |  |  |  |
| $ Application of weights must also consider other factors relating to assay design. These may include the following: primer and/or construct design that enables detection of naturally occurring isoforms, or detection of aberrations that may be expected based on cryptic site prediction; sensitivity of the methodology to detect transcripts of different size ranges; approaches to measure allele-specific quantitation (sanger sequencing, snapshot etc.); possible amplification bias of shorter products; use of nonsense-mediated decay (NMD) inhibitors, and quantitation of PTC transcripts predicted to lead to NMD. See Appendix for justification for use of these operational thresholds. | | | | | | |
| * Baseline PVS1 code weight is determined by interpretation of the transcript profile from available splicing assay(s) against PVS1 decision trees (See Table 4 searchable excel, and Appendix D) | | | | | | |
| ^ Assumed functional transcript: in-frame and not affecting a critical protein domain, initiation or termination codon; expressed as % per-allele expression. If multiple transcripts with different predicted effect observed – apply weight considering total % per-allele expression of (potentially) functional transcripts. When assessing mRNA from patient material for variants that lead to expression of one or more naturally occurring isoforms, an inferred per allele exclusion level is calculated as follows: Two times level of aberration/s observed in the variant carrier minus the average level of aberration/s in multiple samples of the same type, from control individuals. | | | | | | |
| # Patient allele-specific OR patient not allele-specific plus minigene to quantify | | | | | | |
| ## Interpretation of mRNA assay data without quantitation is based on consensus curator judgement. | | | | | | |

#### PS3 and BS3 code use for assays of protein function

Supplementary Tables 4-5 together summarise the sources of functional assay results used to calibrate code strength for PS3 and BS3. These assays were selected for the calibration dataset since they were considered reliable for use in *BRCA1/2* variant interpretation by members of the ENIGMA Functional Working Group, and results from the assays selected (Supplementary Table 4, Supplementary Table 5) were previously been shown to be strongly predictive of variant pathogenicity using likelihood ratio (LR) estimation with known (likely) benign and (likely) pathogenic variants – classified without use of functional data - as reference sets (Parsons et al 2019, PMID: 31131967). At this point in time results from yeast assays have been excluded as a source for variant classification. Review of relevant publications (Supplementary Table 6) highlighted that most studies assayed only a small number of control variants, the single larger-scale proof-of-principle study included only variants previously classified as pathogenic and did not include a validation set. The conclusion was that currently available yeast-based assay results would not provide any information of value over and above existing mammalian-based assay results.

The results here present an expanded analysis using a larger set of reference controls. As previously, results for each variant were collated across multiple variant assays, to provide a conservative representation of variant effect. Variants predicted to alter splicing and those affecting the p.M1 codon were excluded. LR estimations and derived code strengths are shown in Table 10. The LR based on complete impact in at least one study (and no conflict) provided Strong evidence for pathogenicity. No impact in at least one study (and no conflict) provided Strong evidence against pathogenicity. Results were not materially different when assay results for *BRCA1* and *BRCA2* were considered separately. See footnote of Table 10 for further explanation. Although calibration was done against missense substitution variants (drawn from Supplementary Table 4), functional assay calibration is considered relevant to variants with alternative predicted impact (stop gain, intronic or exonic splice-impacting, synonymous) for such variants with results from assays designed to capture impact via mRNA and protein effect.

The expectation is that the datasets to be used for assessing impact in function will be updated over time, as new relevant studies are published, and re-assessed for individual and combined correlation with clinical features, following similar approaches used here.

As noted in Figure 9, assignment of PS3 or BS3 from “protein” functional assay results must consider potential impact on mRNA splicing (based on bioinformatic and/or mRNA assay results), and the functional assay design - some studies assay of effect on protein only, while others may also capture cellular functional effect due to altered mRNA levels or splicing. To reiterate, for exonic missense/in-frame variants within (potentially) clinically important functional domains, protein functional results are required to confirm no protein impact before BP7_Strong (RNA) can be applied for splicing assay results that demonstrate no aberrant transcript profile; in the absence of such protein functional data, results should be noted in the VCI descriptive text without any code weight applied for the splicing data.

##### **Table 10: Estimation of LR towards pathogenicity based on functional effect reported for missense substitution variants classified as (Likely) Benign or (Likely) Pathogenic using multifactorial likelihood analysis**

|  | (Likely) Benign | | (Likely) Pathogenic† | | LR towards Pathogenicity | (95% CI) | ACMG code strength |
| --- | --- | --- | --- | --- | --- | --- | --- |
|  | n | % | n | % |  |  |  |
| ***LR for BRCA1/2 combined:*** | | | | | | | |
| BRCA1/2 Protein functional effect |  |  |  |  |  |  |  |
| None | 118 | 95.16 | 0 | 2.13 | **0.022**‡ | (0.003-0.16) | **Strong B** |
| None/Partial | 4 | 3.23 | 0 |  | NA |  |  |
| Partial | 1 | 0.806 | 0 |  | NA |  |  |
| Partial/Complete | 0 |  | 3 | 6.38 | NA |  |  |
| Complete | 1 | 0.81 | 44 | 93.62 | **108.69** | (15.44-765.14) | **Strong P** |
| *Total* | *124* |  | *47* |  |  |  |  |

| ***LR for BRCA1 and BRCA2 separately:*** | | | | | | | |
| --- | --- | --- | --- | --- | --- | --- | --- |
| BRCA1 Protein functional effect |  |  |  |  |  |  |  |
| None | 81 | 96.43 | 0 | 2.86 | 0.0296‡ | (0.004-0.205) | **Strong B** |
| None/Partial | 3 | 3.57 | 0 |  | NA |  |  |
| Partial | 0 | 0 | 0 |  | NA |  |  |
| Partial/Complete | 0 |  | 3 | 8.57 | NA |  |  |
| Complete | 0 | 1.19 | 32 | 91.43 | 76.8 | (10.92-540.31) | **Strong P** |
| Total | 84 |  | 35 |  |  |  |  |
| BRCA2 Protein functional effect |  |  |  |  |  |  |  |
| None | 37 | 92.5 | 0 | 8.33 | 0.090‡ | (0.014-0.59) | **Moderate > Strong**‡ |
| None/Partial | 1 | 2.5 | 0 |  | NA |  |  |
| Partial | 1 | 2.5 | 0 |  | NA |  |  |
| Partial/Complete | 0 |  | 0 | 0 | NA |  |  |
| Complete | 1 | 2.5 | 12 | 100 | 40 | (5.74-277.06) | **Strong P** |
| Total | 40 |  | 12 |  |  |  |  |
| † Excludes missense variants predicted to alter mRNA splicing. Functional impact codes were assigned based on effect description as originally published. See Supplementary Table 4 for more details.  ‡ % is calculated assuming a single variant in this category, and thus provides a very conservative estimate of the LR. Recalculation assuming a 1% frequency for categories where no variant is observed yields these LRs: BRCA1/2 combined no functional effect, LR 0.011 (Strong Benign); BRCA1 no functional effect, LR 0.0001 (exceeds Strong Benign); BRCA1 complete functional effect, LR 91.43 (Strong Pathogenic); BRCA2 no functional effect, LR 0.01 (Strong Benign). | | | | | | | |

### Appendix F: PS4 and PS4_moderate proband counting

As per published recommendations (Spurdle et al 2019, PMID:30962250), we categorize breast cancer risk levels associated with a given variant, relative to the general population risk, as follows: **High increased risk,** >4-fold; **Moderate increased risk,** 2-4-fold; **Low increased risk,** greater than unity and <2-fold.

#### **PS4 case-control relative risk or odds ratio for (suspected) high risk variants.**

Case-control results may be applied as **strong** level of evidence in favour of pathogenicity, **consistent with a high increased risk cancer** if the following criteria are met:

- prevalence of the variant in affected individuals is significantly increased compared with the prevalence in ethnicity and country-matched controls.
- relative risk or odds ratio is ≥4.0, and the confidence interval around the estimate does not include 2.0.

Replication of findings for individual variants is strongly encouraged. Further, **should individual level data be applicable to analysis using the case-control likelihood ratio approach, such results should be applied in preference to the case-control OR/RR approach.** The case-control LR approach measures age-specific presentation expected for a pathogenic variant versus a non-pathogenic one, can be converted to an evidence weight for or against pathogenicity using LR cutoffs as in Table 1, and has improved performance to provide weighted evidence for rare variants that do not meet statistical significance using a case-control approach (Zanti et al, in preparation, methodology adapted from that described in Parsons et al 2019, PMID: 31131967). Such LRs should be incorporated into a combined clinical code weight for (or against) pathogenicity under codes PP4 or BP5. See Appendix B for more details.

#### PS4 case-control relative risk or odds ratio for (suspected) moderate risk variants

We considered the interpretation of case-control findings to support a PS4 code for an individual (suspected) moderate risk allele. We documented from a single large-scale breast cancer case-control analysis (Shimelis et al 2017, PMID: 28283652) variants that reached a nominal level of significance (P≤0.05), and the OR ≥2, and noted additional evidence relevant for variant interpretation. The intent was to determine what additional information might be required to give confidence that the OR estimate/P value from a single – albeit large-scale – study, can be considered sufficient to apply the PS4 code. A total of 9 variants were screened in Caucasians (8 variants detected) and Asians (6 variants detected). Bonferroni correction may possibly be interpreted as 0.05/8 (0.00025) for the Caucasian analyses. Results are summarized in Table 11. The findings show need for caution before applying a PS4 code for single rare variants reported to have relative risk or odds ratio of > 2.0 but <4.0 based on a single study. **From results to date, we cannot provide clear recommendations regarding the use of case-control data for reliably informing pathogenicity of individual rare *BRCA1* or *BRCA2* variants associated with moderate risk OR >2.0.**

However, we note for information that the following factors may be considered to inform annotation of a variant as (suspected) hypomorph/moderate risk allele: Independent replication of association (OR >2.0, P<0.05); Supporting segregation studies demonstrate evidence of reduced penetrance, with some stipulation on confidence for this evidence; Multiple independent assays showing variant impacts function with effect intermediate between pathogenic and wildtype controls; allele frequency inconsistent with a high-risk variant (may meet BA1 or BS1 code).

##### Table 11: Variants assessed as potential moderate-risk from case-control analysis

| **Gene** | **Variant** | **n/41890 NFE cases** | **n/41607 NFE controls** | **OR**  **(95% CI)**  **NFE** | **P** | **n/41890 Asian cases** | **n/41607 Asian controls** | **OR**  **(95% CI) Asian** | **P** | **Other information relating to risk** |
| --- | --- | --- | --- | --- | --- | --- | --- | --- | --- | --- |
| *BRCA1* | c.5096 G>A p.Arg1699Gln | 16 | 4  (MAF 0.00009, **almost BS1**) | 4.3  (1.43-12.85) | 0.009 | 0 | 0  (MAF 0, **meets PM2**) | N/A |  | Proven reduced penetrance allele PMID:22889855 PMID:28490613 |
| *BRCA2* | c.7522G>A p.Gly2508Ser | 0 | 0 | N/A |  | 31 | 12  (MAF 0.00029, **meets BS1**) | 2.7  (1.37-5.23) | 0.004 | Not replicated: BRIDGES OR 0.82 (0.45-1.48; P=0.507) (personal communication, Soo Teo) |
| *BRCA2* | c.8187G>T p. Lys2729Asn | 3 | 1  (MAF 0.00002, < BS1, **no code applied**) | 2.8  (0.29-27.64) | 0.368 | 164 | 128  (MAF 0.003, **meets BA1**) | 1.4  (1.12-1.78) | 0.004 | Observed in 96 cases and 87 controls in BRIDGES, almost all Asian (personal communication, Douglas Easton) |
| *BRCA2* | c.9104A>C p.Tyr3035Ser | 18 | 7  (MAF 0.00017, **meets BS1)** | 2.5  (1.05-6.05) | 0.038 | 3 | 0  (MAF 0, **meets PM2)** | 6.95  (0.36-134.62) | 0.200 | PMID: 22889855 reports segregation RR 14.8 (2.4-20), optimal cumulative penetrance ~0.75 relative to *BRCA2* PTC variants. Variant co-occurs with *BRCA2* S1882X in Pedigree G, breast cancer 36y/42y, no other features noted. |

#### PS4_moderate proband counting

The original ACMG/AMP publication (Richards et al 2008, PMID: 25741868) describes use of PS4_moderate code as follows: in instances of very rare variants where case–control studies may not reach statistical significance, the prior observation of the variant in multiple unrelated patients with the same phenotype, and its absence in controls, may be used as moderate level of evidence.

We have carried out extensive analysis to assess the utility of the proband counting method as evidence towards pathogenicity. These findings have been presented to members of the ClinGen SVI, to ClinGen *BRCA12* VCEP members and also ENIGMA members at the June 2022 Scientific meeting.

Based on our results, current recommendations for PS4 in relation to proband-counting rule code are as follows:

- PS4 proband counting method requires calibration
- Separate data analysis is required for each dataset to determine PS4 weight.
- Application of PS4_moderate overlaps with code PM2 (absence in controls), thus only one of these two codes should be considered for a variant interpretation

In summary, our findings indicate it would be unwise to stipulate specific criteria for *BRCA1/2* for generic use at this time.

A more extensive analysis that captures personal and family history of cancer (and potentially age at onset and tumour features) is more appropriate. LRs from such analyses have previously been applied only in the context of multifactorial likelihood analysis, but we propose to use such LRs to assign a clinical code weight for (or against) pathogenicity, and may be applied under codes PP4 or BP5. See Appendix B for further details.

### Appendix G: Population frequency data - PM2_Supporting, BA1, BS1

We recommend that assignment of population frequency-based codes should consider assessment of read-depth, sequencing technologies used, and variant type e.g. single nucleotide substitution versus insertion-deletion (Davidson et al 2021, PMID: 33600021). Currently we recommend use of read depth >25 for application of PM2_Supporting, and read depth >20 for assignment for benign codes (BA1, BS1). We do not recommend use of PM2_Supporting for insertion-deletion variants due to the low recall estimates for variants of this type (PMID: 33600021).

#### PM2_Supporting – Absence/rarity in controls.

Using the approach as described in Parsons et al 2019 (PMID: 31131967), we estimated the LR towards pathogenicity based on maximum population allele frequency observed in gnomAD (genome and exome data combined) for a set of 749 variants (all with MAF <0.01 in outbred populations) that have previously been classified as (likely) benign or (likely) pathogenic using multifactorial likelihood analysis (as used for PS4 proband counting analysis above, See Supplementary Table 7). The maximum allele frequency observed across gnomAD non-founder populations (Non-Finnish European, African, Latino, East Asian, South Asian) was assigned for a given variant. Counts and LR estimations for different frequency categories are shown in Supplementary Table 8. Code PM2 (variant absent in controls) was estimated to meet Supporting weight: there were 175/609 (likely) benign variants (28.7%) versus 116/140 (likely) pathogenic variants (82.9%), resulting in an estimated LR towards pathogenicity of 2.88 (95% CI 2.49-3.34). Observation of a variant only once in a gnomAD outbred population was not informative: for this group, there were 71/609 (likely) benign variants (11.7%) versus 16/140 (likely) pathogenic variants (11.4%), LR 0.98 (95% CI 0.59-1.63). Likewise, maximum MAF >0 to ≤0.00002 was not informative.

#### BA1 – Stand-alone allele frequency.

BA1 frequency was calculated using the Whiffin calculator (<http://cardiodb.org/allelefrequencyapp/>), with the following assumptions:

- Prevalence (population-based breast cancer): 0.125 (1: 8)
- Genetic Heterogeneity: 0.01
- Allelic Heterogeneity: 1.00
- Penetrance: *BRCA1* - 0.72; *BRCA2* - 0.69 (estimate from prospective study, PMID:28632866)

Using these inputs, the estimated minimal credible allele frequency is 0.000868 (*BRCA1*) and 0.000906 (*BRCA2*), which were both rounded up to 0.001 for simplicity of application.

Results from the LR-based approach (Supplementary Table 8) were consistent, with LR 0.0009 (Benign Stand-alone) for MAF >0.001 to ≤ 0.01.

#### BS1 – Allele frequency greater than expected for disorder.

BS1 frequency was calculated using the Whiffin calculator (<http://cardiodb.org/allelefrequencyapp/>), with the following assumptions:

- Prevalence (population-based breast cancer): 0.125 (1: 8)
- Genetic Heterogeneity: 0.01
- Allelic Heterogeneity: 0.10
- Penetrance: *BRCA1* - 0.72; *BRCA2* - 0.69 (estimate from prospective study, PMID:28632866)

Using these inputs, the estimated minimal credible allele frequency is 0.0000868 (*BRCA1*) and 0.0000906 (*BRCA2*), which were both rounded up to 0.0001 for simplicity of application.

Results from the LR-based approach (Supplementary Table 8) were consistent, with LR 0.03 (Benign Strong) for MAF >0.0001 to ≤ 0.001. In addition, the LR-based approach indicated that ACMG/AMP code **BS1_Supporting may conservatively be applied for MAF >0.00002 to ≤ 0.0001.**

As per SVI recommendations, we suggest conservative application of BA1 and BS2 codes using filter allele frequency (and not minor allele frequency), and further that this is based on highest filter allele frequency in a non-founder population from gnomAD (TCGA data excluded), assessing exome and genome data separately. Do not apply BA1 or BS1 codes to well-established pathogenic founder variants.

### Appendix H: Biallelic *BRCA1* or *BRCA2* Alterations (PM3 or BS2)

Fanconi Anemia Type D1 (FA-D1), a recessive condition due to biallelic inheritance of *BRCA2* loss-of-function alleles, is largely reported in the literature as a very early onset, severe disorder with specific clinical features. Fanconi Anemia Type S (FA-S), due to biallelic inheritance of *BRCA1* loss-of-function alleles, was presumed to be embryonically lethal based on findings from mouse studies. However, a limited number of examples reporting co-occurrence of *BRCA1* alleles designated as pathogenic for breast-ovarian cancer predisposition in patients with molecular features of Fanconi anemia (FA) suggest that particular combinations of alleles may escape lethality.

Recommendations for application of codes PM3 and BS2 for *BRCA1* and *BRCA2* are described below. These recommendations were informed by previous review of clinical features reported in the literature for FA-D1 patients (n=57 and FA-S patients (n=8), summarized in Supplementary Tables 9 and 10. We note that it has been argued that alleles identified in FA-D1 and FA-S are associated with reduced penetrance compared to the average truncating *BRCA1* or *BRCA2* allele. However, there has been no comprehensive study specifically addressing this hypothesis, and (i) the majority of alleles identified in FA-D1 patients to date have been reported as pathogenic in the context of breast-ovarian cancer (ii) the vast majority of variants reviewed to derive PM3/BS2 specifications are considered high-risk pathogenic for breast-ovarian cancer based on ENIGMA classification criteria V 2.5.1 (June 2017). **The clinical presentations required to meet PM3 or BS2** were derived from examination of the available data to **represent severe FA presentation expected for co-occurrence of two “high-risk” pathogenic variants** (risk equivalent to the average protein termination codon variant).

#### Application of PM3.

The application of PM3 is a modification of SVI recommendation for PM3 - V 1.0 approved May 2, 2019. To be applied for co-occurrence of a *BRCA1* or *BRCA2* VUS *in trans* with a known (likely) pathogenic variant in the same gene, if identified in a patient with *BRCA1* or *BRCA2* related FA features.

Overall prevalence of FA is reported as 1/160,000 (Orphanet). *BRCA2* (FA-D1) accounts for 3% of all FA, so the expected prevalence is 1/5.3M. There are no figures for *BRCA1*-related FA, so the prevalence has to be lower. Specifications for features of an FA-affected patient were adapted from definitions from GeneReviews (last revision June 3, 2021). See below for more details.

**For co-occurrence of *BRCA1* or *BRCA2* variants, consider an individual to have phenotype consistent with *BRCA1*- or *BRCA2*-related FA if:**

(i) Increased chromosome breakage and radial forms on cytogenetic testing of lymphocytes with diepoxybutane (DEB) and/or mitomycin C (MMC) or evidence of spontaneous chromosome breakage, plus **at least one clinical feature indicative of *BRCA1/2*-related FA** that is encapsulated under one of *the three categories described below: Physical features, Pathology findings, Cancer diagnosis ≤5yr*.
(ii) Result unknown for DEB/MMC chromosome breakage test or spontaneous chromosome breakage, and at least two **clinical features indicative of *BRCA1/2*-related FA** that are encapsulated under at least two of the three categories described below: *Physical features, Pathology findings, Cancer diagnosis ≤5yr*.

**Clinical features** indicative of FA:

- ***Physical features*** (in ~75% of affected persons), include: prenatal and/or postnatal short stature; abnormal skin pigmentation (e.g., café au lait macules, hypo-pigmentation); Skeletal malformations (e.g., hypoplastic thumb, hypoplastic radius); Microcephaly, triangular face; Ophthalmic anomalies; Genitourinary tract anomalies. See Orphanet (<https://www.orpha.net/consor/cgi-bin/Disease_HPOTerms_Simple.php?lng=EN>) for full list of >100 HPO terms (and their reported frequency).
- ***Pathology and laboratory findings*** *(non-cancer related)* include: inordinate toxicities from chemotherapy or radiation, and also progressive bone marrow failure (*unrelated to cancer treatment*), aplastic anemia, myelodysplastic syndrome, macrocytosis, cytopenia (especially thrombocytopenia, leukopenia, and neutropenia), and increased fetal haemoglobin (often precedes anemia). Note: FA patients with very early onset onset cancer (***≤5yr***) may not present with hematologic disease, which is reported to have median age at onset of 7 years in FA patients in general (Wagner JE, MacMillan ML, Auerbach AD. Hematopoietic cell transplantation for Fanconi anemia. In: Blume KG, Forman SJ, Appelbaum FR, eds. Thomas’ Hematopoietic Cell Transplantation. 2003.)
- ***Cancer diagnosis ≤5yr*.** This age cut-off was selected as being considered sufficiently conservative to exclude PM3 assignment for hypomorphic *BRCA1* or *BRCA2* variants. Literature review (See Supplementary Tables 9, 10) identified the following cancer types in individuals with *BRCA1*- or *BRCA2*-related FA: *BRCA2* **–** acute myeloid leukemia (AML) (14), medulloblastoma (12), nephroblastoma (Wilms tumour)(13), acute lymphoblastic leukemia (ALL (3, T-cell, B-cell), leukemia unspecified (2), neuroblastoma (4), neuronal neoplasm (1), neuroepithelial (1), brain (2) , astrocytoma (1), glioblastoma (1), rhabdomyosarcoma (3), hepatoblastoma (1), kidney sarcoma (1), unknown intra-abdominal malignancy (1); *BRCA1* – ALL (1, T-cell), neuroblastoma (1). The cancer types denoted (number of cases in parentheses) are those identified in *BRCA1/2-*related FA cases reported with diagnosis ≤5yr in the literature to date (Supplementary Tables 9 and 10). Cancer diagnosis ≤5yr was selected as indicative of severe cancer phenotype associated with classical highly penetrant *BRCA2* pathogenic variants based on a literature review and analysis. Of 55 individuals reported in the public domain to have Fanconi Anemia due to homozygote (1 individual) or compound heterozygous carrier status for *BRCA2* pathogenic alleles, and sufficient clinical data to permit assessment, 47/55 (85%) exhibited a severe cancer phenotype – defined as first cancer ≤5y (Supplementary Table 9). Of 8 individuals reported to have Fanconi Anemia due to compound heterozygote carrier status for *BRCA1* alleles, 2/6 individuals with two ‘high-risk’ pathogenic alleles had developed cancer ≤5y (Supplementary Table 10). *Also see caveat below.*

***Caveat:** individuals carrying hypomorphic *BRCA1* or *BRCA2* variants (homozygote, or compound heterozygote with a pathogenic high-risk allele) may not fit criteria for phenotype-confirmed FA. See PMID: 31347298, describing a patient carrying *BRCA1* founder c.181T>G (p.Cys61Gly) and known reduced penetrant hypomorphic variant c.5096G>A (p.Arg1699Gln). The patient developed breast cancer at age 30y, had some FA features (short stature, microcephaly, triangular face, skin hyperpigmentation), experienced significant toxicity to chemotherapy, but chromosome fragility was not present in patient-derived peripheral blood lymphocytes. Recommend complementation testing to confirm FA subtype.

If a variant co-occurs with a P or LP variant in the same gene, or occurs as a homozygote, in a patient with **phenotype consistent with *BRCA1*- or *BRCA2*-related FA**, award each proband a point value as per Table 12. ***For related individuals, score only most severe presentation****.*

##### Table 12: PM3-related points assigned to an individual with co-occurrent variants and FA phenotype:

| **Presentation of proband with variant under assessment*** | **Points per *BRCA1*-related (FANCS) or *BRCA2*–related (FANCD1) Fanconi Anemia patient** | |
| --- | --- | --- |
|  | **P or LP *in* *trans***  **or homozygote**** | **Phase unknown** |
| Phenotype consistent with *BRCA1*- or *BRCA2-*related FA | 2 | 1 |

* Variant under assessment must be sufficiently rare (meet PM2_Supporting, or PM2 not applicable). Consider other gene panel test results as potential explanation for phenotype.

** Co-occurrent P or LP variant should be assigned classification using VCEP specifications. Stipulation for *in trans* can be met by genotyping of at least one parent, or assumed if VUA is seen with at least 2 different P/LP variants. Further, co-occurrence *in trans* can be inferred in the case of a homozygote FA-affected patient due to a consanguineous union, or if both maternal and paternal lineages present with family history of cancer consistent with *BRCA1* or *BRCA2* related cancers.

**Assign final PM3 code based on sum of PM3-related points from *observations across multiple unrelated individuals*,** as follows:

PM3_Strong = 4 (or more) points; PM3 = 2 points; PM3_Supporting = 1 point.

#### Application of BS2.

A single observation of a *BRCA1* or *BRCA2* VUS co-occurring *in trans* with a pathogenic variant in the same gene equates to a co-occurrence LR of 0.04, assuming the test sample set has a detection rate of 0.025 for Pathogenic variants in that gene (calculation from Goldgar et al 2004, PMID: 15290653). This meets the odds range (0.00285 to 0.05), Benign Strong category (Tavtigian et al 2018, PMID: 29300386, also see Table 1). While it is optimal that a patient is assessed to exclude FA features, a patient with age at cancer diagnosis/last follow-up that exceeds ages reported for *BRCA1*-related or *BRCA2*-related FA is considered very unlikely to have undiagnosed FA due to two highly penetrant alleles (see PM2 above). Applying such assumptions eliminates need for formal assessment of FA features in large datasets providing co-occurrence information.

If a variant is detected in *trans* with a P or LP variant, or as a homozygote, in a (presumed) **FA**-**unaffected** patient, award each proband a point value as per chart below (Table 13). This scheme allows for different levels of confidence in assigning absence of FA phenotype based on age at diagnosis or follow-up, where there has been no formal assessment of molecular features (DEB or MMC chromosome breakage test) or clinical features of FA. Final code assignment is based on the sum of points from observations across multiple individuals.

##### Table 13: BS2-related points assigned to an individual with co-occurrent variants and unlikely FA phenotype:

| **Proband presentation** | **Points per proband meeting age/cancer diagnosis criteria** | | |
| --- | --- | --- | --- |
|  | **VUA *in* *trans* with P or LP in same gene *** | **Homozygote**** | **Phase unknown, co-occurs with P or LP in same gene** |
| Patient first cancer onset >50y or cancer-unaffected at age at last follow-up of >50y | 4 | 2 | 1 |
| Patient first cancer onset 40-50y or cancer-unaffected at age of last follow-up of 40-50y | 2 | 1 | 0.5 |

* VUA = Variant Under Assessment. VUA should not be bioinformatically predicted (or experimentally proven) to have a clinically important effect on protein or mRNA splicing. Co-occurrent P or LP variant should be assigned classification using VCEP specifications. Stipulation for *in trans* can be met by genotyping of at least one parent, or assumed if VUA is seen with at least 2 different P/LP variants.

** Apply only for phenotyped individuals from clinical or research cohorts. NOT to be applied for data used to assign frequency-based codes; we have observed that variants observed as ≥2 homozygotes in GnomAD data (assumed unaffected adults of unspecified age) are already captured by the BA1 or BS1 frequency codes.

**Final BS2 code assignment, based on sum of BS2-related points:**

BS2 = 4 (or more) points; BS2_moderate = 2 points; BS2_Supporting = 1 point.

*.*

### Appendix I: Co-segregation Analysis (PP1 and BS4)

Quantitative co-segregation analysis methods are well established for use in interpretation of causality of *BRCA1* and *BRCA2* variants. The preferred method is a Bayesian analysis that considers both likelihood of causality and likelihood of non-causality of the variant in relation to age-specific penetrance of disease phenotypes relevant for *BRCA1* or *BRCA2*, given the background population prevalence of these disease types (Thompson et al 2003, PMID: 12900794). Unlike meiosis counting methods, the Bayes score directly measures the strength of evidence across all available family evidence, including probabilistic information from untested individuals, and accounting for ascertainment due to proband cancer status.

An online tool for application of this method is available via

<https://fengbj-laboratory.org/cool3/analysis.html>.

Information about pedigree formatting, and the penetrance tables relevant for *BRCA1* and *BRCA2* are available at <https://fengbj-laboratory.org/cool3/manual.html>.

The Bayes score derived from multiple families with the same variant can be combined multiplicatively to generate a combined LR for segregation. This Bayes score is generally used in quantitative multifactorial likelihood analysis approaches (see **Appendix B**). However, the scores can be converted to different PP1 or BS4 strength categories, following the LR thresholds derived from Tavtigian et al 2018 (PMID: 29300386), namely:

- Pathogenic Very Strong: LR≥350:1
- Pathogenic Strong: LR≥18:1
- Pathogenic Moderate: LR≥4:1
- Pathogenic Supporting: LR≥2:1
- Benign Strong: LR≤0.053:1
- Benign Moderate: LR ≤0.23:1
- Benign Supporting: LR ≤0.48:1

**Stipulation**: to apply code as Pathogenic Very Strong, VUS should have bioinformatically predicted (or experimentally proven) effect on protein or mRNA splicing. Further, if co-segregation score is from a single family, or several families from an isolated population, then it is necessary to assess and exclude the possibility of other causative pathogenic variants in the same gene. To apply code as Benign Very Strong, assess the possibility of bi-linearity to explain negative co-segregation.

### Appendix J: Bioinformatic Analysis and Code Application - incorporating PM1, PP3, BP1, BP4, BP7, and PS1

Bioinformatic predictions can be calibrated against clinical information to derive a prior probability of pathogenicity. This information can also be translated into a LR and ACMG code strength. In addition, a LR towards pathogenicity can be estimated for bioinformatic prediction of variant effect due to a missense substitution, using protein functional assay results as a surrogate for pathogenicity.

For splicing predictions, our recommendations are based on results from unpublished analyses conducted by the SVI Splicing subgroup. Calibration of predictors against functional and splicing assay results showed that the SpliceAI tool (Jaganathan et al 2019, PMID: 30661751) outperformed multiple other splicing predictors (including MaxEntScan). For variants outside of the donor/acceptor ±1,2 positions, the optimal Splice AI cutpoints (based on the maximum ∆ score of donor loss, acceptor loss, donor gain or acceptor gain) to predict spliceogenicity were:

- ≤0.1 for BP4 for no predicted splicing impact
- >0.1 and <0.2 for PP3 and BP4 not met for predicted splicing (not informative)
- ≥0.2 for PP3 for predicted splicing impact

We do not recommend inclusion of evolutionary conservation in assessment of silent or intronic variants, based on the following: there is no logic to expect nucleotide conservation at positions where a variant leads to a gain of a splicing motif (donor, acceptor or exonic or intronic splicing enhancers); published evidence from an analysis of 27,733 variants within or adjacent to 2,198 human exons shows that lack of conservation has limited predictive power to detect non-splice disrupting variants (Cheung et al 2019, PMID: **30503770).**

To inform bioinformatic code use for missense prediction, we carried out analysis to determine the predictive capacity of “location inside or outside domain” for a missense variant, and to compare the utility of several missense prediction tools to provide evidence for or against pathogenicity. We also reviewed findings from the literature to inform code weights for variants according to domain and broad effect (missense, silent/synonymous, intronic).

#### Analyses and results justifying use and weight of Bioinformatic and Domain codes for BRCA1/2 missense variant interpretation

***The following tools and annotations were applied:***

- A-GVGD  – missense alterations, scores assigned as per website (<http://agvgd.hci.utah.edu/>)
- BayesDel – missense alterations, including in-frame indels, scores assigned using the PERCH package (<http://fengbj-laboratory.org/lab/index.html>). Scores were categorized in several ways – generic recommendation from tool creator Bing Feng (<0.3, ≥0.30), or as designated in two publications comparing predictors (Tian et al 2019, PMID: 31484976; Hart et al 2019, PMID: 29884841). The web tool (<https://gwiggins.shinyapps.io/lr_shiny/>) was then used to compare the sensitivity and specificity of different BayesDel cutpoints across three categories.
- NVM - missense alterations, score categories drawn from the publication (Hart et al 2019, PMID: 29884841; Table S2 from this publication, Annotation of all possible BRCA1 and BRCA2 missense variants by function and in silico prediction models).
- Annotations to exclude potentially spliceogenic missense variants were based on prior probability derived from MaxEntScan scores (<http://priors.hci.utah.edu/PRIORS/>, See below for details.

##### Method 1: Heterogeneity analysis, using family history as a predictor of pathogenicity

The analysis approach was similar to that used previously to estimate prior probability of variant pathogenicity based on clinical features of a dataset of individuals undergoing BRCA1 and BRCA2 diagnostic testing  (Easton et al 2007, PMID: 1794331; Tavtigian et al 2008, PMID: 18951461; Vallee et al 2016, PMID: 26913838; Li et al 2020, PMID: 31853058). The results presented here are from re-analysis of data used for the Li et al study (PMID: 31853085); the dataset included >138,000 Individuals undergoing MGPT (5-49 genes) from March 2012 to December 2016, with clinical history information obtained from test requisition forms, and no (likely) pathogenic variant in another gene (*ATM, PALB2, BARD1, CDH1, CHEK2, MLH1, MSH2, MSH6, NBN, NF1, PTEN, RAD51C, RAD51D, TP53*).  Results from the model analysis were used to calibrate two *in-silico* predictors of variant impact via effect on protein function (missense substitutions, and additionally in-frame in-dels for BayesDel), and with redefinition of (potentially) clinically important functional domains.  Heterogeneity analysis was used to estimate the proportion of pathogenic variants (of those designated as likely pathogenic or VUS) that are likely to be clinically significant as a function of bioinformatically predicted classifications, and variant location in a domain. Silent substitution variants were not annotated, so it was not possible to estimate prior probability of pathogenicity for this subset. Table 14 shows prior probabilities arising from analysis. The results show convincingly that missense variation *in toto* does not contribute much to *BRCA1/2* related disease (10% of variants predicted to be pathogenic), with the signal limited to variants inside key domains (35% of variants, compared to 1% outside domains). BayesDel outperformed A-GVGD for prediction of missense pathogenicity within key domains (74% of variants with BayesDel ≥0.3 versus 59% for the top A-GVGD tier C65). Missense variants outside a key functional domain had low prior, irrespective of BayesDel prediction.

##### Table 14: Prior probability of pathogenicity for BRCA1 and BRCA2 missense variants according to functional domain and predicted effect

| **Bioinformatic Group** | **# Variants*** | **Prior Probability of Pathogenicity**  **(Confidence Interval)** |
| --- | --- | --- |
| All missense variants | 2007 | 0.10 (0.07-0.14) |
| *Missense in key domain*** | 501 | 0.35 (0.26-0.45) |
| A-GVGD C0 | 277 | 0.23 (0.05-0.41) |
| A-GVGD C15-C25 | 72 | 0.40 (0.02-0.78) |
| A-GVGD C35–C55 | 43 | 0.38 (0.00-0.85) |
| A-GVGD C65 | 109 | 0.59 (0.28-0.90) |
| BayesDel <0.30 | 363 | 0.17 (0.01-0.32) |
| BayesDel ≥0.30 | 138 | 0.74 (0.47-0.10) |
| *Missense outside key domain* | 1507 | 0.01 (0.00-0.04) |
| BayesDel <0.30 | 1459 | 0.007 (0.00-0.03) |
| BayesDel ≥0.30 | 47 | 0.06 (0.00-0.37) |
| * Includes Likely pathogenic and VUS missense only. Amino acid substitution variants also predicted to potentially disrupt splicing (based on MaxEntScan predictions, <http://priors.hci.utah.edu/PRIORS/>, moderate-high prior probability to disrupt a native donor or acceptor), were excluded.  **Key (potentially) clinically important functional domains, used in this analysis, were defined as follows: BRCA1 RING aa 2-101, BRCA1 coiled-coil aa 1391-1424, BRCA1 BRCT repeats aa 1650-1863, BRCA2 PALB2 binding domain aa 10-40, BRCA2 DNA binding aa 2481-3186. Also see Tables 3 and 4. | | |

##### Method 2: Likelihood ratio estimation, using protein functional assay results as a proxy for pathogenicity assertions.

Using the initial functional assay dataset collated for use in *BRCA1/2* expert panel variant curation, variants with Functional impact (complete) were assigned to an “assumed pathogenic” reference set, and variants with no functional impact were assigned to an “assumed benign” reference set (Supplementary Table 11). LR estimations were derived as described in Parsons et al 2019 (PMID: 31131967), by comparing the proportion of “assumed pathogenic” versus “assumed benign” variants that fell into a given bioinformatic category. Results are detailed in Supplementary Table 12, and summarized below in Table 15.

##### Table 15: Summary LR derivation for bioinformatic predictors of missense variants, using functional assay reference datasets*

| **Tool** | **Category** | **LR towards Pathogenicity *BRCA1*** | **ACMG Code Evidence Strength *BRCA1*** | **LR towards Pathogenicity *BRCA2*** | **ACMG Code Evidence Strength *BRCA2*** |
| --- | --- | --- | --- | --- | --- |
| Align-GVGD | C0 | 0.25 (0.20-0.30) | Supporting Benign | 0.14 (0.06-0.35) | Moderate Benign |
|  | C15-C55 | 2.24 (1.86-2.69) | Supporting Pathogenic | 1.21 (0.71-2.08) | No evidence |
|  | C65 | 10.43 (8.14-13.35) | Moderate Pathogenic | 2.44 (1.59-3.76) | Supporting Pathogenic |
| BayesDel | <0.3 | 0.29 (0.24-0.34) | Supporting Benign | 0.34 (0.22-0.53) | Supporting Benign |
|  | ≥0.3 | 7.75 (6.59-9.10) | Moderate Pathogenic | 10.21 (5.06-20.06) | Moderate Pathogenic |
| BayesDel  Tian cutpoints | ≤0.147 | 0.12(0.09-0.17) | Moderate Benign | 0.10 (0.02-0.24) | Moderate Benign |
|  | Between | 2.42 (2.14-2.74) | Supporting Pathogenic | 2.46 (1.69-3.57) | Supporting Pathogenic |
|  | ≥0.425 | 27.17 (17.21-42.92) | Strong Pathogenic | 15.00 (1.88-119.58) | Strong Pathogenic |
| BayesDel  NVM cutpoints | <0.206 | 0.21 (0.17-0.26) | Supporting Benign | 0.34 (0.22-0.53) | Supporting Benign |
|  | ≥0.206 | 5.05 (4.48-5.68) | Moderate Pathogenic | 6.70 (3.74-11.99) | Moderate Pathogenic |
| NVM_ Predict | 0 (B) | 0.31 (0.26-0.36) | Supporting Benign | 0.34 (0.22-0.52) | Supporting Benign |
|  | 1 (P) | 6.15 (5.31-7.12) | Moderate Pathogenic | 9.04 (4.62-17.69) | Moderate Pathogenic |

*Excludes variants with partial impact or conflicting results between studies, and variants also predicted to potentially disrupt splicing (based on MaxEntScan predictions, <http://priors.hci.utah.edu/PRIORS/>).

Derivation of LRs based on functional assay results show that meta-predictors BayesDel (<3/≥3 cutpoint) and NVM_predict (using 0,1 coding as per publication, Hart et al 2019, PMID: 29884841) perform equally well, and more consistently than other tools/cutpoints. In contrast to heterogeneity analysis, the tool scores provided supporting evidence against pathogenicity, and moderate evidence towards pathogenicity.

Since BayesDel has the advantage of providing scores for in-frame indels, we selected this tool for further analysis to generate cutpoints for three categories, to intentionally designate a middle category that represents the range of scores that are not helpful in predicting impact of a missense substitution and for which bioinformatic codes should not be applied i.e. PP3 or BP4 is not met. Analysis was restricted to reference set variants located within (potentially) clinically important functional domains. At this point, additional functional data was available in the public domain for missense variants (*BRCA1* 2160 variants, *BRCA2* 514 variants; Supplementary Table 13); given the increase in reference set size, analyses were separated by gene. Detailed results are shown in Supplementary Table 14. Threshold selection considered sensitivity, specificity and minimizing proportion of variants that would be assigned no evidence. Small-window analysis was conducted to confirm that a window of scores near the selected cutpoint provided at least supporting evidence for or against inferred pathogenicity. The thresholds selected are thus considered conservative for application of supporting bioinformatic evidence.

For missense variants located in (potentially) clinically important functional domains (currently defined as BRCA1 RING aa 2-101, BRCA1 coiled-coil aa 1391-1424, BRCA1 BRCT repeats aa 1650-1857, BRCA2 PALB2 binding domain aa 10-40, BRCA2 DNA binding aa 2481-3186), the optimal BayesDel cutpoints for the two genes were determined to be:

- BP4 for no predicted missense impact: ≤0.15 for BRCA1, ≤0.18 for BRCA2
- PP3 and BP4 not met for predicted missense impact (not informative): BRCA1 >0.15 and <0.28; BRCA2 >0.18 and <0.30.
- PP3 for predicted missense impact” ≥0.28 for BRCA1, ≥0.30 for BRCA2

##### Evidence from published analyses

We reviewed the evidence for pathogenicity for BRCA1 and BRCA2 missense variants by domain from a recent large-scale case-control study of 60,466 breast cancer cases and 53,461 controls (Dorling et al 2021, PMID: 33471991). The results provided no evidence that missense variants located outside a domain were overall associated with increased risk, with risk estimates as follows: *BRCA1* OR 1.02 (0.94-1.10), *BRCA2* OR 0.97 (0.92-1.03). Given that case-control data may be used to provide strong evidence in favour of pathogenicity (ACMG/AMP code PS4), it seems to logical to assert that these case-control findings provide strong evidence against pathogenicity for missense variants located outside of a protein domain. Indeed, this assertion is consistent with the results of an analysis of ClinGen submitter classifications for *BRCA1* and *BRCA2* variants according to protein domain (Dines et al 2020, PMID: 31911673), which provided evidence that variants in “coldspots” could be assigned a strong benign category based on their location alone.

#### ACMG Bioinformatic and Domain code weights to be used for *BRCA1/2* variant interpretation

Based on the findings described above, we provide in Table 16 below a summary of the recommendations for application of initial code weights based on variant type/location and bioinformatic predictions. In some instances codes are considered not applicable since they are captured by other codes/criteria. Silent substitution (synonymous) variants are categorized by location outside versus inside a domain to align weights with those for missense alterations outside of a domain, and to allow for the small possibility that nucleotide changes within a protein functional domain could potentially affect protein translation rate and folding due to codon usage bias. Current preference is for use of SpliceAI for splicing predictions, and BayesDel for missense and in-frame exonic alterations within a (potentially) clinically important functional domain. Please also see **Appendix E**, Figure 9 for recommendation around **use of bioinformatic codes in the context of mRNA and protein functional data.**

##### Table 16: Overview of recommendations for bioinformatic and domain code weight application

| **AMCG/AMP Code** and criterion description* | **Application for *BRCA1* and *BRCA2* Variant Interpretation**** |
| --- | --- |
| **PM1** Located in a mutational hot spot and/or critical and well-established functional domain (e.g., active site of an enzyme) without benign variation | N/A**.** Informed by clinically calibrated bioinformatic prior probability of pathogenicity for missense alterations – missense predictions only apply if a variant is located in a (potentially) clinically important functional domain.  **Evaluate under BP1, BP4, PP3.** |
| **PM4** Protein length changes as a result of in-frame deletions/insertions in a nonrepeat region or stop-loss variants | N/A. Captured by bioinformatic tool prediction.  **Evaluate under BP1, BP4, PP3.** |
| **PM5** Novel missense change at an amino acid residue where a different missense change determined to be pathogenic has been seen before. | N/A. Some residues can harbor pathogenic and non-pathogenic substitutions.  **Evaluate under BP1, BP4, PP3.** |
| **PP2** Missense variant in a gene that has a low rate of benign missense variation and in which missense variants are a common mechanism of disease | N/A. Missense variants are not a common mechanism of pathogenicity for *BRCA1* and *BRCA2*. |
| **PP3** Multiple lines of computational evidence support a deleterious effect on the gene or gene product (conservation, evolutionary, splicing impact, etc.). | **Apply PP3 for missense or in-frame insertion-deletion inside a (potentially) clinically important functional domain and predicted impact via protein change** (BayesDel ≥0.28 for *BRCA1*, ≥0.30 for *BRCA2*).  **Apply PP3 for predicted splicing** (SpliceAI ≥0.2) **for silent, missense/in-frame (irrespective of location in clinically important functional domain) and for intronic variants outside of the donor and acceptor ±1,2 positions.** |
| **BP1** Missense variant in a gene for which primarily truncating variants are known to cause disease | Informed by clinically calibrated bioinformatic prior probability of pathogenicity, that considers functional domain, missense/ in-frame changes, splicing effects.  **Apply BP1_Strong for silent substitution, missense or in-frame insertion-deletion outside a (potentially) clinically important functional domain”** (no splicing predicted, SpliceAI ≤0.1). Missense prediction not applicable. |
| **BP3** In-frame deletions/insertions in a repetitive region without a known function | N/A. Captured by bioinformatic tool prediction, and domain analysis. |
| **BP4** Multiple lines of computational evidence suggest no impact on gene or gene product (conservation, evolutionary, splicing impact, etc.). | **Apply BP4 for missense or in-frame insertion-deletion inside a (potentially) clinically important functional domain, and no predicted impact via protein change or splicing** (BayesDel ≤0.15 (*BRCA1*) or ≤0.18 (*BRCA2*), AND SpliceAI ≤0.1).  **Apply BP4 for silent variant inside a (potentially) clinically important functional domain, if no predicted impact via splicing** (SpliceAI ≤0.1).  **Apply BP4 for intronic variants outside of the donor and acceptor ±1,2 positions, and no predicted impact via splicing** (SpliceAI ≤0.1). |
| **BP7** A synonymous (silent) variant for which splicing prediction algorithms predict no impact to the splice consensus sequence nor the creation of a new splice site AND the nucleotide is not highly conserved. | Following convention, this code is applied in addition to BP4 (no splicing prediction, Splice AI ≤0.1) to capture the low prior probability of pathogenicity of silent variants. Nucleotide conservation is not considered relevant.  **Apply BP7 for silent variant inside a (potentially) clinically important functional domain, if no predicted impact via splicing** (BP4 met).  **Also apply BP7 for intronic variants located outside of conserved donor or acceptor motif positions (**at or beyond positions +7/-21), if no predicted impact via splicing (BP4 met).******* |

* Original ACMG criteria codes and descriptions are as per Richards et al 2008 (PMID: 18414213).

** Known and potentially clinically important functional domain defined as: BRCA1 RING aa 2-101, BRCA1 coiled-coil aa 1391-1424, BRCA1 BRCT repeats aa 1650-1857, BRCA2 PALB2 binding domain aa 10-40, BRCA2 DNA binding aa 2481-3186.

*** Justification for application of BP7 for intronic variants in this region is based on maximum likelihood estimation analysis of breast cancer case-control data, with results indicating that most rare non-coding variants in *BRCA1* and *BRCA2* (without applying any bioinformatic prediction filters) are benign (James et al 2022, PMID: 35191126); Case-control enrichment analysis limited to deeper intronic variants (Supplementary Table 15) indicate that intronic variant location outside of the splice region (+7 to -21) provides moderate evidence against pathogenicity (even without applying bioinformatic prediction of splicing as a filter).

#### PS1 for predicted missense substitutions and predicted spliceogenic variants

##### PS1 for predicted missense substitution variants

PS1 may be applied for a variant predicted to lead to the same missense substitution as a known pathogenic variant that is due to a different nucleotide change. Use at moderate weight if reference variant is classified as likely pathogenic. Both (likely) pathogenic variant and variant under assessment should have no predicted/confirmed effect on mRNA splicing. (Likely) pathogenic variant should be

assigned classification using VCEP specifications.

##### PS1 for predicted spliceogenic variants

Following recommendations drafted by the ClinGen SVI Splicing subgroup, PS1 may also be applied in the context of exonic and intronic variants based on similarity of predicted mRNA effects for a variant under assessment, in comparison to a known pathogenic variant – following the logic that this code captures existing evidence of pathogenicity for a variant with an identical mechanism of pathogenicity. The predicted splicing event could be loss of the native splice sites or gain/strengthened splice sites, but the predicted event of the known (likely) pathogenic variant must precisely match the event of the variant being classified. See Table 17 below for recommended weights.

Application of the code varies considers if the variant under assessment and the known (likely) pathogenic variant are located outside or inside the donor/acceptor ±1,2 positions (termed donor/acceptor dinucleotide for simplicity), and if within the same splicing motif – at the same nucleotide position or not, and dependent on whether the reference variant is pathogenic or likely pathogenic. For example, the code is applied at full strength for a variant located outside the donor/acceptor ±1,2 positions with a similar predicted impact on the same mRNA splicing event as another established pathogenic variant at the same nucleotide, and the weight is reduced to moderate level if the reference variant is classified as likely pathogenic.

PS1(Splicing) may also be applied for a variant located at the donor/acceptor dinucleotide and for which there is another established (likely) pathogenic variant located within the same splice motif (including at the same donor/acceptor dinucleotide) where the same impact on mRNA splicing is predicted, but mostly using reduced strength level to prevent overweighting of the VUA compared to the original (likely) pathogenic referenced variant. Suggested weights consider whether the referenced variant lies within the same donor/acceptor dinucleotide or at other positions within the same motif, the PVS1 weight applicable to the VUA, and whether the referenced variant is classified as pathogenic or likely pathogenic.  For any application of PS1(Splicing), we suggest that the “established (likely) pathogenic variant” be classified following ACMG/AMP guidelines and preferably following VCEP gene-specific recommendations if available.

##### Table 17: PS1 code weights for variants with the same predicted splicing event as a known (likely) pathogenic variant*

| **Variant Under Assessment (VUA)** | **Baseline computational/predictive code applicable to VUA** | **Position of reference variant compared to VUA** | **PS1(Splicing) code applicable to VUA** | |
| --- | --- | --- | --- | --- |
|  |  |  | **with P reference variant** | **with LP reference variant** |
| Located outside donor/acceptor  ±1,2 dinucleotide positions | PP3 | Same nucleotide | PS1 | PS1_Moderate |
|  | PP3 | Within same donor/acceptor motif (including at ±1,2 positions) | PS1_Moderate | PS1_Supporting |
| Located at donor/acceptor  ±1,2 dinucleotide positions | PVS1 | Within the same donor/acceptor dinucleotide | PS1_Supporting | N/A |
|  | PVS1 | Within same donor/acceptor motif, but outside dinucleotide# | PS1_Supporting | PS1_Supporting |
|  | PVS1_Strong, PVS1_Moderate, or PVS1_Supporting | Within the same donor/acceptor dinucleotide | PS1 | N/A |
|  | PVS1_Strong, PVS1_Moderate, or PVS1_Supporting | Within same donor/acceptor motif, but outside dinucleotide# | PS1_Moderate | PS1_Supporting |

* Prerequisite for all: The predicted event of the VUA must precisely match the predicted event of the known (likely) pathogenic variant (e.g. both predicted to lead to exon A skipping, or both to enhanced use of cryptic site B), AND the strength of the prediction for the VUA must be of similar or higher strength than the strength of the prediction for the known (likely) pathogenic variant. (Likely) pathogenic variant should be assigned classification using VCEP specifications. For an exonic variant, predicted or proven functional effect of missense substitution/s encoded by the VUA and (likely) pathogenic variant should also be considered before application of this code. Donor/acceptor dinucleotide refers to donor and acceptor ±1,2 dinucleotides in reference transcript/s used for curation. Designated donor and acceptor motif ranges should be based on position weight matrices for intron category. For GT-AG introns these are defined as follows: the donor motif, last 3 bases of the exon and 6 nucleotides of intronic sequence adjacent to the exon; acceptor motif, first base of the exon and 20 nucleotides upstream from the exon boundary. Consider other motif ranges for non GT-AG introns.

\# If relevant, splicing data for a pathogenic variant outside the donor/acceptor ±1,2 dinucleotide positions may be used to update a PVS1 decision tree, and hence the applicable PVS1 code for a donor/acceptor dinucleotide variant.

### Appendix K: Recommendations for use of BP5

We do not recommend use of BP5 in its original intended form “Variant found in a case with an alternate molecular basis for disease”, which we describe as co-observation of a variant with a pathogenic variant in another gene explaining disease presentation. There are multiple examples of individuals carrying pathogenic variants in more than one known breast cancer predisposition gene. Notably, there is published evidence (Rebbeck et al., 2016; PMID: 27836010) that Individuals who carry both a *BRCA1* and *BRCA2* pathogenic variant, exhibit the features of a BRCA1 pathogenic variant carrier. Given that *BRCA1* and *BRCA2* pathogenic variants are individually associated with a high risk of cancer, the observation that such dual pathogenic variant do not exhibit unusual clinical features indicates that identification of a variant in a case with an alternate molecular basis for disease is not by itself information providing evidence against pathogenicity.

However we will use this code to capture combined LR against pathogenicity, in the context of clinically calibrated evidence types, with sufficient detail to review data sources, types and weights. See Appendix B for further details.
