## Supplemental Section 3 for "Evidence-based recommendations for gene-specific ACMG/AMP variant classification from the ClinGen ENIGMA BRCA1 and BRCA2 Variant Curation Expert Panel": CSpec_BRCA12ACMG-Rules-Specifications_V1.0_2023-04-27.docx

**ACMG/AMP Classification Rules Specified for *BRCA1 & BRCA2***

### Overview of documentation supporting specifications

**Table 1** summarizes the **ACMG/AMP criteria specifications** for *BRCA1* and *BRCA2* variant classification by the Variant Curation Expert Panel (VCEP)**.**

**Figure 1** shows the process to apply bioinformatic codes and considerations for incorporating splicing and functional data depending on variant type and position.

The **Appendices** contain extensive **supporting documentation** to justify weighting and application of ACMG/AMP codes (Richards et al 2015, PMID: 18414213) for the classification of variants in *BRCA1* and *BRCA2*. Where relevant, specifications and appendices refer to published and unpublished recommendations provided by members of the ClinGen Sequence Variant Interpretation Working Group (SVI). Where possible, code weights were determined from empirical data, and assigned based on “odds of pathogenicity categories” (**Table 2**), informed from a Bayesian re-analysis of the ACMG/AMP variant classification guidelines (Tavtigian et al 2018, PMID: 29300386), summarized in **Appendix A**.

**Appendix B** describes data types previously utilized for *BRCA1* and *BRCA2* variant classification using a quantitative multifactorial likelihood modeling approach (Goldgar et al 2004, PMID: 15290653; Easton et al 2007, PMID: 17924331; Tavtigian et al 2008, PMID: 8972225; Parsons et al, PMID: 31131967), and categorization according to the IARC 5 tier system (Plon et al, 2008, PMID: 18951446). As appropriate, such likelihood ratio based clinical evidence will be converted to an appropriate ACMG/AMP code weight, and applied under PP4 (combined clinical evidence towards pathogenicity) or BP5 (combined clinical evidence against pathogenicity).

**Reference sequences** used for annotation are as follows**:**

*BRCA1****.*** Coding DNA reference sequence from genomic refseq [NG_005905.2](http://www.ncbi.nlm.nih.gov/nuccore/NG_005905.2) (same as LRG 292, Ensembl ENSG00000012048) covering *BRCA1* transcript [NM_007294.4](http://www.ncbi.nlm.nih.gov/nuccore/NM_007294.3) (Ensembl transcript ENST00000357654.9). Exons are sequentially numbered to match the exon descriptions of the MANE transcript (NM_007294.4). Exon numbering of *BRCA1* has historically been according to GenBank [U14680.1](http://www.ncbi.nlm.nih.gov/nuccore/U14680), with exon 4 missing due to a correction made after the initial description of the gene, termed in this document as legacy exon numbering.

*BRCA2****.*** Coding DNA reference sequence from genomic refseq [NG_012772.3](http://www.ncbi.nlm.nih.gov/nuccore/NG_012772.3) (same as LRG 293, Ensembl ENSG00000139618), covering *BRCA2* transcript [NM_000059.4](https://www.ncbi.nlm.nih.gov/nuccore/NM_000059.4) (Ensembl transcript ENST00000380152.8)***.***

**Appendix C** provides an overview of *BRCA1* and *BRCA2* exon structure (Appendix Figures 1, 2), and a summary of protein functional domains, and their clinical importance (Appendix Tables 3, 4). A clinically important functional protein domain is defined as a recognized protein functional domain reported to harbor one or more clinically important residues. These include BRCA1 RING aa 2-101, BRCA1 BRCT repeats aa 1650-1857, BRCA2 PALB2 binding domain aa 10-40, BRCA2 DNA binding aa 2481-3186. Although no missense variants in the BRCA1 Coiled-Coil domain (aa 1391-1424) are yet considered formally classified as pathogenic, given the suspicion of cancer risk association for at least some missense substitutions in this region, it is termed a potentially clinically important functional domain.

**Appendices D to K** document additional information of relevance to ACMG/AMP code assignment (as designated in Table 1).

**Table 3** and related text denotes the **combinations of code strengths required to meet variant classes** (benign, likely benign, likely pathogenic, pathogenic).

**Note:** the proposed classification criteria are set up to differentiate germline high risk variants (associated with cancer risk equivalent to classical pathogenic variants known or predicted to encode a premature termination codon i.e. nonsense or frameshift) from variants with low/no risk. As such, “reduced” penetrance variants associated with moderate risk of cancer may not be reliably distinguished as risk-associated pathogenic variants. e.g. *BRCA1* c.5096G>A p.Arg1699Gln (Spurdle et al 2012, PMID: 22889855)**.** Variants with discordances across multiple evidence types will be considered for classification using a points-based approach (see below), and highlighted for further study using approaches aimed at investigating reduced penetrance and/or partial effect on function/splicing.

#### Table 1: Summary of ACMG-AMP* Criteria Specified for *BRCA1* & *BRCA2*

| **PATHOGENIC CRITERIA** | | | | | |  |
| --- | --- | --- | --- | --- | --- | --- |
| **Criteria** | | **Criteria Description** | | **Specification**  See relevant **Appendix** for full justification. | | **Relevant tables, figures, links and other information** |
| **VERY STRONG CRITERIA** | | | |  | |  |
| PVS1_VariableWeight | | Null variant in a gene where loss of function is a known mechanism of disease. | | In alignment with SVI recommendations for PVS1 code application, evidence strength and description has been separated for different variant types. Apply according to PVS1 flowchart, which considers knowledge of clinically important functional domains. For predicted protein termination codon (PTC) variants, apply with exon-specific weights derived for the PM5_PTC code (see description below).  **See Appendix D** | | **See Table 4,** provided as a separate searchable excel file, for a comprehensive summary of codes applicable for all variants considered against the *BRCA1* and *BRCA2* PVS1 decision trees (initiation, nonsense/frameshift, deletion, duplication, splice site (donor/acceptor ±1,2)) – organized by exon. |
| PVS1_Variable Weight (RNA) | | Null variant in a gene where loss of function is a known mechanism of disease, *as measured by effect on mRNA transcript profile – mRNA assay only* | | Assay measures effect via mRNA only.  Apply as PVS1_Variable Weight, denoted in summary descriptions as PVS1_Variable Weight (RNA).  **See Appendix E** | | See **Figure 1B** for process to apply codes for splicing data, in content of location and predicted bioinformatic impact of the variant, and adaptive weighting according to assay methodology and proportion of functional transcript retained. |
| **STRONG CRITERIA** | | | |  | |  |
| PS1_Variable Weight | | Same amino acid change as a previously established pathogenic variant regardless of nucleotide change.  *Also to be applied in the context of splicing predictions*. | | Apply **PS1**, for predicted missense substitutions, where a previously classified pathogenic variant is considered to act via protein change (no confirmed or predicted effect on mRNA splicing (SpliceAI≤0.1)).  Apply **PS1_Moderate**, for predicted missense substitutions, where previously established likely pathogenic variant is considered to act via protein change (no confirmed or predicted effect on mRNA splicing (SpliceAI≤0.1)).  Apply **PS1_Variable Weight**, for exonic and intronic variants with same predicted impact on splicing, as a previously classified (likely) pathogenic variant. Vary weight depending on relative positions, and confidence in classification of the reference variant.  **See Appendix E, J, K.** | | For both missense and splicing scenarios, (Likely) Pathogenic variant classification should be assigned using VCEP specifications.  For application of PS1 for splicing predictions, **see Table 5.** The predicted event of the VUA must precisely match the predicted event of the known (likely) pathogenic variant (e.g. both predicted to lead to exon A skipping, or both to enhanced use of cryptic site B), AND the strength of the prediction for the VUA must be of similar or higher strength than the strength of the prediction for the known (likely) pathogenic variant. For an exonic variant, predicted or proven functional effect of missense substitution/s encoded by the variant and the established pathogenic variant should also be considered before PS1 code application for splicing prediction. |
| PS2 | | *De novo* (paternity confirmed) in a patient with the disease and no family history. | | **Do not use**  *BRCA1/2*-related cancers occur relatively commonly. No information to calibrate the predictive capacity of *de novo* occurrences. | |  |
| PS3 – mRNA assay only – **Use under alternative code.** | | Well-established *in vitro* or *in vivo* functional studies supportive of a damaging effect *as measured by effect on mRNA transcript profile – mRNA assay only* | | Assay measures effect via mRNA only.  Apply as PVS1_Variable Weight, denoted in summary descriptions as PVS1_Variable Weight (RNA).  **See Appendix E** | | See **Figure 1B** for process to apply codes for splicing data, in content of location and predicted bioinformatic impact of the variant, and adaptive weighting according to assay methodology and proportion of functional transcript retained. |
| PS3 - functional assays | | Well-established *in vitro* or *in vivo* functional studies supportive of a damaging effect | | Assay measures effect via protein only OR mRNA and protein combined.  **See Appendix E** | | See **Figure 1C** for simplified flowchart/s to advise application of codes for functional data, in content of variant type and location within a (potentially) clinically important functional domain.  See **Table 9**, provided as a separate excel spreadsheet, for a comprehensive table of applicable codes using published calibrated functional assay results. |
| PS4 | | The prevalence of the variant in affected individuals is significantly increased compared with the prevalence in controls. | | Case-control studies; p-value ≤0.05 and OR ≥4 (lower confidence interval excludes 2.0).  **See Appendix F** | | Case dataset should be ethnicity and country-matched to control dataset.  If case-control LR estimates are available for a given dataset, these should be used in preference to case-control OR data, under code PP4 (or BP5, if appropriate). |
| PS4_Moderate | | Proband Counting: prior observation of the variant in multiple unrelated patients with the same phenotype, and its absence in controls | | **Do not use Proband Counting as originally described.**  **See Appendix F.** | | Note, personal and family history of cancer may be used as predictors of pathogenicity if derived by clinical calibration, and applied under code PP4 (or BP5, if appropriate). |
| **MODERATE CRITERIA** | | | |  | |  |
| PM1 | | Located in a mutational hot spot and/or critical and well-established functional domain. | | **Do not use**  Considered as component of bioinformatic analysis (PP3/BP4). | |  |
| PM2_Supporting | | Absent/rare from controls in an ethnically-matched cohort population sample. | | Absent from gnomAD (exome and genome).  **See** **Appendix G**  for justification regarding code weight and quality control measures.  Preferred datasets to use:  gnomAD v2.1 non-cancer, exomes  gnomAD v3.1 non-cancer, genomes | | Observation of a variant only once in a gnomAD outbred population is not informative (no code applied). Do not apply for insertion, deletion or delins variants. Do not apply if read depth <25 at region around the variant. |
| PM3_VariableWeight | | For recessive disorders, detected in trans with a pathogenic variant. | | Apply for patient with phenotype consistent with *BRCA1*- or *BRCA2*-related Fanconi Anemia (FA), and co-occurrent variants in the same gene.  Phenotype is considered consistent with *BRCA1*- or *BRCA2*-related FA if:  (i) Increased chromosome breakage (DEB, MMC, or spontaneous) and at least one clinical feature indicative of *BRCA1/2*-related FA, categorized under: physical features, pathology and laboratory findings, cancer diagnosis *≤5yr*.  (ii) Result unknown for chromosome breakage, and at least two clinical features indicative of *BRCA1/2*-related FA under at least two of the three categories: physical features, pathology and laboratory findings, cancer diagnosis ≤5yr.  **See Appendix H** | | Co-occurrent P or LP variant should be assigned classification using VCEP specifications.  Variant under assessment must be sufficiently rare (meet PM2_Supporting, or PM2 not applicable).  See **Table 6** for approach to assign points per proband, and final PM3 code assignment based on the sum of PM3-related points.  For related individuals score only most severe presentation.  Also see **Table 6** for additional stipulations |
| PM4 | | Protein length changes due to in-frame deletions/insertions in a non-repeat region or stop-loss variants. | | **Do not use**  Considered as component of bioinformatic analysis (PP3/BP4). | |  |
| PM5 | | Missense change at an amino acid residue where a different missense change determined to be pathogenic has been seen before. | | **Do not use**  Considered as component of bioinformatic analysis (PP3/BP4). | |  |
| PM5_PTC_VariableWeight | | Repurposing of PM5 code.  Protein termination codon (PTC) variant in an exon where a different proven pathogenic PTC variant has been seen before | | Use to justify additional weight for PTC variants annotated as PVS1.  Only applied to genomic PTC changes (not splicing). Weight determined by exon where the termination codon occurs (may not be the same exon as the variant position).  **See Appendix D.** | | **See Table 4,** provided as a separate searchable excel file, for PM5_PTC codes applicable for predicted termination codon variants - organized by exon. |
| PM6 | | Confirmed de novo without confirmation of paternity and maternity. | | **Do not use**  *BRCA1/2*-related cancers occur relatively commonly. No information to calibrate the predictive capacity of *de novo* occurrences. | |  |
| **SUPPORTING CRITERIA** | | | | | |  |
| PP1_VariableWeight | Co-segregation with disease in multiple affected family members | | **See Appendix I**  Apply weight as per Bayes Score:  PP1 - LR >2.08:1  PP1_Moderate – LR>4.3:1  PP1_Strong – LR>18.7:1  PP1_Very Strong – LR>350:1 | | Recommend use of online tool: <https://fengbj-laboratory.org/cool3/analysis.html>  Additional information, including pedigree formatting, is available at:  <https://fengbj-laboratory.org/cool3/manual.html>  Stipulation: to apply code as Pathogenic Very Strong, VUS should have bioinformatically predicted (or experimentally proven) effect on protein or mRNA splicing. If co-segregation score is from a single family, or several families from an isolated population, assess the possibility of a different causative pathogenic variant. | |
| PP2 | Missense variant in a gene that has a low rate of benign missense variation and where missense variants are a common mechanism of disease. | | **Do not use**  High frequency of benign missense variants. | |  | |
| PP3 | Multiple lines of computational evidence support a deleterious effect on the gene or gene product | | **See Appendix J**  Apply PP3 for missense or in-frame insertion, deletion or delins variants inside a (potentially) clinically important functional domain and predicted impact via protein change (BayesDel predicted Impact).  Apply PP3 for predicted splicing (SpliceAI ≥0.2) for silent, missense/in-frame (irrespective of location in clinically important functional domain) and for intronic variants outside of the donor and acceptor ±1,2 positions. | | See **Figure 1A** for process to apply codes according to variant type, location and predicted bioinformatic impact.  **Missense Predictions** (BayesDel)  **BRCA1:**  Impact ≥0.28  **BRCA2:**  Impact ≥0.30  See footnote** for definition of (potentially) clinically important functional domains. | |
| PP4_Variable Weight | Phenotype specific for disease with single genetic etiology. | | **See Appendix B.**  Breast cancer is very common and has a high degree of genetic heterogeneity (caused by pathogenic variants in numerous genes). Use ONLY  to capture combined LR towards pathogenicity, based on multifactorial likelihood clinical data.  PP4 - LR >2.08:1  PP4_Moderate – LR>4.3:1  PP4_Strong – LR>18.7:1  PP4_Very Strong – LR>350:1  Combined LR 1.00-2.08 is not informative (PP4 not applicable). | | Use in the context of clinically calibrated evidence types, with sufficient detail to review data sources, types and weights.  Published data points may include co-segregation with disease, co-occurrence with a pathogenic variant in the same gene, reported family history, breast tumor pathology, and case-control data. Can also apply for unpublished data, where there is no appropriate ACMG/AMP code. Assign weight based on combined LR for clinical data.  See Table 7 for example applications. | |
| PP5_Variable Weight | Reputable source recently reports variant as pathogenic but the evidence is not available to the laboratory to perform an independent evaluation. | | **Do not use** | |  | |

| **BENIGN CRITERIA** | | |  |
| --- | --- | --- | --- |
| **Criteria** | **Criteria Description** | **Specification** | **Relevant Tables and Figures, text description, caveats** |
| **STAND ALONE CRITERIA** | | |  |
| BA1 | Stand-alone allele frequency. | Above 0.001 (0.1%)  **See** **Appendix G** for justification, including quality control measures.  Preferred datasets to use:  gnomAD v2.1 non-cancer, exomes  gnomAD v3.1 non-cancer, genomes | Apply based on maximum filter allele frequency observed in a gnomAD non-founder population, considering exome and genome data separately.  Do not apply if read depth <20.  Do not apply to well-established pathogenic founder variants. |
| **STRONG CRITERIA** | | |  |
| BS1**,**  BS1_Supporting | Allele frequency greater than expected for disease. | BS1 - above 0.0001 (0.01%) BS1_Supporting - >0.00002 (0.002%) to ≤ 0.0001 (0.01%).  **See** **Appendix G** for justification, including quality control measures.  Preferred datasets to use:  gnomAD v2.1 non-cancer, exomes  gnomAD v3.1 non-cancer, genomes | Apply based on maximum filter allele frequency in a gnomAD non-founder population, considering exome and genome data separately.  Do not apply if read depth <20.  Do not apply to well-established pathogenic founder variants. |
| BS2_VariableWeight | Observed in a healthy adult individual for a recessive (homozygous), dominant (heterozygous), or X-linked (hemizygous) disorder, with full penetrance expected at an early age | Applied in absence of features of recessive disease, namely Fanconi Anemia phenotype.  **See Appendix H** | Co-occurrent P or LP variant should be assigned classification using VCEP specifications.  See **Table 8** for approach to assign points per proband, and final BS2 code assignment based on the sum of BS2-related points.  Also see **Table 8** for additional stipulations. |
| BS3 – ***mRNA assay only***  **Use under alternative code.** | Well-established in vitro or in vivo functional studies shows no damaging effect on protein function *as measured by effect on mRNA transcript profile – mRNA assay only.* | Assay measures effect via mRNA only.  Apply as BP7_Strong (RNA) for intronic, silent, and missense/in-frame variants located outside a (potentially) clinically important functional domain.  **See Appendix E.** | See **Figure 1B** for process to apply codes for splicing data, in content of location and predicted bioinformatic impact of the variant, and adaptive weighting according to assay methodology and proportion of functional transcript retained. |
| For BS3 – functional assays | Well-established in vitro or in vivo functional studies shows no damaging effect on protein function. | Assay measures effect via protein only OR mRNA and protein combined.  **See Appendix E.** | See **Figure 1C** for process to apply codes for functional data, in content of variant type and location.  See **Table 9**, provided as a separate excel spreadsheet, for a comprehensive table of applicable codes using published calibrated functional assay results.  See footnote** for definition of (potentially) clinically important functional domains. |
| BS4_VariableWeight | Lack of segregation in affected members of a family. | **See Appendix I.**  Apply weight as per Bayes Score:  BS4_Supporting - LR 0.23-0.48:1  BS4_Moderate - LR <0.23:1  BS4 - LR <0.05:1  BS4_VeryStrong – LR <0.00285:1 | Recommend use of online tool: <https://fengbj-laboratory.org/cool3/analysis.html>  Additional information, including pedigree formatting, is available at:  <https://fengbj-laboratory.org/cool3/manual.html>  Stipulation: To apply code as Benign Very Strong, assess the possibility of bi-linearity to explain negative co-segregation. |
| **SUPPORTING CRITERIA** | | |  |
| BP1_Strong | Missense variant in a gene for which primarily truncating variants are known to cause disease | **See Appendix J.**  **Apply BP1_Strong** for silent substitution, missense or in-frame insertion, deletion or delins variants outside a (potentially) clinically important functional domain  AND  no splicing predicted (SpliceAI ≤0.1).  *Missense prediction not applicable*. | See **Figure 1A** for process to apply codes according to variant type, location and predicted bioinformatic impact.  See footnote** for definition of (potentially) clinically important functional domains. |
| BP2 | Observed *in trans* with a pathogenic variant for a fully penetrant dominant gene/disorder, or observed in cis with a pathogenic variant in any inheritance pattern | **Do not use**  Applied only in the context of BS2. |  |
| BP3 | In-frame deletions/insertions in a repetitive region without a known function | **Do not use**  Captured by bioinformatic tool prediction, and domain analysis.  **See Appendix J.** |  |
| BP4 | Multiple lines of computational evidence suggest no impact on gene or gene product | **See Appendix J.**  **Apply BP4** for:  Missense or in-frame insertion, deletion or delins variants inside a (potentially) clinically important functional domain, and no predicted impact via protein change or splicing (BayesDel predicted No Impact AND SpliceAI ≤0.1).  Silent variant inside a (potentially) clinically important functional domain, if no predicted impact via splicing (SpliceAI ≤0.1).  Intronic variants outside of the  donor and acceptor ±1,2 positions  AND no predicted impact via splicing (SpliceAI ≤0.1). | See **Figure 1A** for process to apply codes according to variant type, location and predicted bioinformatic impact.  **Missense Predictions** (BayesDel)  **BRCA1:**  No impact ≤ 0.15  **BRCA2:**  No impact ≤ 0.18  See footnote** for definition of (potentially) clinically important functional domains. |
| BP5 | Variant found in a case with an alternate molecular basis for disease | **See Appendix B and Appendix K** **for justification.**  N/A for co-observation: cases with pathogenic variants in two (or more) different known breast–ovarian cancer risk genes have no specific phenotype.  Use ONLY to capture combined LR against pathogenicity, based on multifactorial likelihood clinical data.  BP5_VeryStrong – LR <0.00285:1  BP5_Strong - LR <0.05:1  BP5_Moderate - LR <0.23:1  BP5 - LR 0.23-0.48:1  Combined LR 0.48-1.00 is not informative (BP5 not applicable) | Use in the context of clinically calibrated evidence types, with sufficient detail to review data sources, types and weights. Published data points may include co-segregation with disease, co-occurrence with a pathogenic variant in the same gene, reported family history, breast tumor pathology, and case-control data. Can also apply for unpublished data, where there is no appropriate ACMG/AMP code. Assign weight based on combined LR for clinical data.  See Table 7 for example applications |
| BP6 | Reputable source recently reports variant as benign but the evidence is not available to the laboratory to perform an independent evaluation | **Do not use** |  |
| BP7 | A synonymous (silent) variant for which splicing prediction algorithms predict neither an impact to the splice consensus sequence nor the creation of a new splice site AND the nucleotide is not highly conserved. | **See Appendix J.**  **Apply BP7 for**  Silent variant inside a (potentially) clinically important functional domain, IF BP4 met.  Intronic variants located outside conserved donor or acceptor motif positions  (at or beyond positions +7/-21)  IF BP4 met | Following convention, this code is applied in addition to BP4 (no splicing prediction, Splice AI ≤0.1) to capture the low prior probability of pathogenicity of silent variants.  Nucleotide conservation is not considered relevant.  See **Figure 1A** for process to apply codes according to variant type, location and predicted bioinformatic impact.  See footnote** for definition of (potentially) clinically important functional domains. |
| BP7_Strong (RNA) | Re-purposing of BP7 code.  Well-established in vitro or in vivo functional studies shows no damaging effect on protein function *as measured by effect on mRNA transcript profile – mRNA assay only.* | Assay measures effect via mRNA only.  Apply as BP7_Strong (RNA) for intronic, silent, and missense/in-frame variants located outside a (potentially) clinically important functional domain.  **See Appendix E.** | See **Figure 1B** for process to apply codes for splicing data, in content of location and predicted bioinformatic impact of the variant, and adaptive weighting according to assay methodology and proportion of functional transcript retained. |

* Original ACMG criteria codes and descriptions are as per Richards et al 2008 (PMID: 18414213).

** As justified in the appendices, (potentially) clinically important functional domains are defined as: BRCA1 RING aa 2-101; BRCA1 coiled-coil aa 1391-1424; BRCA1 BRCT repeats aa 1650-1857; BRCA2 PALB2 binding domain aa 10-40; BRCA2 DNA binding aa 2481-3186.

#### Table 2: Pathogenic and Benign code strength assignment based on measured odds in favour of pathogenicity*

| **Code Strength** | **Odds range for Pathogenic codes** | **Odds range for Benign Codes** |
| --- | --- | --- |
| Strong | >18.70 to 350 | <0.05 to 0.00285 |
| Moderate | >4.30 to 18.70 | <0.23 to 0.05 |
| Supporting | >2.08 to 4.30 | <0.48 to 0.23 |
| No evidence | 1.00 to 2.08 | 0.48 to 1.00 |

*LR ranges proposed as consistent with ACMG/AMP qualitative rule strengths (Tavtigian et al., 2018, PMID: 29300386).

### Combination of evidence types for different class tiers

We propose to compare two approaches to combining evidence types.

The first approach, to be applied as default, represents a minor adaptation of the traditional ACMG-AMP classification system (PMID: 18414213), incorporating results from the Bayesian Framework analysis of Tavtigian et al 2018 (PMID: 29300386); criteria will be combined to reach different classes as shown in **Table 2.**

#### Table 3: RULES FOR COMBINING CRITERIA following adapted ACMG-AMP approach

| **Pathogenic** | **Uncertain Significance** | **Benign** |
| --- | --- | --- |
| 1 Very strong AND   - ≥1 Strong OR - ≥1 Moderate OR - ≥2 Supporting | Other criteria shown in this table are not met | 1 Stand-alone |
| 1 Strong AND   - ≥3 Moderate OR - 2 Moderate & ≥2 Supporting OR - 1 Moderate & ≥4 Supporting | The criteria for benign and pathogenic are contradictory, and cannot be resolved using a point-based approach (see below). | 1 Very Strong AND   - 1 Strong OR - 1 Moderate OR - 1 Supporting |
|  |  | ≥2 Strong |
| 2 Strong AND   - ≥1 Moderate OR - ≥2 Supporting |  | 1 Strong AND   - 2 Moderate OR - 1 Moderate & ≥1 Supporting OR - ≥3 Supporting |
| ≥3 Strong |  |  |
| **Likely pathogenic** |  | **Likely Benign** |
| 1 Very strong AND 1 Supporting |  | 1 Strong AND   - 1 Supporting OR - 1 Moderate |
| 2 Strong |  | 1 Moderate AND 1 Supporting |
| 1 Strong AND 1–2 Moderate |  |  |
| 1 Strong AND ≥2 Supporting |  | ≥2 Supporting |
| ≥3 Moderate |  |  |
| 2 Moderate AND ≥2 Supporting |  | 1 Strong, if based on multiple evidence types* |
| 1 Moderate AND ≥4 Supporting |  |  |

* Likely Benign can be assigned based on one Strong Benign code if multiple evidence types contribute to the code assigned. For example: BS4, if multiple likelihood ratios contribute to the combined likelihood ratio used to assign strong evidence; BP1_Strong represents a combination of the following evidence types: variant type, position (domain info) and prediction (splicing).

The second approach, to be applied in the context of contradictory evidence, is the point system recently proposed to simplify scoring and class assignment (Tavtigian et al 2020, PMID: 32720330). Contradictory evidence includes uninformative bioinformatic evidence, PP3 and BP4 not assigned. Point values assigned for the different code strengths were as recommended in the original publication: Indeterminate=0; Supporting=1, Moderate=2, Strong=4 and Very Strong=8 for Pathogenic codes; negative values for Benign codes of same strength. Point ranges to assign class will follow the conservative recommendations in the original publication: Benign ≤-7, Likely Benign -6 to -2, Uncertain -1 to 5, Likely Pathogenic 6-9, Pathogenic ≥10.

**Note:** both of these approaches assume a global prior probability of pathogenicity of 0.10. On this basis, any evidence type reaching 1000:1 odds against pathogenicity is sufficient to classify a variant as stand-alone Benign, while evidence with combined odds of 350:1 against (equivalent to lower bound of Very Strong Benign evidence) is sufficient to classify a variant as Likely Benign.

### Tables and Figures with additional information for lookups

#### Table 4: Summary of codes applicable for variants considered against the *BRCA1* and *BRCA2* PVS1 decision trees.

This Table is provided as a separate file in excel format to facilitate searches and look-ups by variant c. nomenclature, exon etc.

It includes PVS1 and PM5 codes recommended for initiation, nonsense/frameshift, deletion, duplication and splice site (donor/acceptor ±1,2) variants– organized by exon. Full gene deletion is not included as this is considered as Stand Alone evidence for pathogenicity.

**Assign final PM3 code based on sum of PM3-related points from *observations across multiple unrelated individuals*,** as follows:

PM3_Strong = 4 (or more) points; PM3 = 2 points; PM3_Supporting = 1 point.

#### Table 7: Example application of PP4 and BP5 codes based on multifactorial likelihood clinical data points

**Assign final BS2 code based on sum of BS2-related points:** BS2 = 4 (or more) points; BS2_moderate = 2 points; BS2_Supporting = 1 point.

#### Table 9: Summary of BRCA1 and BRCA2 functional assay results reviewed for application of PS3 and BS3 codes.

This Table is provided as a separate file in excel format to facilitate searches and look-ups by variant c. and p. nomenclature. It includes PS3 and BS3 code recommendations and rationale for code application of published functional assays data that has been calibrated, and considered against predicted/reported splicing. Recommendations relating to variant type and predicted/observed splicing are in Figure 1C.

#### **Figure 1:** Application of bioinformatic codes and considerations for incorporating splicing and functional data depending on variant type and position.

**Figure 1A:** Application of bioinformatic codes based on variant type and location.

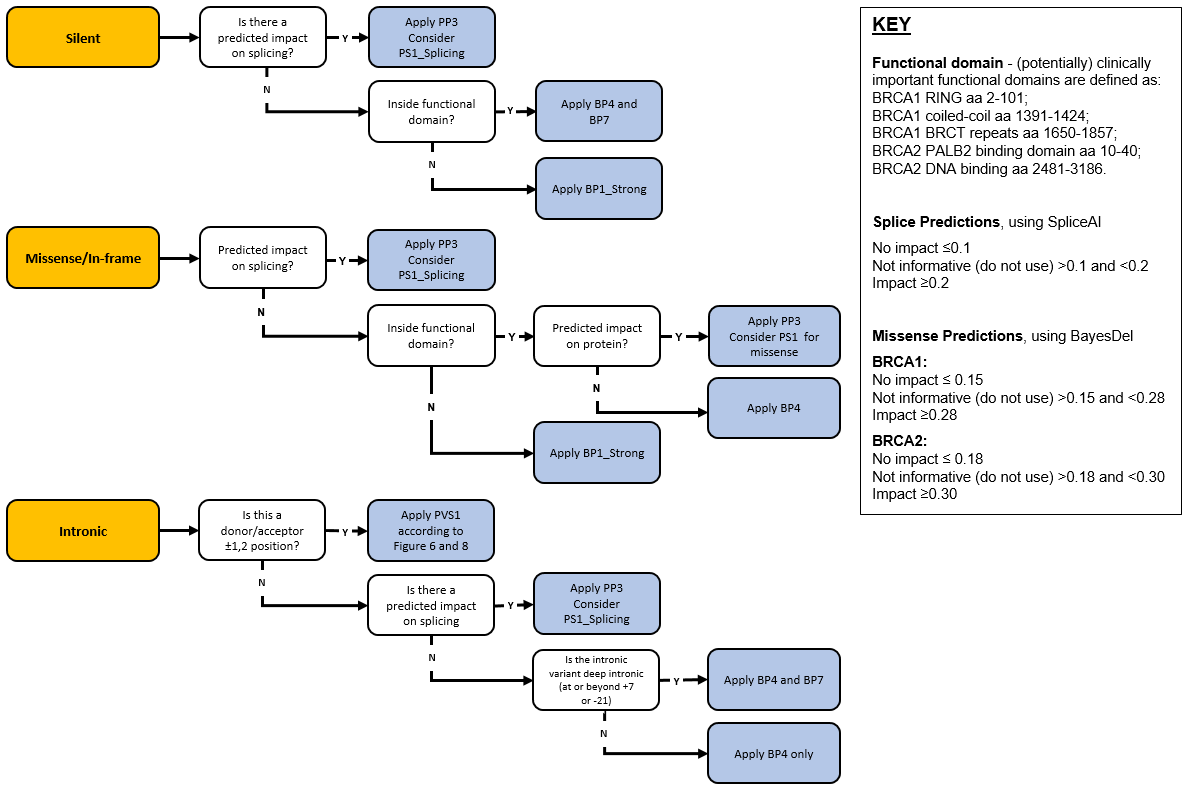

**Figure 1B**: Application of codes based on mRNA assay data.

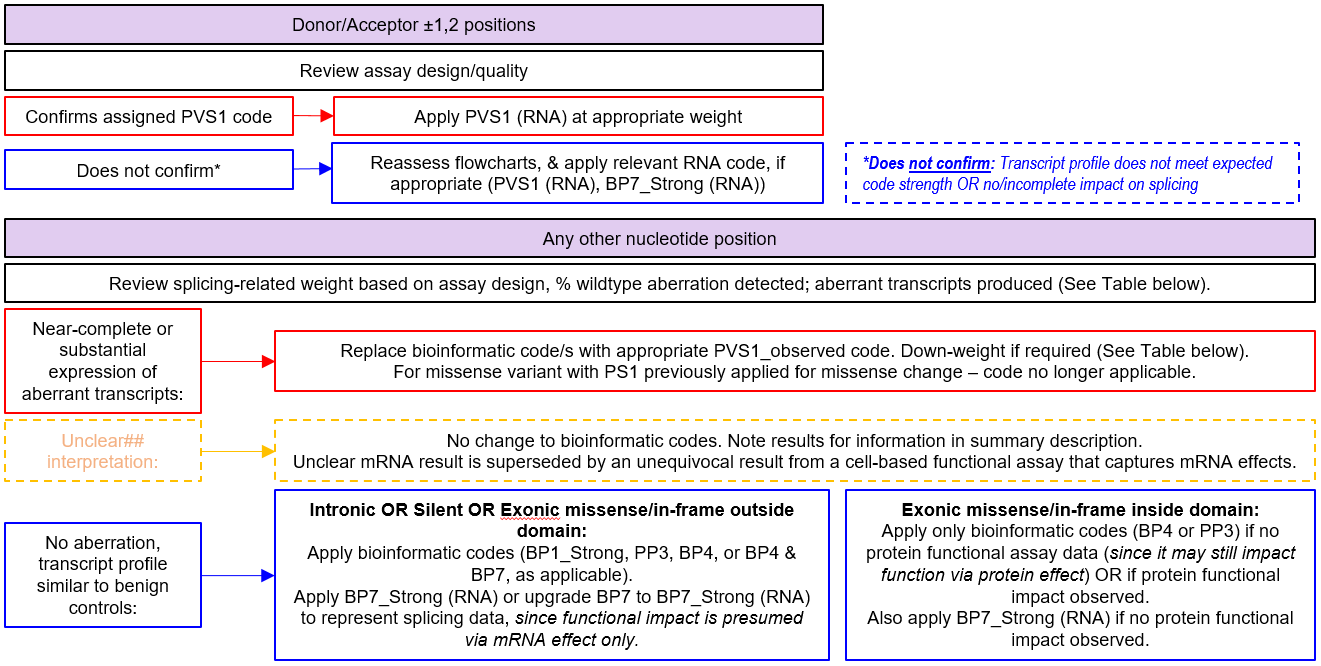

**Figure 1B**: Application of codes based on mRNA data (cont.).
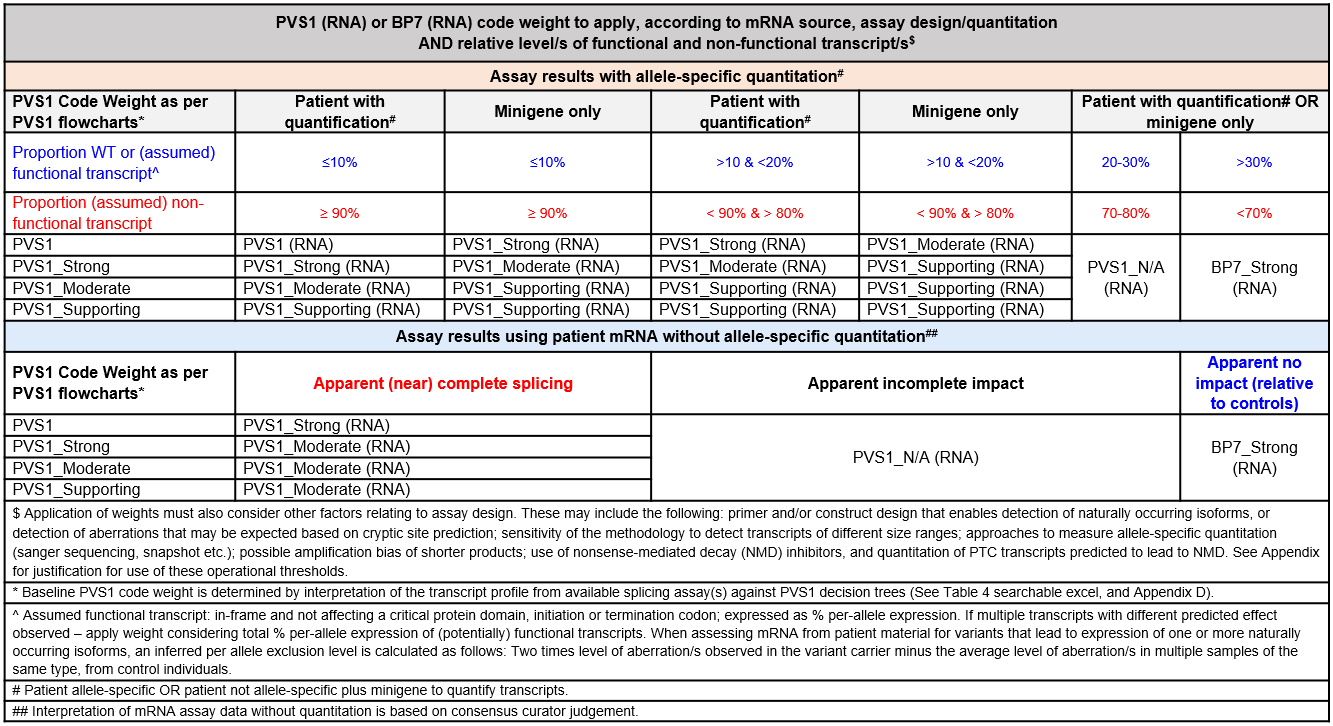

**Figure 1C**: Application of codes based on functional assay data, and variant type and location.

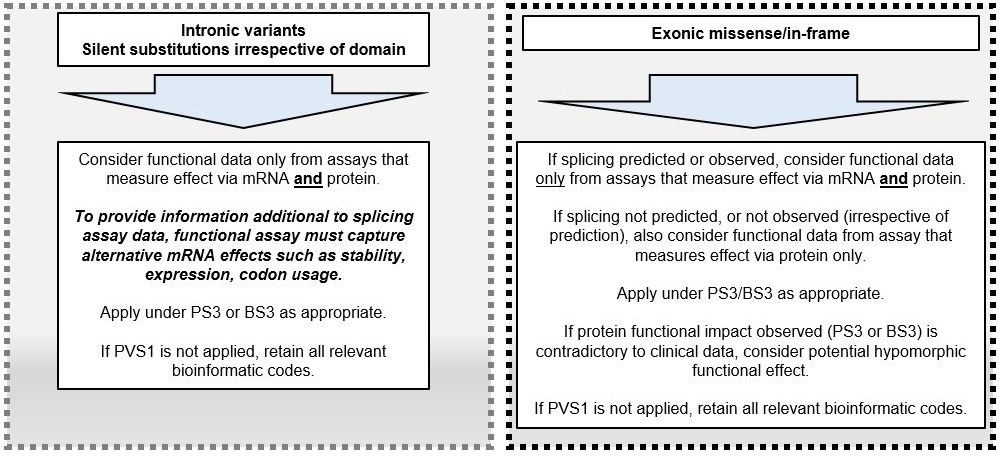
